# Natural history, trajectory, and management of mechanically ventilated COVID-19 patients in the United Kingdom

**DOI:** 10.1101/2020.11.10.20226688

**Authors:** Brijesh V Patel, Shlomi Haar, Rhodri Handslip, Teresa Mei-Ling Lee, Sunil Patel, J. Alex Harston, Feargus Hosking-Jervis, Donna Kelly, Barnaby Sanderson, Barbara Bogatta, Kate Tatham, Ingeborg Welters, Luigi Camporota, Anthony C Gordon, Matthieu Komorowski, David Antcliffe, John R Prowle, Zudin Puthucheary, A. Aldo Faisal, on behalf of the United Kingdom COVID-ICU National Service Evaluation

## Abstract

**Background:** To date the description of mechanically ventilated patients with Coronavirus Disease 2019 (COVID-19) has focussed on admission characteristics with no consideration of the dynamic course of the disease. Here, we present a data-driven analysis of granular, daily data from a representative proportion of patients undergoing invasive mechanical ventilation (IMV) within the United Kingdom (UK) to evaluate the complete natural history of COVID-19.

**Methods:** We included adult patients undergoing IMV within 48 hours of ICU admission with complete clinical data until intensive care unit (ICU) death or discharge. We examined factors and trajectories that determined disease progression and responsiveness to interventions used in acute respiratory distress syndrome (ARDS). Our data visualisation tool is available as a web-based widget (https://www.CovidUK.ICU).

**Findings:** Data for 633 adults with COVID-19 who were mechanically ventilated between 01 March 2020 and 31 August 2020 were analysed. Mortality, intensity of mechanical ventilation and severity of organ injury increased with severity of hypoxaemia. Median PaO_2_/FiO_2_ in non-survivors on the day of death was 12.3(8.9-18.4) kPa suggesting severe refractory hypoxaemia as a major contributor to mortality. Non-resolution of hypoxaemia over the first week of IMV was associated with higher ICU mortality (60.4% versus 17.6%; P<0.001). The reported ideal body weight overestimated our calculated ideal body weight derived from reported height, with three-quarters of all reported tidal volume values were above 6mL/kg of ideal body weight. Overall, 76% of patients with moderate hypoxaemia and 46% with severe did not undergo prone position at any stage of admission. Furthermore, only 45% showed a persistent oxygenation response on prone position. Non-responders to prone position show higher lactate, D-Dimers, troponin, cardiovascular component of the sequential organ failure assessment (SOFA) score, and higher ICU mortality (69.5% versus 31.1%; P<0.001). There was no difference in number of prone sessions between survivors and non-survivors, however, patients who died without receiving prone position had a greater number of missed opportunities for prone intervention (7(3-15.5) versus 2(0-6); P<0.001).

**Interpretation:** A sizeable proportion of patients with progressive worsening of hypoxaemia had no application of and were refractory to evidence based ARDS strategies and showed a higher mortality. Strategies for early recognition and management of COVID-19 patients refractory to conventional management strategies will be critical to improving future outcomes.

**Research in context:** *Evidence before this study:* Beyond the regular literature expertise of our consortium, we enhanced our literature review - due to the fast-evolving Covid-19 publication situation-by searching PubMed for articles published in English or with English language abstracts on October 26, 2020 (and before), with the terms “mechanical ventilation”, “prone position”, “AND (“coronavirus” OR “COVID-19”). Studies including patients not receiving ventilation were excluded, as were those reporting on paediatric and single-centre populations. Note, that neither of those studies analysed the data with respect to the temporal evolution of patients and at our level of granularity. Only four multicentre studies reported detailed ventilator settings and outcomes in ventilated patients with COVID-19. All studies showed only ventilator settings with restricted time points either on admission or the first 4 days of admission. None enabled granular visualisation and analysis of longitudinal ICU trajectory and management.

*Added value of this study:* This study provides a comprehensive analysis and visualisation of routine clinical measurements tracking the whole ICU time course of patients undergoing invasive mechanical ventilation for COVID-19. Mechanically ventilated patients with COVID-19 have a different natural history and trajectory from descriptions of non-COVID ARDS patients, not predictable from admission physiology. Refractory hypoxaemia is an attributable factor associated with poor outcomes in Covid-19 and hence, understanding of use and utility of evidence-based ARDS interventions is clinically crucial. Opportunities to apply prone positioning appropriately are frequently missed, application of high levels of PEEP, and higher tidal volume delivery than planned is common. Lack of responsiveness to advanced ARDS management is associated with hypercoagulation and cardiovascular instability. These data may help homogenise future clinical management protocols and suggest change-of-practice trials.

*Implications of all the available evidence:* This study shows that disease progression in Covid-19 during the first surge occurred more frequently and for longer than other forms of respiratory failure from pre-Covid19 studies. Furthermore, variations in clinical practise occur across sites which may benefit from standardisation of evidence-based practise. Patients that do not resolve hypoxaemia over the first week have a significantly higher mortality, and, crucially, that a significant proportion are refractory to prone interventions and show variability in responses to PEEP changes. Opportunities to implement prone position were missed in many patients and this was compounded with its reduced effect on oxygenation with delayed application. This lack of responsiveness is related to indices of inflammation, thrombosis, and cardiac dysfunction suggesting that pulmonary thrombosis could influence prone responsiveness and should be pro-actively investigated in the setting of refractory Covid-19 ARDS. Prediction of failure to resolve or respond to ARDS interventions could further focus research on this group with worse outcome.

## Introduction

Coronavirus Disease 2019 (COVID-19) caused by the Severe Acute Respiratory Syndrome Coronavirus-2 (SARS-CoV-2) was declared a global pandemic on March 11, 2020 by the World Health Organisation. COVID-19 related severe acute hypoxemic respiratory failure invariably leads to intensive care unit (ICU) admission. These patients fulfil Acute Respiratory Distress Syndrome (ARDS) criteria ^1–4^. However, there remain major uncertainties around the extent of pathological and physiological differences between COVID-19 related ARDS and other causes of ARDS. This ambiguity leads to an ongoing debate on the application of existing evidence-based ARDS management to COVID-19 patients ^5^. Reports based on compliance-based phenotypes and pulmonary angiopathy management in COVID-19 further fuel the uncertainties regarding the clinical management of these acutely unwell patients with a high mortality rate ^6–8^, which we aim to clarify with this data-driven service evaluation.

Pre-COVID evidence-based guidelines for ARDS management include lung-protective ventilation, prone positioning, a conservative fluid strategy, with the option of open lung strategy and neuromuscular blockade (NMBA) ^9^. Patients with refractory respiratory failure should be considered for timely escalation to extracorporeal membrane oxygenation (ECMO) support ^10,11^. Moreover, reports suggest that real-world compliance with evidence based ARDS management strategies is difficult at a system level ^12^, particularly during times of workforce stress, such as during the first wave of the pandemic. Furthermore, these interventions are implemented at various stages of ARDS progression and are time-sensitive over the natural history of illness ^13,14^. Monitoring of dynamic responsiveness to interventions is fundamental to clinical practise in critical care and is increasingly facilitated by advanced analytics ^15^. Whilst there have been reports of the epidemiological characteristics of hospitalised patients with COVID-19 admitted to intensive care in the UK ^16,17^, there has been limited analysis of temporal clinical data combining use of and response to ARDS management strategies.

Accordingly, we undertook a cohort study across a representative set of intensive care units in the United Kingdom, to report the natural history of mechanically ventilated COVID-19 patients. Our specific aims were to ascertain use, compliance, duration and effect of established ARDS management strategies and, to define, from routine clinical measurements, factors associated with disease progression, responsiveness to prone positioning, and mortality.

## Methods

### Study design

We performed a multicentre, observational cohort study in patients with SARS-CoV-2 infection who required mechanical ventilation for severe COVID-19 infection in the United Kingdom.

### Exposure

Adult patients (aged ≥18 years) with laboratory confirmed SARS-CoV-2 infection who required invasive mechanical ventilation (IMV) in the United Kingdom between March 1^st^ and August 31^st^, 2020. Only patients transferred to the study sites within 48 hours of intubation were included, and patients progressing to ECMO were excluded due to the nature of ECMO provision in the UK.

### Ethical approval

The United Kingdom Health Research Authority determined that the study be exempt from review by an NHS Research Ethics Committee. Each site registered the study protocol as a service evaluation. The “Strengthening The Reporting of Observational Studies in Epidemiology” statement guidelines were applied (see supplementary appendix pages 4-5) ^18^.

### Data collection and procedures

To manage the considerable daily data flow we set up a standardised data processing pipeline where only routine, pseudonymised data were collected with no change to clinical care. In brief, the case report form captured admission demographics, twice daily (8am and 8pm) respiratory physiology and blood gas results, daily ARDS interventions, daily COVID-19 interventions, daily blood results and outcome status. Table S1 lists the participating sites. Patients were identified through daily review of paper or electronic medical records using a standardised case record form (CRF), with retrospective and prospective data collection permitted. Data were extracted from either electronic healthcare records (EHRs) or paper-based records into the COVID-ICU secure REDCap database (REDCap v10.0.10; Vanderbilt University, US).

### Missing data and imputation

We made the heuristic decision of setting the threshold of data completeness (i.e. missingness) to balance of patients we could include against the number of variables. We defined this by examination of the available variables in the first 48 hours of admission or the last 36 hours before prone or the first 36 hours after prone. If in these 3/4 twelve-hour measurement points, all were missing, then we counted this patient as ‘missing’ data. The missingness is thus the percentage of patients where there is no measurement in this 36/48-hour window for a modality. Percentage of missing data per modality are shown in Table S2, and details of missing data are shown in Table S18. Data imputation was applied using k-nearest neighbours’ algorithm. We ran the imputation with a k of 3, 5, and 7 both on the continuous variable and on the quartile categorization. The maximal odds ratio difference between the imputation approaches for each variable was 0.04 (IQR 0.03-0.07) and had no effect on the significance. All reported results are based on 5-nearest neighbours’ imputation on the quartile categorization.

### Statistical analysis

Descriptive variables are expressed as percentage, or median and interquartile range (IQR), as appropriate. Continuous variables were analysed with Mann Whitney U or Kruskall Wallis tests, as appropriate. Categorical variables were compared using Fisher’s exact test or the Chi-square test for equal proportion, as appropriate. All statistical tests were 2-sided and *p*≤0.05 was considered statistically significant. The incidence and duration of interventions as well as ventilation settings were analysed and reported to current strategies e.g., low tidal volume ventilation and ARDSNet Positive End Expiratory Pressure (PEEP) tables. The analysis for mortality across the surge was assessed in quartiles of patients admitted with the median as the peak of the surge (peak: 31^st^ March; median: 1^st^ April 2020). We defined an intervention period as a daily application of the intervention with a day of no intervention defining the end of the current period. For group-wise analysis, the outcome of the therapies was measured as categorical variables of “Mild, Moderate, or Severe”, “Survival or Death”, “resolver or non-resolver”, and “prone responder or non-responder”. The severity of hypoxaemia was categorised as per Berlin Definition criteria ^19^: mild hypoxaemia (PaO_2_/FiO_2_>26 kPa); moderate hypoxaemia (PaO_2_/FiO_2_: 26.7-13.3 kPa); and severe hypoxaemia (PaO_2_/FiO_2_<13.3 kPa). To evaluate features associated with progression of hypoxaemia, we analysed evolution of hypoxaemia over the first 7 days of invasive mechanical ventilation and categorised them into two groups, “resolvers” and “non-resolvers”. Patients who changed from severe to moderate; severe to mild; moderate to mild; remained mild or got discharged from ICU were considered “resolvers” while those who changed from mild to moderate; mild to severe; moderate to severe, remained moderate or severe, or died, were considered “non-resolvers”. We further considered the longer-term effect on PaO_2_/FiO_2_ after prone positioning and defined prone responsiveness as maintenance of a mean PaO_2_/FiO_2_ >20kPa over 7 days after the first prone episode. Finally, we defined a prone window as a PaO_2_/FiO_2_<20kPa, with an FiO_2_≥0.6, a PEEP≥5cmH_2_O ^20^ to assess opportunities to apply the intervention. Prone windows were measured at 8am and 8pm with the ventilator and arterial blood gas evaluation.

### Logistic regression models

Multivariate logistic regression models were applied (with screening univariate, p<0.1) to each outcome variable to test associations with independent variables. The full list of variables tested for inclusion in these models is shown in Table S2, but only variables with less than 40% missingness were included in each outcome model (missing value analysis in the relevant time points is shown in Table S2). Variables that showed clinical overlap (e.g. SOFA renal and creatinine) had one variable excluded. A data driven approach to collinearity was not taken as many clinical variables associated with each other due to relationships with severity of illness. A full correlation matrix can be found in the supplementary figures. For all outcomes, only patients with more than 80% of the variables were included in the models. Accordingly, up to 20% of the data were missing and thus were imputed. Data were assumed to be missing at random (MAR) owing to the nature of different personnel at many different sites completing each data entry (the full missing value analysis, by site and by day, is shown in Tables S17 and S18). To enable interpretable and comparable odds ratios, all continuous variables were transformed to categorical by splitting them into quartiles. Accordingly, the odds ratio is the risk increase per quartile increase in the measurement. For age, the odds ratio is the risk increase per decade increase; for SOFA scores, the odds ratio is the risk increase per unit increase in the SOFA score; and for binary variables (e.g., gender, comorbidities) the odds ratio is the risk increase of being positive (e.g. being male, having comorbidity).

### Statistical analysis of natural history and management

The association between the change over time of each independent variable and the outcome measures was tested in repeated measures (rm) ANOVA. For the survival and first week resolver outcome, rmANOVA was applied on the physiology variables over the first week of mechanical ventilation, while for the prone responder outcome, it was applied on the physiology variables over a week from the day before the first PP episode. The rmANOVA was applied separately to each physiology variable, and for each variable, only patients with more than 80% of the variable’s measurements over that week were included in the model. Variables for which fewer than 30 patients had more than 80% of the measurements were not analysed. To prevent the risk of too many false positive, we accounted for multiple comparisons in the interaction statistic by controlling the false discovery rate (FDR).

Analyses were carried out using MATLAB (MathWorks Inc., Natick, MA). Detailed data science methods are described in the supplementary appendix.

## Results

### Clinical characteristics on ICU admission

A total of 633 mechanically ventilated patients admitted to 13 UK National Health Service (NHS) Trusts with 18 ICU sites between 01 March 2020 and 31 August 2020 had complete daily data up to ICU death or discharge (figure 1). The variation in admissions amongst ICUs is shown in supplementary table S1. Baseline demographics were similar to the Intensive Care National Audit and Research Centre cohort ^23^ with an ICU survival of 57.7% (figure 1, Tables S3, and S4). Median duration of symptoms prior to ICU admission was 8 (6-12) days. In total, 25.1% of patients were transferred within 48 hours of intubation from another ICU and were included in the analysis. Transfers over 48 hours after intubation were excluded for the purposes of this analysis.

**Figure 1.**
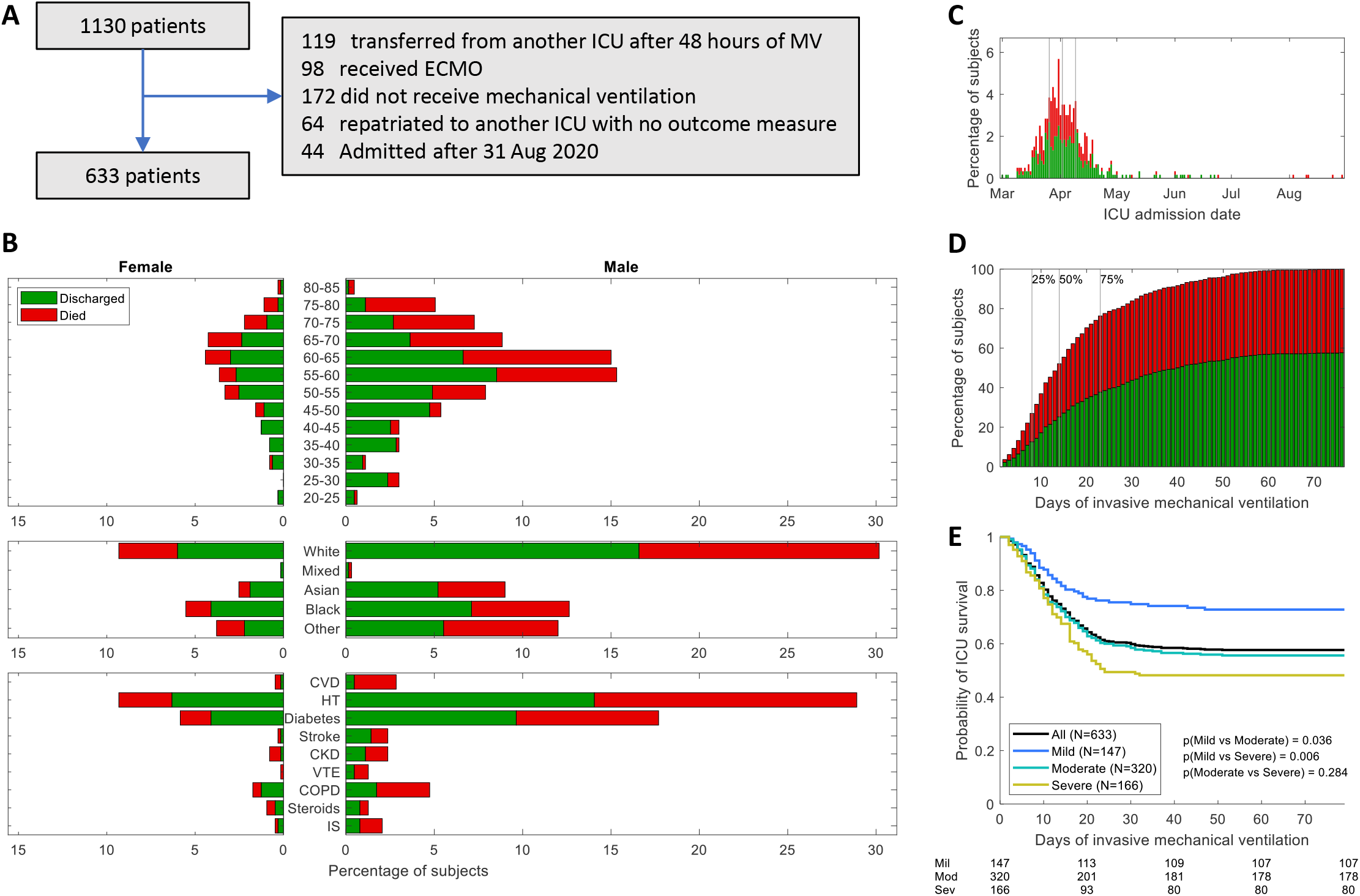
(**A**) Study population flowchart. (**B**) Age, ethnicities, and pre-admission co-morbidities of COVID-19 patients undergoing invasive mechanical ventilation [Cardiovascular Disease (CVD); Hypertension (HT), Chronic Kidney Disease (CKD), Venous thromboembolism (VTE), Chronic Obstructive Pulmonary Disease (COPD), Immunosuppression (IS). (**C**) Admission dates to intensive care unit (ICU) across first pandemic surge; Grey vertical lines represent quartiles based on number of patients admitted. (**D**) Outcome to ICU admission. Grey vertical lines labelled 25%, 50%, 75% indicate the time points (8, 14 and 23 days, respectively) by which the stated proportion of patients that were either discharged or deceased. (**E**) ICU survival curves for the different admission severities of hypoxaemia following the 3 ARDS severity groups as defined in the main text. [(B, C, D) Discharged (green) and died (red) distributions are stacked one over the other. Percentages are out of the total number of patients (N=633).]

On initiation of mechanical ventilation, the severity of mild, moderate and severe hypoxaemia was 23.2%; 50.6%, and 26.2%, respectfully, with an expected mortality gradient (figure 1). Increased severity was associated with increased intensity of mechanical ventilation, severity of organ failure (including dynamic respiratory system compliance, oxygenation index, and ventilatory ratio), and increased application of interventions (table 1). Time series analyses showed that admission severity groups generally maintained the same severity over the first 7 days whereas all other parameters were of no clinical or statistical relevance (figure S4).

**Table 1.**
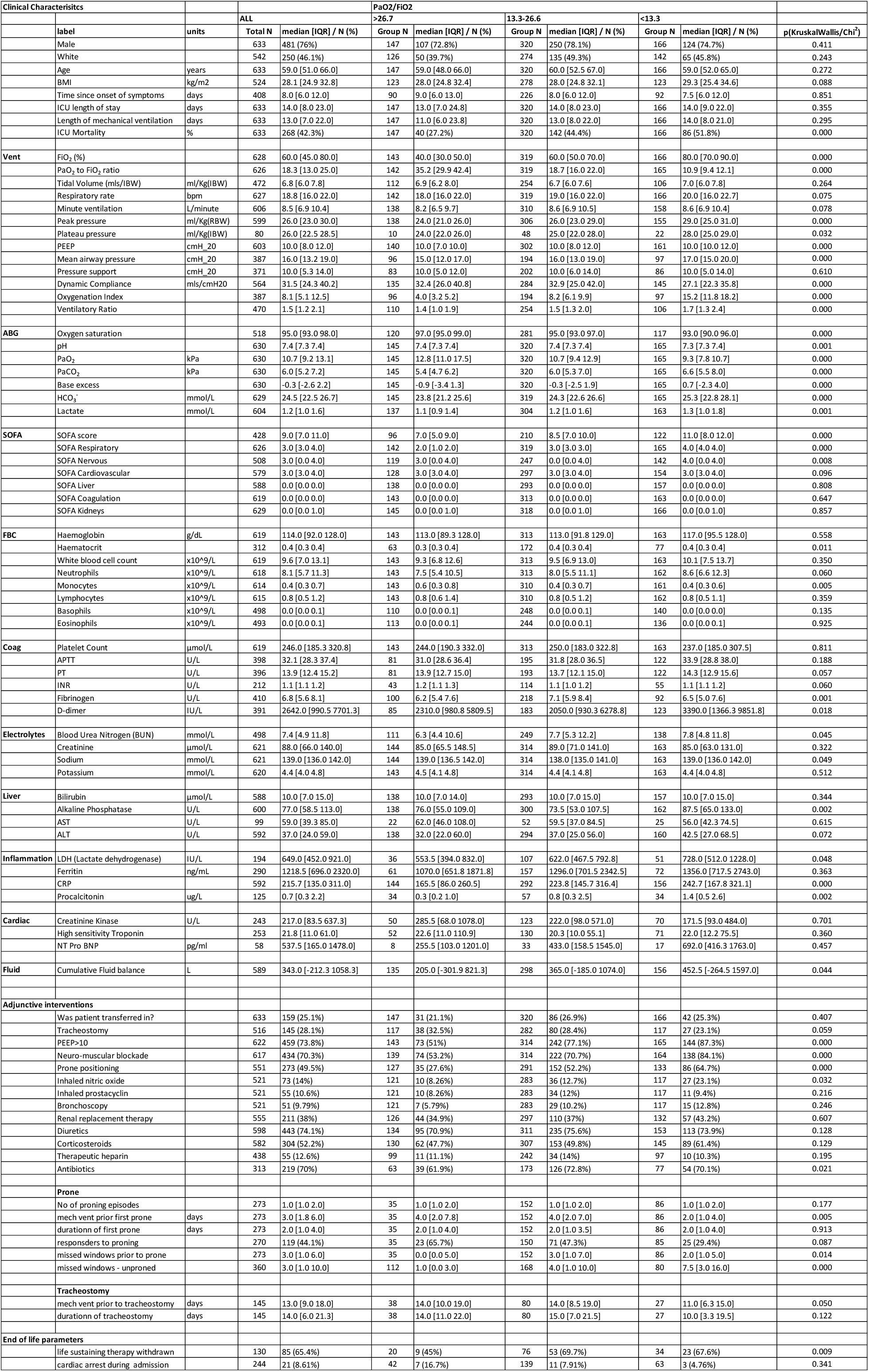
Clinical and physiological characteristics, outcomes and interventions according to severity of hypoxaemia on admission.

### Application of interventions

The application of ARDS interventions are sequential as part of routine management whereby lack of response leads to further interventions. Our web-based tool enables interactive exploration and visualisation of this sequence within the patient journey (https://www.coviduk.icu/). Application of PEEP>10cmH_2_O (74%), continuous NMBA (70%), prone position (50%), inhaled nitric oxide (14%), inhaled prostacyclin (11%), and antimicrobial usage (70%) increased with increasing admission severity of ARDS (table 1). All reported percentages are out of the sub-group of patients for which we have the full records for each intervention (table 1). Figure 2 shows the time of starting and duration of daily recorded interventions. The application, median start date and duration of the first episode of each intervention is shown in table S5. NMBA was commenced on admission (1[0-3] days) and lasted 4(1-7) days. Prone position was applied on day 2(1-5) and lasted 2(1-4) days. Inhaled nitric oxide and prostacyclin were commenced on day 6(3-9) and 7(3-15) and were continued for 4(2-7) days and 3(1-7) days, respectively. Tracheostomy was performed in 29% at a median 14(9-18) days, predominantly in those patients likely to survive (40% versus 10.9%; P<0.001). Diuresis was utilised in 74% and applied on day 1(1-3) and lasted 3(1-5) days. Renal replacement therapy was utilised in 38% of patients with a median commencement on day 3(1-6) after IMV, and a median duration of 5(3-11) days. Anti-microbial prescribing was common in 70% of patients and were administer on or before day of admission, often lasting 6(4-9) days.

**Figure 2.**
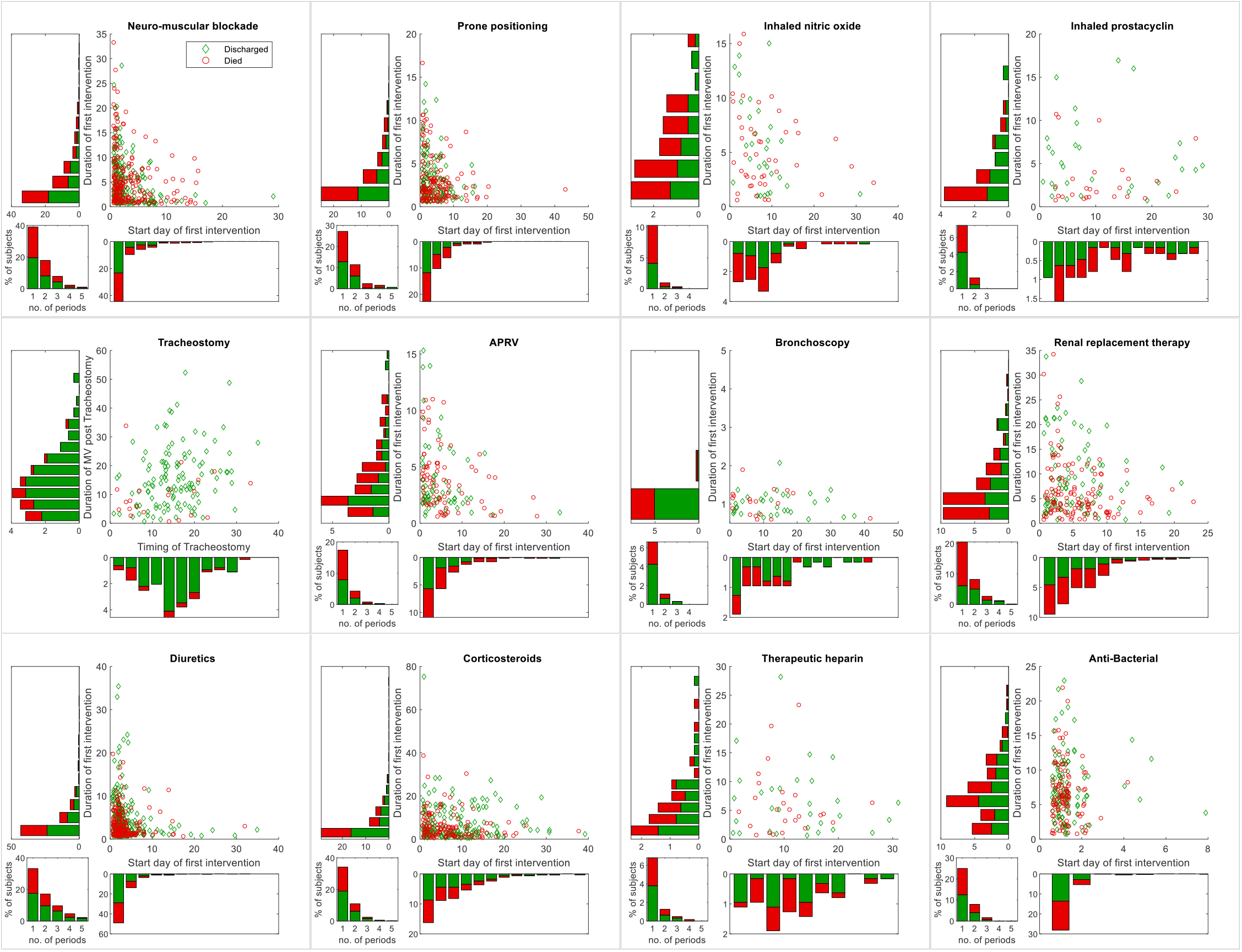
Visualisation of the variability of when and how long interventions were applied for and their associated patient outcome (green - discharge, red - deceased) showing data in a separate boxed panel for each intervention. Each boxed panel contains a scatter plot of the number of days of invasive mechanical ventilation (IMV) at which first intervention was applied (x-Axis) vs duration of the first period of that intervention in days (y-Axis). Parallel to the respective axis we show the marginal histogram of the data points in the scatter plot (e.g. the histogram for start of first intervention on the x-Axis). In the bottom left of the boxed panel we show the histogram of the number of repeated intervention periods a patient underwent (see main text for details). See Table S5 for an overview of the interventions and their respective use statistics. Most patients will have had multiple interventions, see our online interface (www.covidUK.icu) to explore the interactions between interventions.

The reported ideal body weight overestimated our calculated ideal body weight derived from reported height (http://ardsnet.org) in 92.6% of patients (Figure 3). Hence, median tidal volume per kg on actual ideal body weight was 7.0 [IQR 6.0-8.4] mL/kg across all breaths and 5.6 [IQR 4.7-6.6] mL/kg on reported ideal body weight. Three-quarters of all reported tidal volume values were above 6mL/kg of ideal body weight (figure 3). Survivors and non-survivors showed the same distribution of tidal volume variation. Over 65% of reported PEEP values were set outside +/-1cmH_2_O and 53% set outside +/-2cmH_2_O of the ARDSNet PEEP-FiO_2_ tables (figure 3). Patients with BMI<40 had a higher set PEEP than recommended by the PEEP-FiO_2_ table. In contrast, patients with BMI>40 had a lower set PEEP than recommended by the PEEP-FiO_2_ table. Changes in PEEP were widespread over the first 7 days of IMV with both increases and decreases leading to unpredictable changes in PaO_2_/FiO_2_ (Figure 3).

**Figure 3.**
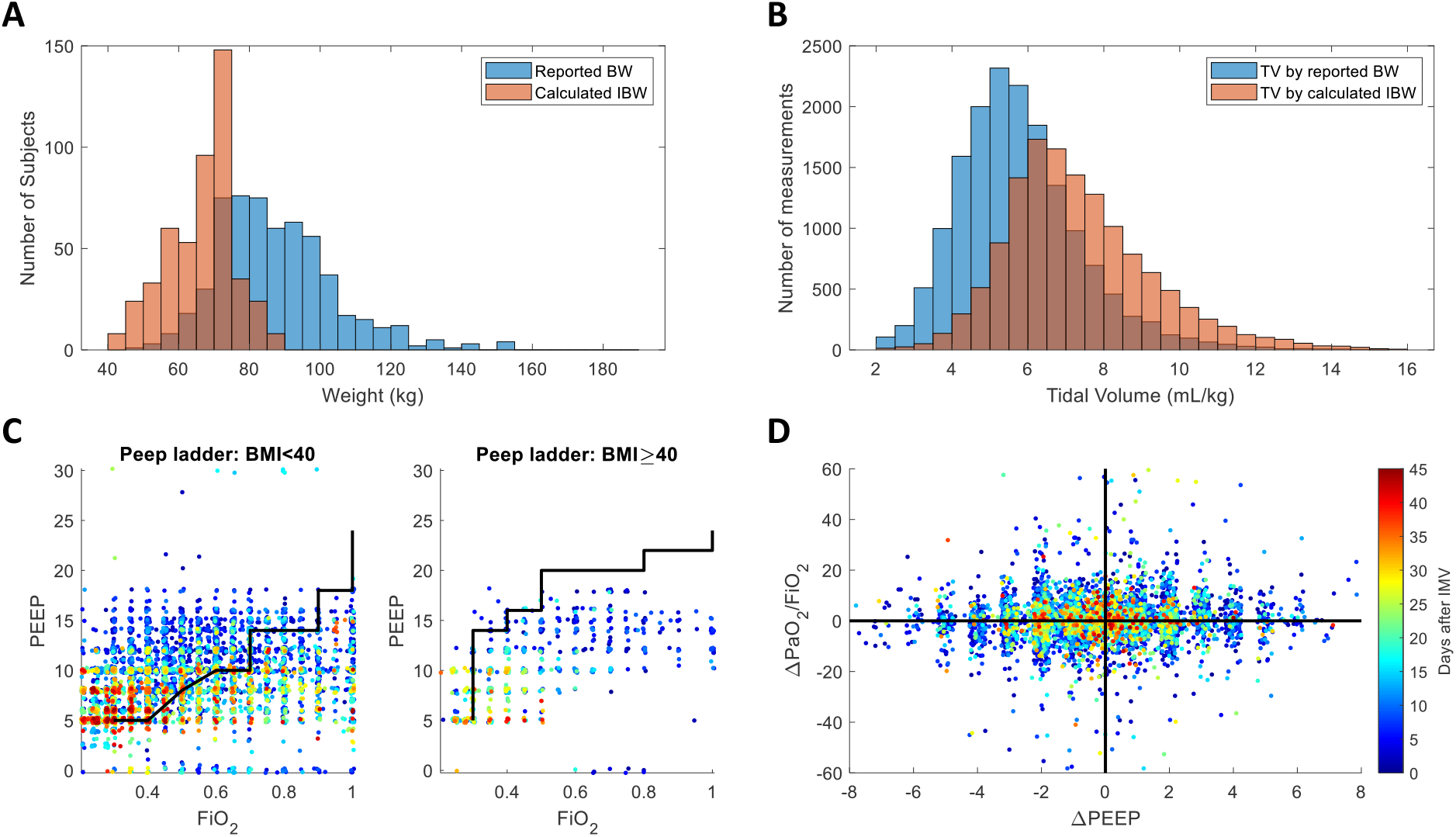
Overview of weight effect on tidal volume and PEEP management. (**A**) The distributions of reported (light blue) and calculated ideal (light red) body weights highlighting systematic differences. (**B**) The distributions tidal volumes in ml/Kg for reported and ideal body weights. (**C**) The management of PEEP as a function of FiO_2_ plotted in a scatter plot for non-morbidly obese patients (BMI<40, left) and morbidly obese (BMI>=40, right). Each plot shows the pre-COVID recommended PEEP ladder of ARDSNet (solid black line) against actual data points showing clearly visible departure from recommended pre-COVID PEEP ladder. Data points are colour coded by Days after initiation of IMV (see colour bar in the next sub-figure). (**D**) Visualisation of the changes in PEEP against the change in PaO_2_/FiO_2_ ratio. The changes are measured across two adjacent time points with the PEEP change being introduced at some point between the two time points. Data points are colour coded by days after initiation of IMV (see colour bar).

### Progression of hypoxaemia in COVID-19-induced respiratory failure

To ensure comparability with pre-COVID ARDS studies we chose an analysis horizon spanning the first 7 days. The movement of patients across severity groups (mild, moderate and severe hypoxaemia) showed deterioration in 31.4% of cases, remained static in 45.1% and resolved in only 23.5% of patients over the first 7 days (Table 2; Figure 4). Overall, progression to a worse PaO_2_/FiO_2_ severity group occurred in twice the number of patients as compared to pre-COVID studies of ARDS (Table 2). ICU mortality in hypoxaemia resolvers was significantly lower than non-resolvers (17.6% versus 60.4%; P<0.001; Figure 4). Differences between resolvers and non-resolvers were apparent in demographic, ventilatory, physiological, and laboratory parameters on admission as shown in Table S6. Unsurprisingly, non-resolution was associated with increased application of interventions. Resolvers were younger (57(47-64) vs 60(54-67) years; P<0.001) and showed a longer duration of symptoms prior to ICU admission 9.0 (7-14) vs 7 (6-11) days (p=0.004). Non-resolvers had a longer duration of IMV and ICU stay (Table S6). Resolvers had prone position applied significantly earlier (2[1-5] vs 4[2-7] days; P=0.007). There were also clinically, and statistically higher admission counts of blood lymphocytes in resolvers, whereas non-resolvers had higher ferritin and cardiovascular SOFA score (Table S6). Time series analysis over the first 7 days showed important clinically significant interactions between groups in indices of respiratory physiology, cumulative fluid balance, acid-base status, renal function, CRP and SOFA score (Figure 4; Figure S2; and Table S7). Multivariate regression showed that increased age and increasing cardiovascular SOFA were associated with worsening of hypoxaemia within the first week of IMV (Figure 4; table S8).

**Table 2.** Progression of hypoxaemia in COVID-19 as compared to pre-COVID ARDS publications. Tables show patient numbers and proportions changing between mild, moderate, and severe hypoxaemia categories from day 1 to day 7 of invasive mechanical ventilation. Table 2a – COVID-ICU database; 2b – LUNG-SAFE study ^19^; 2c – Berlin definition study ^25^.

| COVID-ICU |  |  |  |  |  |
| --- | --- | --- | --- | --- | --- |
|  |  | P/F on Day 7 |  |  | Total |
|  |  | >26.7 | 13.3-26.6 | <13.3 |  |
| P/F on Admission | >26.7 | 51 | 71 | 18 | 140 |
|  | 13.3-26.6 | 55 | 137 | 75 | 267 |
|  | <13.3 | 8 | 58 | 42 | 108 |
|  |  | 114 | 266 | 135 | 515 |

|  |  | P/F on Day 7 |  |  |  |
| --- | --- | --- | --- | --- | --- |
|  |  | >26.7 | 13.3-26.6 | <13.3 |  |
| P/F on Admission | >26.7 | 36.4% | 50.7% | 12.9% | 31.8% |
|  | 13.3-26.6 | 20.6% | 51.3% | 28.1% |  |
|  | <13.3 | 7.4% | 53.7% | 38.9% |  |
|  |  | 23.5% |  |  | 44.7% |

| LUNG SAFE |  |  |  |  |  |
| --- | --- | --- | --- | --- | --- |
|  |  | P/F on Day 7 |  |  | Total |
|  |  | 26.7-40 | 13.3-26.6 | <13.3 |  |
| P/F on Admission | 26.7-40 |  | 184 | 32 | 714 |
|  | 13.3-26.6 |  |  | 140 | 1106 |
|  | <13.3 |  |  |  | 557 |
|  |  |  |  |  | 2377 |

|  |  | P/F on Day 7 |  |  | Overall |
| --- | --- | --- | --- | --- | --- |
|  |  | 26.7-40 | 13.3-26.6 | <13.3 |  |
| P/F on Admission | 26.7-40 |  | 25.8% | 4.5% | 15.0% |
|  | 13.3-26.6 |  |  | 12.7% |  |
|  | <13.3 |  |  |  |  |

| Berlin Definition |  |  |  |  |  |
| --- | --- | --- | --- | --- | --- |
|  |  | P/F on Day 7 |  |  | Total |
|  |  | 26.7-40 | 13.3-26.6 | <13.3 |  |
| P/F on Admission | 26.7-40 |  | 234 | 33 | 819 |
|  | 13.3-26.6 |  |  | 230 | 1820 |
|  | <13.3 |  |  |  | 1031 |
|  |  |  |  |  | 3670 |

|  |  | P/F on Day 7 |  |  | Overall |
| --- | --- | --- | --- | --- | --- |
|  |  | 26.7-40 | 13.3-26.6 | <13.3 |  |
| P/F on Admission | 26.7-40 |  | 28.6% | 4.0% | 13.5% |
|  | 13.3-26.6 |  |  | 12.6% |  |
|  | <13.3 |  |  |  |  |

**Figure 4.**
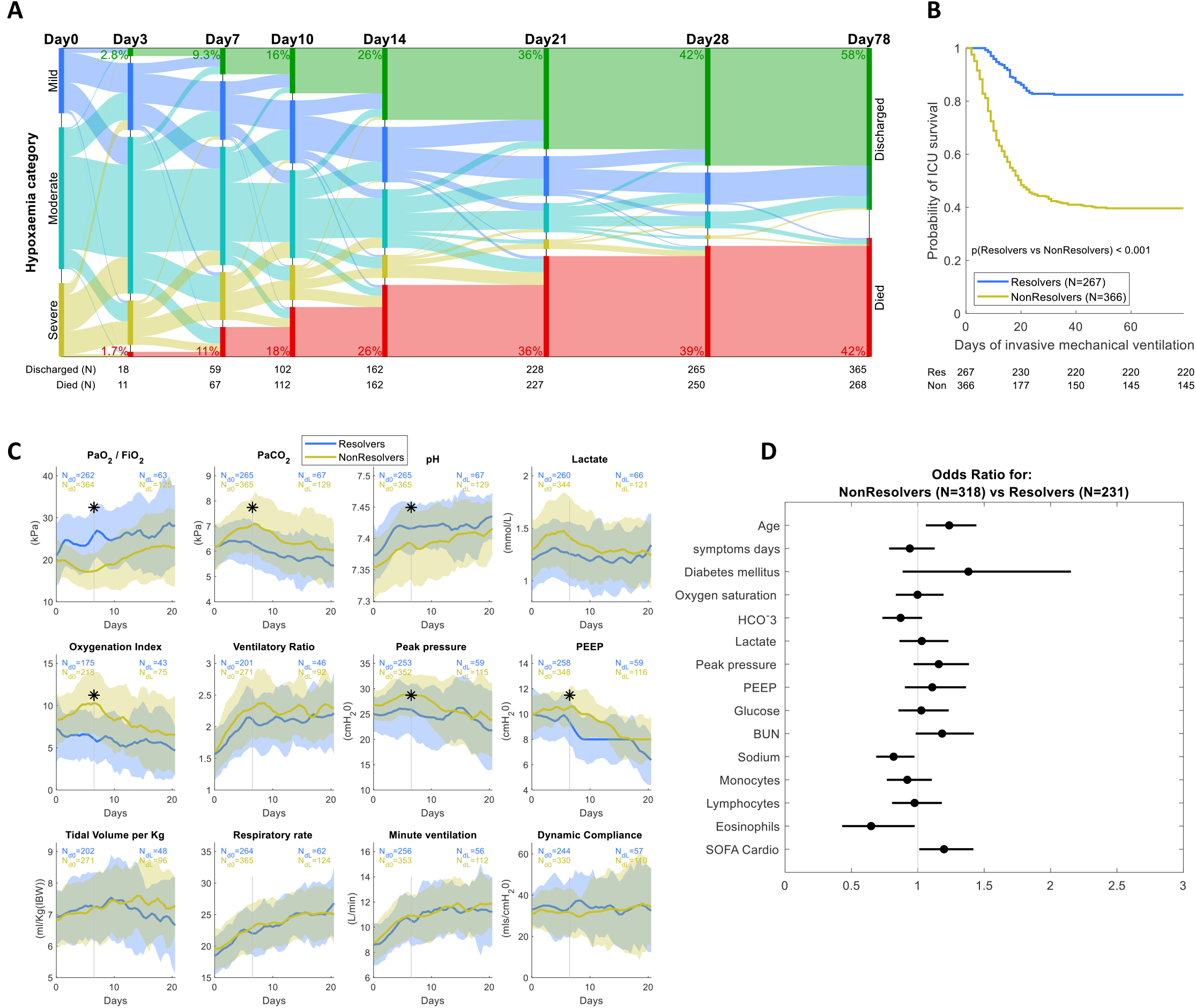
(**A**) Alluvial diagram of patient movements between ARDS severity groups: Mild hypoxaemia (PaO_2_/FiO_2_>26), Moderate hypoxaemia (PaO_2_/FiO_2_: 26.7-13.3), and severe hypoxaemia (PaO_2_/FiO_2_<13.3) and patient outcome (Discharged - green, deceased - red). Each solid bar represents an ARDS severity group at a given number of days since initiation of IMV. Shaded coloured streams between bars represent transitions of patients between the severity groups from one time point to the next, which is either their new severity or their outcome. The height of the bars represents the proportion of patients at that time point (i.e., they stack up to 100%) and the height of a stream field represents the size of the components contained in both bars connected by the stream. (**B**) ICU survival curves for patients who were showing improvement in hypoxaemia category over the first week on IMV (resolvers, light blue) versus deterioration in hypoxaemia category (non-resolvers, yellow). (**C**) Each panel presents the time-series of a physiological measure of resolvers (light blue) versus non-resolvers (yellow) over the first 3 weeks of IMV (*P<0.05 interaction with mixed model ANOVA over the first week of IMV, see table S7). The Solid lines are the group medians and the shaded areas are the semi-interquartile range. The number of subjects decay over time as patients die and discharge and the initial and final numbers available for each measure are presented on the graph. The full set of variables is presented in Figure S2. (**D**) The odds ratio and their 95% confidence interval for Multivariate logistic regression models where a higher odds ratio is the increased likelihood for progression of hypoxaemia for each step increase in the admission variable and physiological measures. Continuous variables were discretely sized by a split into quartiles (see supplementary methods for details and table S8 for the full stats). All variables (of the list in table S2) with less than 40% missingness were included in the model. Subjects with more than 20% missing data were removed from the analysis.

### Responsiveness to prone positioning

In this first wave of the pandemic, prone position was used in 49.5% of patients. Of patients that received of prone position, 63% were pronated once and 37% twice or more. Mortality was 52.7% and 28.4% in those patients who did and did not undergo prone positioning, respectively. Prone position was applied earlier in patients with greater severity. While patients that did not undergo prone position may overall have had milder disease, we found that 76% of these patients who had moderate hypoxaemia and 46% who had severe, at any stage of admission, did not undergo prone position at all. In patients who received no prone position, there was 1(IQR 0-2) prone windows per patient ignored during the first 48 hours and 3(IQR 1-10) during the whole patient journey. In patients who received prone interventions, there were on average 3(IQR 1-6) prone windows per patient before prone initiation that were missed. There was no difference in number of prone sessions between survivors and non-survivors, however, patients who died without receiving prone position had a greater number of missed prone windows (7(3-15) versus 2(0-6); P<0.001; Table S13).

We analysed change in PaO_2_/FiO_2_ over 36 hours around the first prone intervention. The median duration of first prone cycle was 2(1-4) days. Responsiveness to prone position was found to decrease the later the prone episode was initiated after intubation (Figure 5; Spearman r=0.16, P=0.012). We further considered the longer-term effect on PaO_2_/FiO_2_ after prone positioning. Only 44.4% of patients maintained a mean PaO_2_/FiO_2_ > 20kPa over 7 days after the initiation of prone position. Mortality was significantly lower in prone responders than in non-responders (31.1% versus 69.5%, p<0.001 as seen in Figure 5). Of note, 40% of patients that improved hypoxaemia category (resolvers) did not respond to prone position and 36% of patients that worsened hypoxaemia category (non-resolvers) showed an overall improved oxygenation response with prone positioning. Furthermore, only 58% of non-resolvers received prone position and often later in their course. Importantly, there were a higher number of missed opportunities to prone in non-resolvers compared to resolvers (6 [3-13] versus 1 [0-4] windows per patient; P<0.0001; Table S6).

**Figure 5.**
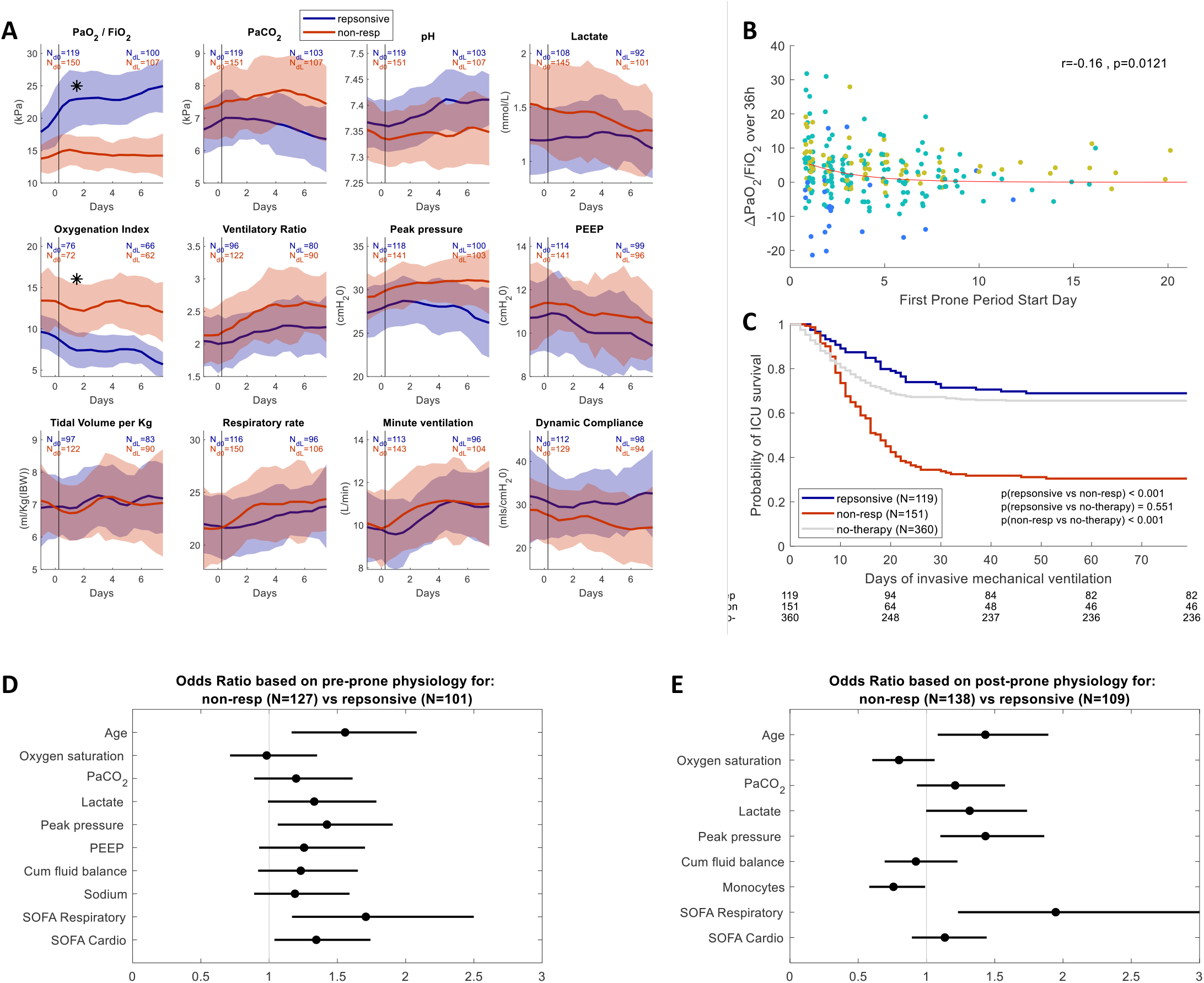
Responsiveness to prone position with responders defined as maintenance of a mean PaO_2_/FiO_2_ >20kPa over 7 days after the first prone episode. (**A**) Each panel presents the time-series of a physiological measure of prone responders (blue) versus non-responders (red) from a day before the first prone manoeuvre to 7 days after (*P<0.05 interaction with mixed model ANOVA over this period, see table S10). The Solid lines are the group medians and the shaded areas are the semi-interquartile range. The number of subjects decay over time as patients die and discharge and the initial and final numbers available for each measure are presented on the graph. The full set of variables is presented in Figure S3. (**B**) Changes in PaO_2_/FiO_2_ ratio over 36 hours around the first prone manoeuvre (from the last measurement before until the first measurement the day after) as a function of the duration of IMV prior to the manoeuvre. The dots are colour coded by ARDS severity prior to the manoeuvre. The red line presents an exponential fit, and the reported r is the Spearman rank correlation. (**C**) ICU survival curves for prone responder (blue) versus non-responder (red) versus patients who received no prone position (grey). (**D-E**) The odds ratio and their 95% confidence interval for Multivariate logistic regression models where a higher odds ratio is the increased likelihood of not responding to prone position for each step increase in the admission variable and in the (**D**) pre-pronation (the last record within 24 hours prior intervention) or (**E**) post-pronation (the first record in the day after intervention) physiological measures. Continuous variables were discretely sized by a split into quartiles (see supplementary methods for details and table S11 and s12 for the full stats for pre- and post-prone respectively). All variables (of the list in table S2) with less than 40% missingness were included in the model. Subjects with more than 20% missing data were removed from the analysis.

Non-responders to prone position tended to be older and showed worse respiratory mechanics, higher lactate, higher cumulative fluid balance, higher troponin, higher D-dimer, and higher Cardiovascular and Respiratory SOFA scores. A prone response was associated with an improved oxygenation index (OI) over the first week of prone position (Figure 5A; Figure S3; Table S10). Whilst there were no significant differences in the duration of IMV prior to the first prone period, the duration of the first period, or the number of future prone periods between responders and non-responders; non-responders had a higher number of missed prone windows than responders (3 [1-7] versus 2 [1-5] windows per patient; P<0.05; Table S6). Moreover, multivariate logistic regression analysis suggests that higher peri-proning lactate, cardiovascular SOFA and respiratory SOFA values were associated with poor prone responsiveness (Figure 5; Table S11).

### Determinants of mortality

Survival to ICU discharge was 57.7%, and admission characteristic differences between survivors and non-survivors are shown in supplementary table S13. Statistically significant interactions were noted in the group-wise ANOVA within several parameters (Figure S5; Table S13) between survivors and non-survivors. With the median being the peak of the surge, the first quartile of patients admitted during the surge had a death rate of 37.3%; the second quartile, 53%; the third quartile, 43.4%; and the last quartile, 35.9%. The multivariate model showed clinical variables independently associated with mortality were higher age (HR 1.95 per decade, 95% CI 1.58–2.4), male gender (HR 2.05, 95% CI 1.17–3.61), higher lactate (HR 1.52 per quartile (0.6 mmol/L), 95% CI 1.21–1.92), and higher SOFA coagulation score (HR 1.95, 95% CI 1.17–3.26) (Figure S5; Table S15). Median PaO_2_/FiO_2_ in non-survivors on the day of death was 12.3(8.9-18.4) kPa suggesting many patients died with (and possibly as a result of) severe refractory hypoxaemia. In those that died, active withdrawal of support occurred in 65% of patients (85/130), in the 13 sites which reported, and unanticipated cardiac arrest occurred in 11% of patients (13/122). Patients who had life support withdrawn had a median age of 64(57-70) years, a length of mechanical ventilation of 11(6-18) days; a last PaO_2_/FiO_2_ of 12.8 (10-19.5) kPa and had a high incidence of prone intervention (72%). 100% of deaths were attributed to COVID-19.

## Discussion

The United Kingdom saw 10,834 patients admitted to 258 intensive care units between 1^st^ February and 31^st^ August with 7702 requiring advanced respiratory support ^17^. We describe a complete natural history of a cohort of patients with COVID-19 undergoing invasive mechanical ventilation (IMV) including detailed interventions for COVID-19-induced respiratory failure. We report differences between 1) admission severities of ARDS, 2) early (< 1 week) resolution and non-resolution; 3) prone responders vs non-responders; and 4) survivors vs non-survivors. Most patients showed disease severity consistent with moderate ARDS, however, severity of hypoxaemia was greater, median length of IMV in survivors was longer as compared to the pre-COVID LUNG-SAFE study (15(8-28) vs 8(4-15) days, respectively) ^13^. This longer period of COVID-19 ventilation has significant implications for provision of critical care bed capacity and further analysis of the impact of COVID-19 therapies on this outcome will be needed ^21^. The COVID-19 pandemic has highlighted the potential for significant deviations in care through several mechanisms including overwhelmed healthcare systems to application of local guidelines from small single centre reports. There was considerable heterogeneity in application of interventions between centres, and our granular analyses of the time course and duration of interventions advise on areas where clinical management could be better standardised and improved. In comparison, the disease progression and applications of interventions was similar to that described by the REVA network ^3^. Finally, this may prompt specific clinical trials to be conducted to evaluate these interventions in this new disease.

### Mechanical ventilation strategies

Covid-19 has triggered immense debate on whether ventilation strategies should be different compared to non-COVID ARDS, due to the severity of hypoxaemia with relatively preserved compliance, and the vasocentric nature of disease ^22^. In our cohort, most patients received lung protective ventilation with tidal volumes less than 8mL/kg and plateau pressures less than 30cmH_2_O. This was despite systematic errors in measurement of height and derived ideal body weight. However, PEEP was set higher than the ARDSNet PEEP table, and changes in PEEP over 12 hours did not equate to improvements in PaO_2_/FiO_2_. Other measures such as the recruitment/inflation index may provide better approaches to PEEP titration ^23^. ARDS, in particular that caused by COVID-19, can show significant pulmonary vascular perfusion defects secondary to significant endothelial inflammation ^7,24^. These data show that a decline in dynamic compliance is associated with increasing severity of disease (despite the application of higher PEEP), highlighting the need for personalised approaches to PEEP application in all causes of ARDS and for it to be assessed in high quality clinical studies during the pandemic. This may have specific relevance to the care of obese patients with ARDS, as UK practise does not seem to comply with high BMI PEEP tables.

### Disease progression

Although differences in admission markers of inflammation (LDH (Lactate dehydrogenase), CRP, ferritin, procalcitonin), thrombosis (platelets, D-dimers) and cardiac dysfunction (troponin and BNP (Brain Natriuretic Peptide)) were seen between patients that survived and died, time series mixed model analysis suggests survivors also showed progressive reductions in CRP and neutrophils, in addition to restoration of lymphocyte and platelet counts. Indeed, survivors demonstrated improvements in dynamic respiratory system compliance along with reductions in ventilatory requirements over the first week. Consistent with other studies, increasing age and male gender were associated with mortality ^16^. Higher lactate and coagulation SOFA were associated with an increased risk of death. This is consistent with recent data showing lower platelet counts being associated with the hyperinflammatory phenotype of ARDS which is associated with worse outcome ^25^.

The extent to which patients resolved was dependent on progression of disease as well as the responsiveness to various ARDS interventions. A period of the first 7 days was chosen to specifically enable a robust comparison with pre-COVID ARDS studies, notably the Berlin definition and LUNG-SAFE studies ^13,19^. Furthermore, by day 14, 51% of the study cohort achieved an outcome of death or survival and hence, examination of early resolution would pick up true signal differences (immediate resolvers may be patients of less concern versus non-resolvers who are patients of more concern). These data show that within the first week, only 25% of patients resolved their hypoxaemia whereas three-quarters, remained static or worsened, despite the increased application of adjunctive ARDS interventions such as PEEP>10cmH_2_O, prone position and inhaled vasodilators in the non-resolving group. This is in stark contrast to previous pre-COVID ARDS studies showed progression of 13.5% ^19^ and 15% ^13^ (see table 2), underlining the severity and different progression, of this disease during the first surge. Non-resolvers were older, had a shorter duration of symptoms prior to ICU admission (suggesting an earlier stage and/or more aggressive course of disease), and a mortality of 59.4% whereas, patients whose hypoxaemia resolved in the first week had a 16.3% mortality. Additionally, resolvers showed improvements in non-respiratory SOFA score and faster reduction in CRP and resolution of lymphopenia. Resolvers underwent prone position on average 2 days earlier than non-resolvers. Higher age and male gender independently predicted non-resolution in the first week of IMV.

Importantly, hypoxaemia could have a significant attributable impact on mortality in Covid-19. In particular, many patients die with (and possibly as a result of) severe refractory hypoxaemia, often refractory to interventions such as high PEEP, conservative fluid balance, NMBA, and prone positioning, potentially explaining the high reported rates of withdrawal of support during a pandemic. It has been suggested that failure to improve oxygenation index (OI) over the first 7 days may provide indications for failure of interventions in clinical trials ^26^. We chose PaO_2_ /FiO_2_ as this is used as clinical criteria for application and termination of interventions. Indeed, there is a strong inverse significant relationship between oxygenation index (OI) and PaO_2_/FiO_2_ in our dataset (supplementary figure S7 - Spearman R=-0.89; P<0.001), suggesting that PaO_2_/FiO_2_ could potentially be replaced by OI in our analysis. Furthermore, the ANOVA shows that OI separates significantly between all groups. Given the retrospective nature of this analysis, prospective validation to assess utility of OI in clinical risk prediction and failure of interventions is important for future studies.

### Prone Positioning

Prone position is a proven evidence-based intervention for ARDS which should be applied to patients suitably fitting criteria ^20^. In particular, the enormous clinical impact of prone position on gas exchange and mortality in pre-COVID ARDS, prompted this evaluation of prone position during Covid-19 to assess how its application could be better understood and improved. Whilst prone positioning improves oxygenation, however, there are conflicting reports as to whether this equates to improved mortality ^27,28^. Indeed, the oxygenation response to prone position is likely a non-linear interaction between improved ventilation perfusion matching, more homogenous distribution of lung stress and lung strain with lower ventilator-induced lung injury, and reduced loading and strain of the right ventricle ^29^. However, in our study, less than half of patients who underwent prone positioning showed a sustained response in PaO_2_/FiO_2_ over the following week which is a key clinical indicator for termination of prone interventions ^30^. Prone responders showed improvements in PaCO_2_, OI, ventilatory ratio (VR), and lower peak pressures. This is consistent with findings from the APRONET study which showed improved oxygenation with reduced driving pressure ^31^. Non-responders had a higher peri-pronation lactate, Ferritin, D-dimer, and worse cardiac indices (i.e. higher cardiovascular SOFA and respiratory SOFA score, troponin, and BNP). We have recently shown in a cohort of 90 patients that right ventricular fractional area change (FAC) and ventricular-pulmonary artery coupling (as measured by FAC:Right ventricular systolic pressure ratio) correlated significantly not only with troponin, BNP and pulmonary vascular resistance but also with measures of ventilation (namely PEEP and PaO_2_/FiO_2_) and a liver marker of congestion (alanine transaminase)^32^. These data suggest the need for closer attention to measures indicative of pulmonary vascular inflammation, thrombosis, and subsequent right heart dysfunction as determinants of which COVID-19 patients fail prone position and, for appropriate escalation to more advanced support such as ECMO ^10,11,33^.

The median PaO_2_/FiO_2_ in non-survivors was 12.3(8.9-18.4) kPa suggesting many patients died with severe hypoxaemia, the attributable mortality of which remains undetermined in Covid-19. The intention of our analysis was to discover characteristics between proned patients who did and did not show a sustained change in oxygenation response to enable earlier risk stratification for patients. Non-resolvers underwent prone positioning 2 days later than resolvers; however, prone responders and non-responders underwent the interventions for a similar length but yet non-responders showed nearly double the mortality of responders. Importantly, responsiveness to prone positioning decayed with duration of IMV prior to the first prone position intervention suggesting that prospective studies should examine whether earlier prone positioning regardless of hypoxaemia severity could beneficially modulate disease progression in Covid-19, especially considering the current application of prone position in self-ventilating patients ^34^. Although the time prone position was applied after intubation was similar between responders and non-responders; non-responders could have received prone positioning earlier as they had greater missed prone windows. This could be due to a higher severity of illness and other factors known to impede application of this important intervention ^31^. Pre-pandemic, ECMO referral criteria in the UK have a critical time window, usually within 7 days of mechanical ventilation, and once a patient is refractory to interventions such as prone position. Understanding personalised responsiveness of prone position in COVID-19 may enable these traditional criteria to be re-evaluated. Of importance half of patients with severe ARDS did not have prone inteventions, suggesting that factors outside the scope of the current dataset (e.g. systems-related or lack of clinical awareness e.g. judgement that hypoxaemia is not severe enough or cardiovascular instability) may need to be assessed in future prospective studies ^31^.

### Strengths and Limitations

There are clear limitations of this analysis, not least its observational, retrospective nature, with testing not standardized across sites, and some sites not being able to complete all data for all patients. For instance, we have not included an analysis between sites and have focussed on the physiology and progression of patients solely undergoing invasive mechanical ventilation for COVID-19. In contrast, to many studies with a focus on all hospitalised patients we chose to focus on patients undergoing mechanical ventilation as this remains a key defining criteria for admission to ICU as well as a decision for active treatment ^16,35^. Further limitations include bias towards ICUs that could contribute data and 38% of patients were managed in specialist severe acute respiratory failure centres often out of necessity as these centres saw a significant number of capacity related transfers (34%). Information censuring because of death or improvement may result in bias in the longitudinal data, but only patients with continuous data for the first 7 days were included in these analyses. Long-term outcome (e.g. 60-/90-day mortality) were not captured as this was not the focus of this analysis. However, the UK Intensive Care National Audit and Research Centre (ICNARC) ^17^ report that most deaths (88.7%) and ICU discharges (79.6%) occurred before day 30 whereas most acute hospital discharges (82%) occurred between days 60 and 90. This is not dissimilar to our findings where 93% of deaths and 72% of discharges occurred at 28-days. Furthermore, this report shows that 52.2% of patients receiving invasive mechanical ventilation survived ICU and 48.2% achieved hospital discharge. Hence, only 4% of patients died in hospital having been discharged from ICU. Patients progressing to ECMO were excluded due to the lack of data prior to ECMO support. Finally, this study evaluated routine clinical care, and hence as such, there may be missing data, as I) some ICUs may not have collected any data at all of a certain measurement modality, in contrast to the majority of the sites (e.g. based on laboratory service configuration); ii) ICUs may have collected the data of a modality but it was not collected for specific patients in contrast to the other patients at that site (i.e. based on clinical need); iii) ICUs may have planned to collect the data of a modality for all patients at their site, but some data points (e.g. on the afternoon of the 3rd day) are missing (i.e. data capture within the clinical information systems); and iv) ICUs may not have transposed the data into the eCRF (i.e. RedCAP data entry was missed). We made a heuristic decision of setting the threshold of missingness to balance of patients we could include against the number of variables (tables S17 and S18 describe missingness by site and over time).

Regarding strengths, we opted for a twice daily collection of data in contrast to a worst daily value, to appreciate the progression of disease and impact of complex interventions, but also achieve a pragmatic balance with ease of data collection for sites. The novelty of this analysis is the utilisation of routinely measured clinical physiological and laboratory parameters over time to determine trajectories and application of interventions. We have described here the most complete COVID critical care dataset published thus far with most data missingness likely due to the variations in care ^22^. Given the granular data there are, of course, many considerations with respect to additional analytics, including Artificial Intelligence based methods ^15,36,37^, that can highlight details and temporal trajectories of patients. However, we chose to describe here the fundamental service evaluation aimed at answering front-line clinically relevant critical care questions manage the ongoing Covid-19 emergency. For this purpose, we used established statistical methods that can be immediately understood by busy front-line colleagues. In turn, to help understand the data better we have therefore focused a lot on the visualisation and provided the corresponding intuitive visualisation tool to visualise the patient journeys in any generic web browser (https://www.coviduk.icu/).

### Implications for clinical service provision

The natural history of COVID-ARDS shows significant admission severity with longer duration of ventilation and refractory hypoxaemia with severe hypoxaemia as a primary cause of death. Whilst admission characteristics correlated with severity and outcome, our data suggests that 75% of patients continued to deteriorate over the first 7 days, resulting in a significantly longer period of mechanical ventilation than non-COVID ARDS. The time series analyses of longitudinal disease trajectory emphasises the importance of routine and frequent assessment of inflammation, thrombosis, and cardiac dysfunction in unpicking lack of clinical response to advanced ARDS management. This was reflected in the fact that while the utility of severity scores such as APACHE II have been questioned ^17^, the use of SOFA score, particularly its cardiovascular and coagulation components, may be useful for prediction of progression. The application of ARDS interventions (high PEEP; NMBA; Prone position; and inhaled nitric oxide) increased with hypoxaemia severity. Regarding prone positioning, less than half of patients maintain a PaO_2_/FiO_2_ above 20kPa after its application, and crucially, its effectiveness decayed over time. Many patients did not receive prone position and had many missed prone windows when it could have been applied either earlier or at all. Usage of antibiotics was over 70% and further evaluation is required to understand whether this use was empirical or microbiologically guided. Of note, renal replacement therapy was applied to over a third of patients with no relation to hypoxaemia severity.

We describe key clinical determinants, hypoxaemia resolution and responsiveness to prone position, which may be more reliable for understanding disease trajectory and prognostication in COVID-19 associated ARDS. These data advocate for the development of randomised controlled trials to develop a COVID-19 specific evidence-base for established ARDS interventions in this phenotypically distinct ARDS population. Particular priorities would be assessment of indications, timing, and efficacy of prone position ventilation, benefit and hazards of “open lung” strategies, and optimisation of fluid management. While this evidence-base is developed, recommendations for the practical clinical management should include pro-active, serial re-evaluation of clinical response to advanced ARDS management strategies.

## Conclusions

Mechanically ventilated patients with COVID-19 have a different natural history and trajectory from descriptions of non-COVID ARDS patients, not predictable from admission physiology. Non-responsiveness to advanced ARDS management is high and associated with hypercoagulation and cardiovascular instability. Variations in clinical practise and subsequent clinical trajectories occur which may benefit from re-evaluation and standardisation of evidence-based practise, as evidenced in this study by data-driven means. Our granular data-driven approach and digital online tool demonstrates how a form of “standing” multi-centre service evaluation could help monitor and inform better clinical practice.

## Data Availability

All data is available by emailing the corresponding authors.

https://www.CovidUK.ICU

## Acknowledgements

The authors are grateful to the patients, families and NHS staff of those affected by COVID-19 for whom the NHS had the privilege of delivering care during the pandemic. The data extracted for this evaluation would not have been possible without the expertise and goodwill of multiple doctors, nurses, other allied health professionals, and data coordinators across the NHS hospital organizations that contributed. We also thank the Intensive Care Society for endorsing this national service evaluation. Collaborators are listed in the supplementary appendix page 3.

## Figure Legends

***Figure S1***. (**A**) Outcome of adjunctive interventions application (bold colours) versus no application (lighter colours): from the bottom we stack discharged and intervened (solid green), deceased and intervened (solid red), discharged and not-intervention (light green), deceased and non-intervention (light red). Percentages are out of the total number of patients (N=633). Most patients will have had multiple interventions, so the bar chart does double count. (**B**) Percentages of patients being in a low or high hypoxaemia severity (PaO_2_/FiO_2_ ratio > or < 20kPa) at the last time point before the intervention (i.e. either morning of the day of the intervention or the evening before).

***Figure S2***. (**A**) Time-series analyses of first week resolution. Each panel presents the time-series of a physiological measure of resolvers (light blue) versus non-resolvers (yellow) over the first 3 weeks of IMV (*P<0.05 interaction with mixed model ANOVA over the first week of IMV, see table S7). The Solid lines are the group medians and the shaded areas are the semi-interquartile range. The number of subjects decay over time as patients die and discharge and the initial and final numbers available for each measure are presented on the graph. A subset of these variables is presented in Figure 4C. (**B**) Correlation matrix between all variables considered for the logistic regression (the final set of parameters was based on less then 40% missingness, see table S2). (**C**) The odds ratio and their 95% confidence interval for Univariate and Multivariate logistic regression models where a higher odds ratio is the increased likelihood for progression of hypoxaemia for each step increase in the admission variable and physiological measures. Continuous variables were discretely sized by a split into quartiles (see supplementary methods for details and table S8 for the full stats). All variables (of the list in table S2) with less than 40% missingness were included in the model. Subjects with more than 20% missing data were removed from the analysis.

***Figure S3***. (**A**) Time-series analyses of prone responsiveness. Each panel presents the time-series of a physiological measure of prone responders (blue) versus non-responders (red) from a day before the first prone manoeuvre to 7 days after (*P<0.05 interaction with mixed model ANOVA over this period, see table S10). The Solid lines are the group medians and the shaded areas are the semi-interquartile range. The number of subjects decay over time as patients die and discharge and the initial and final numbers available for each measure are presented on the graph. A subset of these variables is presented in Figure 5A. (**B**) Correlation matrix between all variables considered for the logistic regression (the final set of parameters was based on less then 40% missingness, see table S2). (**C**) The odds ratio and their 95% confidence interval for Univariate and Multivariate logistic regression models where a higher odds ratio is the increased likelihood of not responding to prone position for each step increase in the admission variable and in the pre-pronation (the last record within 24 hours prior intervention) physiological measures. Continuous variables were discretely sized by a split into quartiles (see supplementary methods for details and table S11 for the full stats for pre- and post-prone respectively). All variables (of the list in table S2) with less than 40% missingness were included in the model. Subjects with more than 20% missing data were removed from the analysis.

***Figure S4***. (**A**) Correlation matrix between all variables considered for the logistic regression (the final set of parameters was based on less then 40% missingness, see table S2). (**B**) The odds ratio and their 95% confidence interval for Univariate and Multivariate logistic regression models where a higher odds ratio is the increased likelihood of not responding to prone position for each step increase in the admission variable and in the post-pronation (the first record in the day after intervention) physiological measures. Continuous variables were discretely sized by a split into quartiles (see supplementary methods for details and table s12 for the full stats for pre- and post-prone respectively). All variables (of the list in table S2) with less than 40% missingness were included in the model. Subjects with more than 20% missing data were removed from the analysis.

***Figure S5***. Time-series analyses of admission severity. Each panel presents the time-series of a physiological measure over the first 3 weeks of IMV, grouped by hypoxaemia on IMV initiation: Mild (PaO_2_/FiO_2_>26), Moderate (PaO_2_/FiO_2_: 26.7-13.3), and severe (PaO_2_/FiO_2_<13.3). (*P<0.05 interaction with mixed model ANOVA over the first week of IMV, see table S16). The Solid lines are the group medians, and the shaded areas are the semi-interquartile range. The number of subjects decay over time as patients die and discharge and the initial and final numbers available for each measure are presented on the graph.

***Figure S6***. Time-series analyses of ICU outcome. (**A**) Each panel presents the time-series of a physiological measure of patients who were discharged (green) versus died (red) over the first 3 weeks of IMV (*P<0.05 interaction with mixed model ANOVA over the first week of IMV, see table S14). The Solid lines are the group medians, and the shaded areas are the semi-interquartile range. The number of subjects decay over time as patients die and discharge and the initial and final numbers available for each measure are presented on the graph. (**B**) Correlation matrix between all variables considered for the logistic regression (the final set of parameters was based on less then 40% missingness, see table S2). (**C**) The odds ratio and their 95% confidence interval for Univariate and Multivariate logistic regression models where a higher odds ratio is the increased likelihood of dying for each step increase in the admission variable and physiological measures. Continuous variables were discretely sized by a split into quartiles (see supplementary methods for details and table S14 for the full stats). All variables (of the list in table S2) with less than 40% missingness were included in the model. Subjects with more than 20% missing data were removed from the analysis.

***Figure S7***. Variations in the reported application of interventions between sites. On the y axis are the percentages of patients who received each intervention in each site. On the bars are the number of the patients who received each intervention in each site over the total number of patients from that site.

***Figure S8***. Oxygenation Index (OI) dependency on P/F ratio. Scatter plot of admission measurements of Oxygenation Index vs PaO_2_/FiO_2_ (in red) shows a strong exponential link. Twice daily measurements of Oxygenation Index vs PaO_2_/FiO_2_ of all patients during the entire ICU stay (black) shows the same strong exponential link.

## Table Legends

Table S1 - Individual site contributions

Table S2 - Variables included in logistic regression models

Table S3 - Distribution of comorbidities with a) severity on admission and b) ICU outcome

Table S4 - Comparison between COVID-ICU and the UK Intensive Care National Audit and Research Centre (ICNARC)

Table S5 - The application, median start date and duration of the first episode of interventions

Table S6 - Clinical and physiological characteristics, outcomes, and interventions according to resolution of hypoxaemia over the first week of invasive mechanical ventilation.

Table S7 - Time series mixed model ANOVA according to resolution of hypoxaemia over the first week of mechanical ventilation.

Table S8 - Uni- and multivariate model analysis of factors associated with progression of hypoxaemia over the first week of invasive mechanical ventilation

Table S9 - Clinical and physiological characteristics, outcomes and interventions according to prone responsiveness.

Table S10 - Time series mixed model ANOVA according to prone responsiveness.

Table S11 - Uni- and multivariate model analysis of pre-pronation factors associated with prone responsiveness.

Table S12 - Uni- and multivariate model analysis of post-pronation factors associated with prone responsiveness.

Table S13- Clinical and physiological characteristics, outcomes and interventions according to ICU outcome.

Table S14 - Time series mixed model ANOVA according to ICU outcome.

Table S15- Uni- and multivariate model analysis of factors associated with ICU mortality.

Table S16 - Time series mixed model ANOVA according to ARDS on admission.

Table S17 – Percentages of missing values for each parameter in each site.

Table S18 – Percentages of missing values for each parameter on each day.

