## Supplementary Appendix for "Natural history, trajectory, and management of mechanically ventilated COVID-19 patients in the United Kingdom"

**Brijesh V Patel*, Shlomi Haar, Rhodri Handslip, Teresa Mei-Ling Lee, Sunil Patel, J. Alex Harston, Feargus Hosking-Jervis, Donna Kelly, Barnaby Sanderson, Barbara Bogatta, Kate Tatham, Ingeborg Welters, Luigi Camporota, Anthony C Gordon, Matthieu Komorowski, David Antcliffe, and John R Prowle, Zudin Puthucheary, A. Aldo Faisal*; on behalf of the United Kingdom COVID-ICU National Service Evaluation.**

***Clinical Science contact –** Dr Brijesh V Patel MD PhD. Clinical Senior Lecturer & Consultant in Intensive Care Medicine. Division of Anaesthetics, Pain Medicine & Intensive Care, Department of Surgery & Cancer, Faculty of Medicine, Imperial College London; Department of Adult Intensive Care, The Royal Brompton and Harefield NHS Foundation Trust, Sydney Street, London SW3 6NP

***Data Science contact –** Professor A. Aldo Faisal PhD. Professor of AI & Neuroscience. Dept. Of Computing & Dept. Of Bioengineering and Behaviour Analytics Lab, Data Science Institute and UKRI Centre for Doctoral Training in AI for Healthcare, Imperial College London, SW7 2AZ London, UK, and MRC London Institute for Medical Sciences, London W12 0NN UK

#### UK COVID-ICU National Service Evaluation Consortium members

***Study Committee:*** Brijesh V Patel, Shlomi Haar, Rhodri Handslip, David Antcliffe, Ingeborg Welters, Luigi Camporota, John Prowle, Zudin Puthucheary, and Aldo Faisal

***Writing Committee:*** Brijesh V Patel, Shlomi Haar, David Antcliffe, Ingeborg Welters, Barnaby Sanderson, Luigi Camporota, John Prowle, Zudin Puthucheary, and Aldo Faisal

***Data Science Team:*** Dr Brijesh Patel, Dr Rhodri Handslip, Feargus Hosking-Jervis, Dr Matthieu Komorowski; Dr Shlomi Haar, Professor Aldo Faisal

***Site Contributions (*Site leads):*** **Aintree University Hospitals NHS Foundation Trust:** Emma Addie; Barbara Borgatta*; Roz Chisholm; Alexandra Crocokft; Anna Gilfedder; Peter Harding; Gladys Madzamba; Namratha Mathai; Maya Patel; Amie Reddy; Leanne Smith; Ian Turner-Bone; Lilian Wajero; Raegan Wignall; Laura Wilding. **Northern health and social care Trust:** (Antrim Area hospital, Causeway hospital) Paul Johnston*; Ciaran Mackle; Nicholas McNaughton; Scott Rafferty. **Guys & St Thomas' NHS Foundation Trust:** Gill Arbane; Luigi Camporota*; Alison Dixon; Barnaby Sanderson*. **Imperial College Heathcare NHS Trust:** David Antcliffe*; Mariana Ferrari Barbosa; Carlos Gomez; Abdalla Kaware, Fady Tarek Khalil, Matthieu Komorowski*, Maie Templeton, Raja Bilal Zafar. **University Hospitals of Leicester NHS Trust:** Mrs Michelle Craner; Mrs Sarah Edwards: Neil Flint; Dr Graziella Isgro; Hakeem Yusuff*. **Liverpool University Hospitals NHS Foundation Trust:** Jaime Fernandez Roman; Oliver Hamilton; Brian Johnson; Emily Johnson; Maria Lopez Martinez; Suleman Mulla; David Shaw; Alicia Waite; Victoria Waugh; Ingeborg Welters*; Karen Williams. **Newcastle upon Tyne Hospitals NHS Foundation Trust:** Benjamin Evans; Lewis Gray; Sam Horne; Natalie Jones; Donna Kelly*; Rebecca Lawrance; Jennifer Partridge; Antonia Snell; Lucy Venyo; Thomas Walker. **Royal Brompton and Harefield NHS Foundation Trust:** Frank Barrett; Sebastian Cabero Alonso; Masoomeh Farazdaghi; Ben Garfield; Clara Hernandez Caballero; Stephane Ledot; Teresa Lee; James Leslie; Helen McGuire; Lorraine O'Connor; Maurizio Passariello; Brijesh Patel*; Sunil Patel; Alex Rosenberg; Jennifer Thomson; Dhulamini Wijeratne. **Royal Cornwall Hospitals NHS Trust:** Sarah Bean; Karen Burt; Rosanna Jury; Oliver Quick; Michael Spivey*; Josh Thorns. **Barts Health NHS Trust:** Sana Ali; Thomas Beazer; Matilda Boa; Rosemarie Burd; Zoe Burdon; J Burgess; Natalie Cameron; A Daou; Charles Fadipe; FAJ Freer; Gayle Fung; Lindsey Iles; Aaron Jones; Agne Jovaisaite; Gabriella Knight; Veda Kudva; J Lampard; Ruby Lightfoot; Nicholas Low; Caroline Olabisi; Osama Omrani; Rebecca Pease; John Prowle*; Zudin Puthucheary*; Johnathan Mak; Aamir Saiyed; Maria Salamanca; Gauri Saxena; W Stanley; Nida Tahir; Yize Wan; Anton Weatherhead. **Royal Papworth Hospital NHS Foundation Trust:** Linzi Dunham; Melissa Earwaker*; Amy Gladwell. **Royal Marsden Hospital NHS Foundation Trust:** Thomas Bemand; Ethel Black; Samantha Coulson; Arnold Dela Rosa; Shamen Jhanji; Ravishankar Rao Baikady; Kate Tatham; Zoszka Webb*. **South Tyneside and Sunderland NHS Foundation Trust:** Sarah Cornell; Anthony Rostron*; Fiona Wakinshaw; Lindsey Woods.

#### STROBE statement

| | Item No | Recommendation | Page No |
|---|---|---|---|
| **Title and abstract** | 1 | (*a*) Indicate the study's design with a commonly used term in the title or the abstract | 1-4 |
| | | (*b*) Provide in the abstract an informative and balanced summary of what was done and what was found | 4 |
| **Introduction** | | | |
| Background/rationale | 2 | Explain the scientific background and rationale for the investigation being reported | 5-7 |
| Objectives | 3 | State specific objectives, including any prespecified hypotheses | 7 |
| **Methods** | | | |
| Study design | 4 | Present key elements of study design early in the paper | 8-10 |
| Setting | 5 | Describe the setting, locations, and relevant dates, including periods of recruitment, exposure, follow-up, and data collection | 8 |
| Participants | 6 | (*a*) Give the eligibility criteria, and the sources and methods of selection of participants. Describe methods of follow-up | 8 |
| | | (*b*) For matched studies, give matching criteria and number of exposed and unexposed | NA |
| Variables | 7 | Clearly define all outcomes, exposures, predictors, potential confounders, and effect modifiers. Give diagnostic criteria, if applicable | 8-10 |
| Data sources/ measurement | 8* | For each variable of interest, give sources of data and details of methods of assessment (measurement). Describe comparability of assessment methods if there is more than one group | 8-10 |
| Bias | 9 | Describe any efforts to address potential sources of bias | 8-10 |
| Study size | 10 | Explain how the study size was arrived at | 8 |

| | | | |
|---|---|---|---|
| Quantitative variables | 11 | Explain how quantitative variables were handled in the analyses. If applicable, describe which groupings were chosen and why | 8-10 |
| Statistical methods | 12 | (*a*) Describe all statistical methods, including those used to control for confounding | 8-10 |
| | | (*b*) Describe any methods used to examine subgroups and interactions | 8-10 |
| | | (*c*) Explain how missing data were addressed | 8 |
| | | (*d*) If applicable, explain how loss to follow-up was addressed | NA |
| | | (*e*) Describe any sensitivity analyses | 8-10 |
| **Results** | | | |
| Participants | 13* | (a) Report numbers of individuals at each stage of study—e.g., numbers potentially eligible, examined for eligibility, confirmed eligible, included in the study, completing follow-up, and analysed | 11 |
| | | (b) Give reasons for non-participation at each stage | 11 |
| | | (c) Consider use of a flow diagram | Fig1 |
| Descriptive data | 14* | (a) Give characteristics of study participants (eg demographic, clinical, social) and information on exposures and potential confounders | 11 |
| | | (b) Indicate number of participants with missing data for each variable of interest | NA |
| | | (c) Summarise follow-up time (eg, average and total amount) | 11-14 |
| Outcome data | 15* | Report numbers of outcome events or summary measures over time | 11-14 |

| | | | |
|---|---|---|---|
| Main results | 16 | (*a*) Give unadjusted estimates and, if applicable, confounder-adjusted estimates and their precision (eg, 95% confidence interval). Make clear which confounders were adjusted for and why they were included | 11-14 |
| | | (*b*) Report category boundaries when continuous variables were categorized | NA |
| | | (*c*) If relevant, consider translating estimates of relative risk into absolute risk for a meaningful time period | NA |
| Other analyses | 17 | Report other analyses done—e.g. analyses of subgroups and interactions, and sensitivity analyses | 11-14 |
| **Discussion** | | | |
| Key results | 18 | Summarise key results with reference to study objectives | 15-18 |
| Limitations | 19 | Discuss limitations of the study, taking into account sources of potential bias or imprecision. Discuss both direction and magnitude of any potential bias | 18-19 |
| Interpretation | 20 | Give a cautious overall interpretation of results considering objectives, limitations, multiplicity of analyses, results from similar studies, and other relevant evidence | 20-21 |
| Generalisability | 21 | Discuss the generalisability (external validity) of the study results | 20-21 |
| **Other information** | | | |
| Funding | 22 | Give the source of funding and the role of the funders for the present study and, if applicable, for the original study on which the present article is based | 3 |

*Give information separately for exposed and unexposed groups.

**Note:** An Explanation and Elaboration article discusses each checklist item and gives methodological background and published examples of transparent reporting. The STROBE checklist is best used in conjunction with this article (freely available on the Web sites of PLoS Medicine at http://www.plosmedicine.org/, Annals of Internal Medicine at http://www.annals.org/, and Epidemiology at http://www.epidem.com/). Information on the STROBE Initiative is available at http://www.strobe-statement.org.

#### Detailed Methods

***Study design:*** We performed a multicentre, observational cohort study in patients with SARS-CoV-2 infection who required mechanical ventilation for severe Covid-19 infection in the United Kingdom.

***Exposure:*** Adult patients (aged ≥18 years) with laboratory confirmed SARS-CoV-2 infection who required mechanical ventilation in the United Kingdom between March 1st and August 31st 2020.

***Ethical approval:*** Each site registered the protocol as a service evaluation, as approved by the United Kingdom's Health Research Authority. All patients lacked capacity, and the need for individual informed consent was waived for collection of data during routine care, with no breach of privacy or anonymity. The "Strengthening the Reporting of Observational Studies in Epidemiology" statement guidelines were applied (see supplementary appendix pages 4-5) [1].

***Data collection and procedures:***

We set up a data processing pipeline where only routine, anonymised data was collected with no change to clinical care. Patients were identified through daily review of paper or electronic medical records using a standardised case record form (CRF), with retrospective and prospective data collection permitted. Data was extracted from either electronic healthcare records (EHRs) or paper-based records. Sites were given the option to submit their EHRs either in a predetermined format set by the Covid-ICU own secure REDCap database form fields or as a raw dump of data in CSV file format, that was then record-by-record manually screened for data consistency. Paper-based ICU operations sites were able to enter the data directly by the site representative into our RedCAP Electronic Data Capture (RedCAP v10.0.10; Vanderbilt University, US, local version hosted by Imperial College servers in the UK).

Each site's representative was given a unique username and password for data entering-only access to the database to connect via an encrypted online connection. Confidentiality was protected through a de-identified study number. A range of ICUs were included including secondary and tertiary care organisations (Table S1). In brief, the CRF captured admission demographics, twice daily (8am and 8pm) respiratory physiology and blood gas results, daily ARDS interventions, daily Covid interventions, daily blood results and outcome status. Patients were categorized within 48 hours of invasive mechanical ventilation (IMV) based on their $PaO_2/FIO_2$ ratio into no ARDS, mild, moderate, and severe ARDS [2].

**Missing data and imputation:** We made the heuristic decision of setting the threshold of data completeness (i.e. missingness) to balance of patients we could include against the number of variables. We defined this by examination of the available variables in the first 48 hours of admission or the last 36 hours before prone or the first 36 hours after prone. If in these 3/4 twelve-hour measurement points, all were missing, then we counted this patient as 'missing' data. The missingness is thus the percentage of patients where there is no measurement in this 36/48-hour window for a modality. Percentage of missing data per modality are shown in Table S2, and details of missing data are shown in Table S18. Data imputation was applied using k-nearest neighbours' algorithm. We ran the imputation with a k of 3, 5, and 7 both on the continuous variable and on the quartile categorization. The maximal odds ratio difference between the imputation approaches for each variable was 0.04 (IQR 0.03-0.07) and had no effect on the significance. All reported results are based on 5-nearest neighbours' imputation on the quartile categorization.

**Resolution of hypoxaemia over first week of IMV.** Hypoxaemia was categorised as per Berlin definition of ARDS [2]. First week resolvers were defined by moving over the first week of mechanical ventilation to a less severe ARDS hypoxaemia category, and vice-versa for non-resolvers [2,3]. In addition to the patients who resolved, also those who remained mild or got discharged were considered "resolvers" while those who deteriorate, remained moderate or severe, or died, were considered "non-resolvers".

**Responsiveness to prone position.** We considered the longer-term effect on $PaO_2/FiO_2$ after prone positioning and defined prone responsiveness as maintenance of a mean $PaO_2/FiO_2$ >20kPa over 7 days after the first prone episode. Finally, we defined a proning window as a $PaO_2/FiO_2$<20kPa, with an $FiO_2$≥0.6, a PEEP≥5cm$H_2O$ to assess opportunities to apply the intervention. Prone windows were measured at 8am and 8pm with the ventilator and arterial blood gas evaluation.

**Interventions periods.** The incidence and duration of interventions as well as ventilation setting were analysed and reported to current strategies e.g. low tidal volume ventilation and ARDSNet PEEP tables. We defined an intervention period as a daily application of the intervention with a day of no intervention defining the end of the current period and the onset of the next period.

**Statistical analysis.** Descriptive variables are expressed as percentage, or median and interquartile range (IQR), as appropriate. The distribution of each variable on each day in each outcome group was tested for normality using the Shapiro-Wilks test, and where necessary and appropriate, because of skewed distributions, continuous variables were log-transformed (e.g. $FiO_2$). Continuous variables were analysed with Mann Whitney U or Kruskall Wallis tests, as appropriate. Categorical variables were compared using Fisher's exact test or the Chi-square test for equal proportion, as appropriate. All statistical tests were 2-sided and $p \leq 0.05$ was considered statistically significant. For group-wise analysis, the outcome of the therapies was measured as categorical variables of “Mild or Moderate or Severe”, “Survival or Death”, “resolver or non-resolver”, and “prone responder or prone non-responder”. Hypoxaemia was categorised as per Berlin definition of ARDS [2]. First week resolvers were defined by moving over the first week of mechanical ventilation to a less severe ARDS hypoxaemia category, and vice-versa for non-resolvers [2]. We defined a positive prone responsiveness as a sustained increase in mean $PaO_2/FiO_2$ above 20kPa over 7 days, following the first PP episode. Analyses were carried out using Matlab (Mathworks Inc., Natick, MA).

**Logistic regression models.** Multivariate logistic regression models were applied (with screening univariate, p<0.1) to each outcome variable to test associations with independent variables. The full list of variables tested for inclusion in these models is shown in Table S2, but only variables with less than 40% missingness were included in each outcome model (missing value analysis in the relevant time points is shown in Table S2). Variables that showed clinical overlap (e.g. SOFA renal and creatinine) had one variable excluded. A data driven approach to collinearity was not taken as many clinical variables associated with each other due to relationships with severity of illness. A full correlation matrix can be found in the supplementary figures. For all outcomes, only patients with more than 80% of the variables were included in the models. Accordingly, up to 20% of the data were missing and thus were imputed. Data were assumed to be missing at random (MAR) owing to the nature of different personnel at many different sites completing each data entry (the full missing value analysis, by site and by day, is shown in Tables S17 and S18). To enable interpretable and comparable odds ratios, all continuous variables were transformed to categorical by splitting them into quartiles. Accordingly, the odds ratio is the risk increase per quartile increase in the measurement. For age, the odds ratio is the risk increase per decade increase; for SOFA scores, the odds ratio is the risk increase per unit increase in the SOFA score; and for binary variables (e.g., gender, comorbidities) the odds ratio is the risk increase of being positive (e.g. being male, having comorbidity).

**Statistical analysis of natural history and management.** The association between the change over time of each independent variable and the outcome measures was tested in repeated measures (rm) ANOVA. For the survival and first week resolver outcome, rmANOVA was applied on the physiology variables over the first week of mechanical ventilation, while for the prone responder outcome, it was applied on the physiology variables over a week from the day before the first PP episode. The rmANOVA was applied separately to each physiology variable, and for each variable, only patients with more than 80% of the variable's measurements over that week were included in the model. Variables for which fewer than 30 patients had more than 80% of the measurements were not analysed. To prevent the risk of too many false positive, we accounted for multiple comparisons in the interaction statistic by controlling the false discovery rate (FDR).

**References.**

1 Elm E von, Altman DG, Egger M, et al. The Strengthening the Reporting of Observational Studies in Epidemiology (STROBE) Statement: guidelines for reporting observational studies*. B World Health Organ 2007; 85: 867–72.

2 Acute Respiratory Distress Syndrome: The Berlin Definition. Jama 2012; 307: 2526–33.

3 Bellani G, Laffey JG, Pham T, et al. Epidemiology, Patterns of Care, and Mortality for Patients With Acute Respiratory Distress Syndrome in Intensive Care Units in 50 Countries. Jama 2016; 315: 788–800.

#### Table S1 - Individual site contributions

| ICU site | Number of patients | Proportion of total | Outcome (Mortality rate) | percentage transferred in |
|---|---|---|---|---|
| A | 2 | 0.3% | 100.0% | 0.0% |
| B | 1 | 0.2% | 0.0% | 100.0% |
| C | 18 | 2.8% | 27.8% | 5.6% |
| D | 35 | 5.5% | 31.4% | 82.9% |
| E | 49 | 7.7% | 38.8% | 79.6% |
| F | 19 | 3.0% | 26.3% | 15.8% |
| G | 36 | 5.7% | 47.2% | 44.4% |
| H | 57 | 9.0% | 59.6% | 7.0% |
| I | 34 | 5.4% | 64.7% | 0.0% |
| J | 34 | 5.4% | 29.4% | 76.5% |
| K | 3 | 0.5% | 66.7% | 0.0% |
| L | 64 | 10.1% | 51.6% | 4.7% |
| M | 54 | 8.5% | 48.1% | 3.7% |
| N | 5 | 0.8% | 80.0% | 40.0% |
| O | 9 | 1.4% | 33.3% | 77.8% |
| P | 31 | 4.9% | 35.5% | 16.1% |
| Q | 9 | 1.4% | 88.9% | 0.0% |
| R | 173 | 27.3% | 32.4% | 12.1% |
| **Total** | **633** | | | |

#### Table S2 - Variables included in logistic regression models

| | % of Missingness | | |
|---|---|---|---|
| **FieldLabel** | **Admission** | **Before PP** | **After PP** |
| Age | 0% | 0% | 0% |
| Male | 0% | 0% | 0% |
| BMI | 17% | 11% | 11% |
| Height | 23% | 16% | 16% |
| symptoms days | 36% | 33% | 33% |
| Hypertension | 0% | 0% | 0% |
| Diabetes mellitus | 0% | 0% | 0% |
| Oxygen saturation | 18% | 14% | 10% |
| pH | 0% | 6% | 0% |
| PaCO_2 | 0% | 6% | 0% |
| HCO^-3 | 1% | 6% | 0% |
| Lactate | 5% | 10% | 6% |
| Peak pressure | 5% | 13% | 5% |
| PEEP | 5% | 14% | 7% |
| Minute ventilation | 4% | 14% | 6% |
| Dynamic Comp | 11% | 21% | 13% |
| Oxygenation Index | 39% | 50% | 46% |
| Ventilatory Ratio | 26% | 27% | 20% |
| Cum fluid balance | 7% | 14% | 19% |
| Glucose | 8% | 11% | 17% |
| BUN | 21% | 31% | 33% |
| Sodium | 2% | 4% | 2% |
| Potassium | 2% | 4% | 3% |
| ALP | 5% | 6% | 9% |
| ALT | 6% | 9% | 13% |
| Creatinine Kinase | 62% | 56% | 61% |
| LDH | 69% | 63% | 67% |
| Haemoglobin | 2% | 4% | 2% |
| Haematocrit | 51% | 37% | 35% |
| Neutrophils | 2% | 4% | 2% |
| Monocytes | 3% | 4% | 3% |
| Lymphocytes | 3% | 4% | 3% |
| Basophils | 21% | 14% | 16% |
| Eosinophils | 22% | 15% | 14% |
| APTT | 37% | 31% | 37% |
| PT | 37% | 31% | 36% |
| Fibrinogen | 35% | 33% | 44% |
| Ferritin | 54% | 42% | 51% |
| D-dimer | 38% | 40% | 44% |
| Triglicerides | 70% | 69% | 77% |
| CRP | 6% | 10% | 11% |
| Procalcitonin | 80% | 77% | 82% |
| High sensitivity Troponin | 60% | 56% | 64% |
| NT Pro BNP | 91% | 93% | 94% |
| SOFA Respiratory | 1% | 6% | 0% |
| SOFA Nervous | 20% | 33% | 27% |
| SOFA Cardio | 9% | 11% | 9% |
| SOFA Liver | 7% | 15% | 13% |
| SOFA Coagulation | 2% | 4% | 2% |
| SOFA Kidneys | 1% | 3% | 0% |
| SOFA score | 32% | 49% | 41% |
| Prone initiation day | - | 0% | 0% |

### Table S3 - Distribution of co-morbidities with a) severity on admission and b) ICU outcome

| Co-morbidity according to severity | ALL | | | MILD | | | MODERATE | | | SEVERE | | | P value |
|---|---|---|---|---|---|---|---|---|---|---|---|---|---|
| | Total N | n | % | Group N | n | % | Group N | n | % | Group N | n | % | |
| BMI>30 kg/m2 | 524 | 209 | 39.9 | 123 | 44 | 35.77 | 278 | 107 | 38.5 | 123 | 58 | 47.2 | 0.626 |
| Myocardial infarction | 573 | 17 | 3.0 | 129 | 6 | 4.65 | 292 | 7 | 2.4 | 152 | 4 | 2.6 | 0.485 |
| Congestive Heart Failure | 573 | 11 | 1.9 | 128 | 2 | 1.56 | 293 | 6 | 2.0 | 152 | 3 | 2.0 | 0.922 |
| Peripheral vascular disease | 574 | 9 | 1.6 | 129 | 1 | 0.78 | 293 | 6 | 2.0 | 152 | 2 | 1.3 | 0.577 |
| CVA or TIA | 574 | 17 | 3.0 | 129 | 4 | 3.10 | 293 | 9 | 3.1 | 152 | 4 | 2.6 | 0.966 |
| Dementia | 574 | 2 | 0.3 | 129 | 1 | 0.78 | 293 | 1 | 0.3 | 152 | 0 | 0.0 | 0.564 |
| Hypertension | 574 | 242 | 42.2 | 129 | 51 | 39.53 | 293 | 132 | 45.1 | 152 | 59 | 38.8 | 0.283 |
| Pregnancy | 366 | 1 | 0.3 | 71 | 0 | 0.00 | 183 | 1 | 0.5 | 112 | 0 | 0.0 | 0.613 |
| COPD | 575 | 41 | 7.1 | 129 | 10 | 7.75 | 294 | 24 | 8.2 | 152 | 7 | 4.6 | 0.372 |
| Connective tissue disease | 573 | 5 | 0.9 | 129 | 1 | 0.78 | 292 | 4 | 1.4 | 152 | 0 | 0.0 | 0.331 |
| Peptic ulcer disease | 573 | 8 | 1.4 | 129 | 1 | 0.78 | 293 | 5 | 1.7 | 151 | 2 | 1.3 | 0.728 |
| Diabetes mellitus | 574 | 176 | 30.7 | 129 | 46 | 35.66 | 293 | 93 | 31.7 | 152 | 37 | 24.3 | 0.161 |
| Diabetes mellitus - end organ damage | 569 | 18 | 3.2 | 127 | 5 | 3.94 | 291 | 10 | 3.4 | 151 | 3 | 2.0 | 0.637 |
| Hemiplegia | 365 | 2 | 0.5 | 70 | 1 | 1.43 | 183 | 1 | 0.5 | 112 | 0 | 0.0 | 0.564 |
| Moderate to severe CKD | 366 | 20 | 5.5 | 71 | 4 | 5.63 | 183 | 13 | 7.1 | 112 | 3 | 2.7 | 0.380 |
| Solid tumour | 575 | 24 | 4.2 | 129 | 7 | 5.43 | 294 | 10 | 3.4 | 152 | 7 | 4.6 | 0.653 |
| Leukemia | 366 | 8 | 2.2 | 71 | 1 | 1.41 | 183 | 7 | 3.8 | 112 | 0 | 0.0 | 0.095 |
| Lymphoma | 365 | 5 | 1.4 | 71 | 1 | 1.41 | 182 | 4 | 2.2 | 112 | 0 | 0.0 | 0.331 |
| AIDS | 574 | 5 | 0.9 | 129 | 3 | 2.33 | 293 | 2 | 0.7 | 152 | 0 | 0.0 | 0.113 |
| Steroid use | 365 | 14 | 3.8 | 71 | 3 | 4.23 | 182 | 10 | 5.5 | 112 | 1 | 0.9 | 0.198 |
| Other Immunosupression use | 366 | 16 | 4.4 | 71 | 4 | 5.63 | 183 | 11 | 6.0 | 112 | 1 | 0.9 | 0.166 |
| History of Deep Vein Thrombosis | 363 | 7 | 1.9 | 70 | 4 | 5.71 | 181 | 2 | 1.1 | 112 | 1 | 0.9 | 0.102 |
| History of Pulmonary Embolus | 364 | 5 | 1.4 | 71 | 1 | 1.41 | 181 | 3 | 1.7 | 112 | 1 | 0.9 | 0.911 |

| Co-morbidity according to outcome | Survivors | | | Non-Survivors | | | P value |
|---|---|---|---|---|---|---|---|
| | Group N | n | % | Group N | n | % | |
| BMI>30 kg/m2 | 314 | 134 | 42.68 | 210 | 75 | 35.71 | 0.021 |
| Myocardial infarction | 326 | 4 | 1.23 | 247 | 13 | 5.26 | 0.004 |
| Congestive Heart Failure | 326 | 2 | 0.61 | 247 | 9 | 3.64 | 0.008 |
| Peripheral vascular disease | 326 | 5 | 1.53 | 248 | 4 | 1.61 | 0.898 |
| CVA or TIA | 326 | 10 | 3.07 | 248 | 7 | 2.82 | 0.922 |
| Dementia | 326 | 1 | 0.31 | 248 | 1 | 0.40 | 0.826 |
| Hypertension | 326 | 129 | 39.57 | 248 | 113 | 45.56 | 0.081 |
| Pregnancy | 185 | 0 | 0.00 | 181 | 1 | 0.55 | 0.243 |
| COPD | 327 | 19 | 5.81 | 248 | 22 | 8.87 | 0.129 |
| Connective tissue disease | 326 | 1 | 0.31 | 247 | 4 | 1.62 | 0.087 |
| Peptic ulcer disease | 326 | 3 | 0.92 | 247 | 5 | 2.02 | 0.245 |
| Diabetes mellitus | 326 | 95 | 29.14 | 248 | 81 | 32.66 | 0.244 |
| Diabetes mellitus - end organ damage | 323 | 8 | 2.48 | 246 | 10 | 4.07 | 0.250 |
| Hemiplegia | 185 | 1 | 0.54 | 180 | 1 | 0.56 | 0.826 |
| Moderate to severe CKD | 185 | 8 | 4.32 | 181 | 12 | 6.63 | 0.104 |
| Solid tumour | 326 | 12 | 3.68 | 249 | 12 | 4.82 | 0.439 |
| Leukemia | 185 | 2 | 1.08 | 181 | 6 | 3.31 | 0.060 |
| Lymphoma | 185 | 1 | 0.54 | 180 | 4 | 2.22 | 0.087 |
| AIDS | 326 | 4 | 1.23 | 248 | 1 | 0.40 | 0.310 |
| Steroid use | 185 | 8 | 4.32 | 180 | 6 | 3.33 | 0.968 |
| Other Immunosupression use | 185 | 7 | 3.78 | 181 | 9 | 4.97 | 0.254 |
| History of Deep Vein Thrombosis | 184 | 2 | 1.09 | 179 | 5 | 2.79 | 0.117 |
| History of Pulmonary Embolus | 184 | 2 | 1.09 | 180 | 3 | 1.67 | 0.422 |

#### Table S4 - Comparison between Covid-ICU and ICNARC

| | Covid-19 undergoing IMV | |
|---|---|---|
| | **COVID-ICU** | **ICNARC*** |
| | **N=633** | **N=7,702** |
| **Patient characteristics** | | |
| Age (years), median (IQR) | 59.0 [51.0 66.0] | 60 (51-67) |
| Sex, n (%) | | |
| Male | 481 (76%) | 5,544 (72.0%) |
| Ethnicity, n (%) | | |
| White | 250 (39.5%) | 4,653 (60.4%) |
| Asian | 73 (11.5%) | 1,256 (16.3%) |
| Black | 115 (18.2%) | 804 (10.4%) |
| Other | 103 (16.3%) | 693 (9.0%) |
| Not stated | 91 (14.4%) | 296 (3.8%) |
| BMI (kg/m2), median (IQR) | 28.1 [24.9 32.8] | 28.3 (24.9-33.0) |
| BMI categories (kg/m$^2$), n (%) | | |
| >30 | 209 (39.9%) | 39.9% |
| **Acute illness severity**[4] | | |
| P/F Ratio (kPa),[5] median (IQR) | 18.3 [13.0 25.0] | 15.0 (10.8-21.0) |
| P/F ratio categories (kPa)[5] | | |
| ≤13.3kPa (≤100mmHg) | 166 (26.2%) | 3,033 (40.4%) |
| 13.3-26.7kPa (100-200mmHg) | 320 (50.6%) | 3,530 (47.0%) |
| >26.7kPa (>200mmHg) | 147 (23.2%) | 952 (12.7%) |
| **Organ support**[8,9] | | |
| *Renal support* | | |
| Receipt, n (%) | 211 (33.3%) | 2,708 (35.2%) |
| Duration (calendar days), median (IQR) | 8.0 [4.0 14.0] | 8 (4-15) |
| **Outcome** | | |
| *Critical care* | | |
| Survived, n (%) | 365 (57.7%) | 4,018 (52.2%) |
| Died, n (%) | 268 (42.3%) | 3,684 (47.8%) |
| **Duration of stay (calendar days)** | | |
| *IMV*, median (IQR) | 13.0 [7.0 22.0] | 13 (7–23) |
| Survived – median (IQR) | 15.0 [8.0 28.0] | - |
| Died – median (IQR) | 11.0 [6.0 16.0] | - |
| Critical care, median (IQR) | 14.0 [8.0 23.0] | 15.0 (8.0-27.0) |
| Survived – median (IQR) | 17.0 [9.8 30.0] | 22.0 (12.0-36.0) |
| Died – median (IQR) | 11.0 [7.0 18.0] | 10.0 (6.0-17.0) |

* Richards-Belle, A., Orzechowska, I., Gould, D., Thomas, K., Doidge, J., Mouncey, P., Christian, M., Shankar-Hari, M., Harrison, D., Rowan, K., Banjo, Y., Borowczak, K., Cousins, T., Cummins, P., Dalemo, K., Darnell, R., Demissie, H., Drikite, L., Fleming, A., Frederiksen, D., Furnell, S., Hussein, A., Koelewyn, A., Matthews, T., Peters, S., Samuels, T., Saull, M. (2020). COVID-19 in critical care: epidemiology of the first epidemic wave across England, Wales and Northern Ireland Intensive Care Medicine https://dx.doi.org/10.1007/s00134-

#### Table S5 - The application, median start date and duration of the first episode of interventions

| | Start day of first intervention | Duration of first intervention | Number of periods |
|---|---|---|---|
| **interventions** | median [IQR] | median [IQR] | median [IQR] |
| Neuro-muscular blockade | 1.0 [0.0 3.0] | 4.0 [1.0 7.0] | 1.0 [1.0 2.0] |
| Prone positioning | 2.0 [0.8 5.0] | 2.0 [1.0 4.0] | 1.0 [1.0 2.0] |
| Inhaled nitric oxide | 6.0 [3.0 9.0] | 4.0 [2.0 7.3] | 1.0 [1.0 1.0] |
| Inhaled prostacyclin | 7.0 [3.0 15.0] | 3.0 [1.0 6.8] | 1.0 [1.0 1.0] |
| Tracheostomy | 14.0 [9.0 18.0] | 13.0 [6.0 20.0] | 1.0 [1.0 1.0] |
| APRV | 3.0 [0.5 6.0] | 3.0 [2.0 5.0] | 1.0 [1.0 1.0] |
| Bronchoscopy | 9.0 [3.0 15.8] | 1.0 [1.0 1.0] | 1.0 [1.0 1.0] |
| Renal replacement therapy | 3.0 [1.0 6.0] | 5.0 [3.0 11.0] | 1.0 [1.0 2.0] |
| Diuretics | 1.0 [1.0 3.0] | 3.0 [1.0 5.0] | 2.0 [1.0 3.0] |
| Corticosteroids | 5.0 [1.0 10.0] | 4.0 [2.0 9.0] | 1.0 [1.0 2.0] |
| Therapeutic heparin | 9.0 [6.0 14.0] | 5.0 [2.0 8.8] | 1.0 [1.0 1.0] |
| Anti-Bacterial | 0.0 [0.0 0.0] | 6.0 [4.0 9.0] | 1.0 [1.0 2.0] |

### Table S6 - Clinical and physiological characteristics, outcomes and interventions according to resolution of hypoxaemia over first week of invasive mechanical ventilation.

| Clinical Characterisitcs | | | | | Trajectory between admission and day7 | | | | |
|---|---|---|---|---|---|---|---|---|---|
| | | | ALL | | Resolving | | Non resolving | | |
| | label | units | Total N | median [IQR] / N (%) | Group N | median [IQR] / N (%) | Group N | median [IQR] / N (%) | P Value |
| | Male | | 633 | 481 (76%) | 267 | 196 (73.4%) | 366 | 285 (77.9%) | 0.194 |
| | White | | 542 | 250 (46.1%) | 216 | 98 (45.4%) | 326 | 152 (46.6%) | 0.220 |
| | Age | years | 633 | 59.0 [51.0 66.0] | 267 | 57.0 [47.0 64.0] | 366 | 60.0 [54.0 67.0] | 0.000 |
| | BMI | kg/m2 | 524 | 28.1 [24.9 32.8] | 222 | 28.0 [24.8 32.9] | 302 | 28.2 [25.1 32.3] | 0.666 |
| | Time since onset of symptoms | days | 408 | 8.0 [6.0 12.0] | 164 | 9.0 [6.5 13.5] | 244 | 7.0 [6.0 11.0] | 0.004 |
| | ICU length of stay | days | 633 | 14.0 [8.0 23.0] | 267 | 13.0 [7.3 20.0] | 366 | 15.0 [8.0 25.0] | 0.025 |
| | Length of mechanical ventilation | days | 633 | 13.0 [7.0 22.0] | 267 | 12.0 [6.0 18.8] | 366 | 14.0 [8.0 24.0] | 0.003 |
| | ICU Mortality | % | 633 | 268 (42.3%) | 267 | 47 (17.6%) | 366 | 221 (60.4%) | 0.000 |
| Vent | $FiO_2$ (%) | | 628 | 60.0 [45.0 80.0] | 263 | 60.0 [40.0 80.0] | 365 | 60.0 [50.0 72.8] | 0.269 |
| | $PaO_2$ to $FiO_2$ ratio | | 626 | 18.3 [13.0 25.0] | 262 | 17.9 [11.7 27.5] | 364 | 18.4 [14.6 24.1] | 0.323 |
| | Tidal Volume (mls/IBW) | ml/Kg(IBW) | 472 | 6.8 [6.0 7.8] | 202 | 6.9 [6.1 7.8] | 270 | 6.8 [6.0 7.7] | 0.373 |
| | Respiratory rate | bpm | 627 | 18.8 [16.0 22.0] | 263 | 18.0 [16.0 22.0] | 364 | 19.2 [16.0 22.4] | 0.249 |
| | Minute ventilation | L/minute | 606 | 8.5 [6.9 10.4] | 255 | 8.5 [7.0 10.2] | 351 | 8.5 [6.9 10.5] | 0.500 |
| | Peak pressure | ml/Kg(RBW) | 599 | 26.0 [23.0 30.0] | 253 | 25.0 [22.0 29.0] | 346 | 26.7 [24.0 30.0] | 0.005 |
| | Plateau pressure | ml/Kg(IBW) | 80 | 26.0 [22.5 28.5] | 28 | 24.5 [21.5 29.0] | 52 | 26.0 [23.0 28.0] | 0.438 |
| | PEEP | cmH_20 | 603 | 10.0 [8.0 12.0] | 257 | 10.0 [8.0 12.0] | 346 | 10.0 [8.5 12.0] | 0.008 |
| | Mean airway pressure | cmH_20 | 387 | 16.0 [13.2 19.0] | 174 | 15.8 [13.0 18.0] | 213 | 16.0 [14.0 20.0] | 0.037 |
| | Pressure support | cmH_20 | 371 | 10.0 [5.3 14.0] | 160 | 10.0 [5.0 12.5] | 211 | 10.0 [6.3 14.0] | 0.651 |
| | Dynamic Compliance | mls/cmH20 | 564 | 31.5 [24.3 40.2] | 242 | 32.5 [25.2 40.9] | 322 | 30.3 [24.0 40.1] | 0.253 |
| | Oxygenation Index | | 387 | 8.1 [5.1 12.5] | 174 | 7.7 [4.6 13.0] | 213 | 8.2 [5.3 11.6] | 0.600 |
| | Ventilatory Ratio | | 470 | 1.5 [1.2 2.1] | 200 | 1.4 [1.2 2.0] | 270 | 1.6 [1.3 2.1] | 0.250 |
| ABG | Oxygen saturation | | 518 | 95.0 [93.0 98.0] | 224 | 96.0 [93.0 98.0] | 294 | 95.0 [92.0 97.0] | 0.060 |
| | pH | | 630 | 7.4 [7.3 7.4] | 265 | 7.4 [7.3 7.4] | 365 | 7.4 [7.3 7.4] | 0.058 |
| | $PaO_2$ | kPa | 630 | 10.7 [9.2 13.1] | 265 | 10.4 [8.8 12.4] | 365 | 10.9 [9.4 13.4] | 0.019 |
| | $PaCO_2$ | kPa | 630 | 6.0 [5.2 7.2] | 265 | 6.0 [5.2 7.2] | 365 | 6.0 [5.2 7.1] | 0.959 |
| | Base excess | | 630 | -0.3 [-2.6 2.2] | 265 | 0.3 [-1.9 2.5] | 365 | -0.7 [-3.0 1.8] | 0.001 |
| | $HCO_3$ | mmol/L | 629 | 24.5 [22.5 26.7] | 265 | 24.9 [23.0 27.2] | 364 | 24.0 [22.0 26.4] | 0.001 |
| | Lactate | mmol/L | 604 | 1.2 [1.0 1.6] | 260 | 1.2 [1.0 1.6] | 344 | 1.3 [1.0 1.6] | 0.045 |
| SOFA | SOFA score | | 428 | 9.0 [7.0 11.0] | 184 | 9.0 [7.0 11.0] | 244 | 9.0 [7.0 11.0] | 0.780 |
| | SOFA Respiratory | | 626 | 3.0 [3.0 4.0] | 262 | 3.0 [2.0 4.0] | 364 | 3.0 [3.0 3.0] | 0.042 |
| | SOFA Nervous | | 508 | 3.0 [0.0 4.0] | 218 | 2.0 [0.0 4.0] | 290 | 4.0 [0.0 4.0] | 0.663 |
| | SOFA Cardiovascular | | 579 | 3.0 [3.0 4.0] | 242 | 3.0 [1.0 4.0] | 337 | 3.0 [3.0 4.0] | 0.004 |
| | SOFA Liver | | 588 | 0.0 [0.0 0.0] | 252 | 0.0 [0.0 0.0] | 336 | 0.0 [0.0 0.0] | 0.971 |
| | SOFA Coagulation | | 619 | 0.0 [0.0 0.0] | 257 | 0.0 [0.0 0.0] | 362 | 0.0 [0.0 0.0] | 0.281 |
| | SOFA Kidneys | | 629 | 0.0 [0.0 1.0] | 264 | 0.0 [0.0 1.0] | 365 | 0.0 [0.0 1.0] | 0.453 |
| FBC | Haemoglobin | g/dL | 619 | 114.0 [92.0 128.0] | 257 | 113.0 [90.0 126.0] | 362 | 115.0 [94.0 130.0] | 0.164 |
| | Haematocrit | | 312 | 0.4 [0.3 0.4] | 126 | 0.4 [0.3 0.4] | 186 | 0.4 [0.3 0.4] | 0.049 |
| | White blood cell count | x10^9/L | 619 | 9.6 [7.0 13.1] | 257 | 9.6 [6.9 12.7] | 362 | 9.6 [7.1 13.5] | 0.818 |
| | Neutrophils | x10^9/L | 618 | 8.1 [5.7 11.3] | 257 | 7.9 [5.4 11.1] | 361 | 8.3 [5.8 11.5] | 0.514 |
| | Monocytes | x10^9/L | 614 | 0.4 [0.3 0.7] | 256 | 0.5 [0.3 0.7] | 358 | 0.4 [0.3 0.7] | 0.084 |
| | Lymphocytes | x10^9/L | 615 | 0.8 [0.5 1.2] | 257 | 0.8 [0.6 1.3] | 358 | 0.8 [0.5 1.1] | 0.048 |
| | Basophils | x10^9/L | 498 | 0.0 [0.0 0.1] | 201 | 0.0 [0.0 0.1] | 297 | 0.0 [0.0 0.1] | 0.251 |
| | Eosinophils | x10^9/L | 493 | 0.0 [0.0 0.1] | 205 | 0.0 [0.0 0.1] | 288 | 0.0 [0.0 0.1] | 0.003 |
| Coag | Platelet Count | µmol/L | 619 | 246.0 [185.3 320.8] | 257 | 245.0 [191.0 325.0] | 362 | 246.0 [182.0 318.0] | 0.372 |
| | APTT | U/L | 398 | 32.1 [28.3 37.4] | 162 | 32.3 [28.3 38.0] | 236 | 32.1 [28.3 36.5] | 0.633 |
| | PT | U/L | 396 | 13.9 [12.4 15.2] | 162 | 13.6 [12.1 15.1] | 234 | 14.0 [12.7 15.4] | 0.297 |
| | INR | U/L | 212 | 1.1 [1.1 1.2] | 98 | 1.1 [1.1 1.2] | 114 | 1.1 [1.0 1.2] | 0.381 |
| | Fibrinogen | U/L | 410 | 6.8 [5.6 8.1] | 179 | 6.6 [5.7 7.7] | 231 | 6.9 [5.6 8.3] | 0.280 |
| | D-dimer | IU/L | 391 | 2642.0 [990.5 7701.3] | 177 | 2920.0 [977.5 8104.8] | 214 | 2418.0 [1060.0 7530.0] | 0.946 |
| Electrolytes | Blood Urea Nitrogen (BUN) | mmol/L | 498 | 7.4 [4.9 11.8] | 200 | 6.8 [4.5 10.8] | 298 | 7.8 [5.4 12.1] | 0.009 |
| | Creatinine | µmol/L | 621 | 88.0 [66.0 140.0] | 258 | 84.0 [64.0 129.0] | 363 | 90.0 [67.0 148.0] | 0.050 |
| | Sodium | mmol/L | 621 | 139.0 [136.0 142.0] | 259 | 139.0 [137.0 142.0] | 362 | 138.0 [135.0 141.0] | 0.003 |
| | Potassium | mmol/L | 620 | 4.4 [4.0 4.8] | 258 | 4.4 [4.0 4.8] | 362 | 4.4 [4.1 4.8] | 0.480 |
| Liver | Bilirubin | µmol/L | 588 | 10.0 [7.0 15.0] | 252 | 10.0 [7.0 14.0] | 336 | 10.0 [7.0 15.0] | 0.143 |
| | Alkaline Phosphatase | U/L | 600 | 77.0 [58.5 113.0] | 255 | 79.0 [58.3 120.8] | 345 | 75.0 [58.8 107.3] | 0.279 |
| | AST | U/L | 99 | 59.0 [39.3 85.0] | 43 | 56.0 [34.8 69.5] | 56 | 62.0 [41.5 90.5] | 0.128 |
| | ALT | U/L | 592 | 37.0 [24.0 59.0] | 252 | 36.0 [23.0 59.5] | 340 | 37.0 [24.0 59.0] | 0.649 |
| Inflammation | LDH (Lactate dehydrogenase) | IU/L | 194 | 649.0 [452.0 921.0] | 82 | 678.0 [443.0 1032.0] | 112 | 615.5 [459.5 873.0] | 0.224 |
| | Ferritin | ng/mL | 290 | 1218.5 [696.0 2320.0] | 129 | 1023.0 [665.8 2080.3] | 161 | 1374.0 [780.5 2492.3] | 0.031 |
| | CRP | | 592 | 215.7 [135.0 311.0] | 255 | 208.0 [119.3 308.9] | 337 | 218.0 [145.0 312.3] | 0.189 |
| | Procalcitonin | ug/L | 125 | 0.7 [0.3 2.2] | 61 | 0.6 [0.3 2.2] | 64 | 0.8 [0.3 2.1] | 0.456 |
| Cardiac | Creatinine Kinase | U/L | 243 | 217.0 [83.5 637.3] | 108 | 194.5 [79.0 563.0] | 135 | 226.0 [95.8 664.8] | 0.219 |
| | High sensitivity Troponin | | 253 | 21.8 [11.0 61.0] | 116 | 17.5 [9.0 48.3] | 137 | 25.7 [12.0 62.0] | 0.079 |
| | NT Pro BNP | pg/ml | 58 | 537.5 [165.0 1478.0] | 28 | 454.0 [196.5 1498.5] | 30 | 611.0 [161.0 1478.0] | 0.834 |
| Fluid | Cumulative Fluid balance | L | 589 | 343.0 [-212.3 1058.3] | 256 | 287.0 [-244.5 991.8] | 333 | 361.0 [-201.1 1160.3] | 0.412 |
| **Adjunctive interventions** | | | | | | | | | |
| | Was patient transferred in? | | 633 | 159 (25.1%) | 267 | 79 (29.6%) | 366 | 80 (21.9%) | 0.027 |
| | Tracheostomy | | 516 | 145 (28.1%) | 220 | 59 (26.8%) | 296 | 86 (29.1%) | 0.679 |
| | PEEP>10 | | 622 | 459 (73.8%) | 262 | 159 (60.7%) | 360 | 300 (83.3%) | 0.000 |
| | Neuro-muscular blockade | | 617 | 434 (70.3%) | 256 | 152 (59.4%) | 361 | 282 (78.1%) | 0.000 |
| | Prone positioning | | 551 | 273 (49.5%) | 234 | 90 (38.5%) | 317 | 183 (57.7%) | 0.000 |
| | Inhaled nitric oxide | | 521 | 73 (14%) | 224 | 21 (9.38%) | 297 | 52 (17.5%) | 0.014 |
| | Inhaled prostacyclin | | 521 | 55 (10.6%) | 224 | 18 (8.04%) | 297 | 37 (12.5%) | 0.137 |
| | Bronchoscopy | | 521 | 51 (9.79%) | 224 | 20 (8.93%) | 297 | 31 (10.4%) | 0.655 |
| | Renal replacement therapy | | 555 | 211 (38%) | 238 | 75 (31.5%) | 317 | 136 (42.9%) | 0.017 |
| | Diuretics | | 598 | 443 (74.1%) | 247 | 171 (69.2%) | 351 | 272 (77.5%) | 0.005 |
| | Corticosteroids | | 582 | 304 (52.2%) | 247 | 115 (46.6%) | 335 | 189 (56.4%) | 0.033 |
| | Therapeutic heparin | | 438 | 55 (12.6%) | 176 | 18 (10.2%) | 262 | 37 (14.1%) | 0.137 |
| | Antibiotics | | 313 | 219 (70%) | 127 | 74 (58.3%) | 186 | 145 (78%) | 0.002 |
| | **Prone** | | | | | | | | |
| | No of proning episodes | | 273 | 1.0 [1.0 2.0] | 90 | 1.0 [1.0 2.0] | 183 | 1.0 [1.0 2.0] | 0.941 |
| | mech vent prior first prone | days | 273 | 3.0 [1.8 6.0] | 90 | 2.0 [1.0 5.0] | 183 | 4.0 [2.0 7.0] | 0.007 |
| | durationn of first prone | days | 273 | 2.0 [1.0 4.0] | 90 | 2.0 [1.0 4.0] | 183 | 2.0 [1.0 4.0] | 0.943 |
| | responsders to proning | | 270 | 119 (44.1%) | 88 | 53 (60.2%) | 182 | 66 (36.3%) | 0.563 |
| | missed windows prior to prone | | 273 | 3.0 [1.0 6.0] | 90 | 2.0 [1.0 5.0] | 183 | 3.0 [1.0 7.0] | 0.026 |
| | missed windows - unproned | | 360 | 3.0 [1.0 10.0] | 177 | 1.0 [0.0 4.0] | 183 | 6.0 [3.0 13.0] | 0.000 |
| | **Tracheostomy** | | | | | | | | |
| | mech vent prior to tracheostomy | days | 145 | 13.0 [9.0 18.0] | 59 | 13.0 [7.0 16.0] | 86 | 15.0 [10.0 19.0] | 0.015 |
| | durationn of tracheostomy | days | 145 | 14.0 [6.0 21.3] | 59 | 11.0 [4.3 22.0] | 86 | 15.0 [8.0 21.0] | 0.078 |
| **End of life parameters** | | | | | | | | | |
| | life sustaining therapy withdrawn | | 130 | 85 (65.4%) | 19 | 12 (63.2%) | 111 | 73 (65.8%) | 0.000 |
| | cardiac arrest during admission | | 244 | 21 (8.61%) | 85 | 4 (4.71%) | 159 | 17 (10.7%) | 0.029 |

Table S7 - Time series mixed model ANOVA according to resolution of hypoxaemia over first week of mechanical ventilation.

| | | p-values | | | |
|---|---|---|---|---|---|
| **label** | **N** | **Resolvers vs Non-Resolvers** | **parameter over time** | **interaction** | **FDR interaction** |
| PaO_2 / FiO_2 | 472 | 0.000 | 0.000 | **0.000** | **0.000** |
| Dynamic Compliance | 341 | 0.408 | 0.039 | 0.226 | 0.379 |
| Respiratory rate | 458 | 0.556 | 0.000 | 0.678 | 0.721 |
| FiO_2 (%) | 503 | 0.000 | 0.000 | **0.000** | **0.000** |
| Tidal Volume per Kg | 347 | 0.782 | 0.411 | 0.624 | 0.711 |
| Tidal Volume | 433 | 0.193 | 0.612 | 0.654 | 0.721 |
| Minute ventilation | 404 | 0.292 | 0.000 | 0.516 | 0.626 |
| Peak pressure | 426 | 0.002 | 0.000 | **0.001** | **0.003** |
| PEEP | 390 | 0.001 | 0.000 | **0.001** | **0.003** |
| Dynamic delta pressure (peakP-peep) | 124 | 0.559 | 0.813 | 0.197 | 0.341 |
| Mean airway pressure | 256 | 0.004 | 0.000 | **0.000** | **0.000** |
| I:E ratio | 93 | 0.280 | 0.487 | 0.458 | 0.567 |
| Pressure support | 169 | 0.110 | 0.032 | 0.076 | 0.166 |
| Oxygen saturation | 417 | 0.000 | 0.175 | **0.001** | **0.004** |
| pH | 487 | 0.001 | 0.000 | **0.001** | **0.005** |
| PaO_2 | 484 | 0.008 | 0.033 | **0.000** | **0.001** |
| PaCO_2 | 487 | 0.035 | 0.003 | **0.000** | **0.000** |
| Base excess | 478 | 0.048 | 0.000 | 0.063 | 0.149 |
| HCO^-3 | 479 | 0.106 | 0.000 | **0.005** | **0.014** |
| Lactate | 451 | 0.001 | 0.310 | **0.026** | 0.069 |
| Oxygenation Index | 237 | 0.000 | 0.000 | **0.000** | **0.000** |
| Ventilatory Ratio | 314 | 0.301 | 0.026 | 0.103 | 0.195 |
| Mean arterial pressure (lowest) | 386 | 0.003 | 0.000 | **0.001** | **0.005** |
| Liver-Bilirubin | 167 | 0.262 | 0.040 | 0.158 | 0.285 |
| Daily fluid balance | 480 | 0.001 | 0.000 | 0.081 | 0.172 |
| Cumulative fluid balance | 480 | 0.015 | 0.074 | **0.001** | **0.005** |
| SOFA Score | 192 | 0.157 | 0.000 | **0.000** | **0.000** |
| Non-Respiratory SOFA | 224 | 0.349 | 0.000 | **0.001** | **0.004** |
| Glucose | 465 | 0.963 | 0.012 | 0.056 | 0.138 |
| BUN | 382 | 0.002 | 0.010 | **0.004** | **0.012** |
| Creatinine | 520 | 0.042 | 0.096 | **0.000** | **0.001** |
| Sodium | 523 | 0.796 | 0.008 | 0.258 | 0.421 |
| Potassium | 518 | 0.010 | 0.167 | 0.160 | 0.285 |
| Bilirubin | 401 | 0.284 | 0.000 | **0.002** | **0.007** |
| Alkaline Phosphatase | 441 | 0.171 | 0.000 | 0.706 | 0.721 |
| AST | 55 | 0.914 | 0.743 | 0.279 | 0.430 |
| ALT | 418 | 0.226 | 0.000 | 0.098 | 0.195 |
| Creatinine Kinase | 127 | 0.921 | 0.174 | 0.580 | 0.674 |
| LDH | 66 | 0.182 | 0.046 | 0.397 | 0.539 |
| Haemoglobin | 515 | 0.787 | 0.285 | 0.076 | 0.166 |
| WBC | 513 | 0.516 | 0.043 | 0.269 | 0.426 |
| RBC | 415 | 0.857 | 0.840 | 0.709 | 0.721 |
| Platelet Count | 511 | 0.048 | 0.000 | 0.099 | 0.195 |
| Haematocrit | 252 | 0.866 | 0.000 | **0.026** | 0.069 |
| Neutrophils | 511 | 0.994 | 0.442 | 0.309 | 0.452 |
| Monocytes | 509 | 0.005 | 0.000 | 0.303 | 0.452 |
| Lymphocytes | 509 | 0.124 | 0.000 | **0.026** | 0.069 |
| Basophils | 361 | 0.138 | 0.162 | 0.683 | 0.721 |
| Eosinophils | 386 | 0.001 | 0.000 | 0.564 | 0.670 |
| APTT | 255 | 0.236 | 0.660 | 0.326 | 0.465 |
| PT | 260 | 0.912 | 0.456 | 0.431 | 0.558 |
| INR | 163 | 0.815 | 0.175 | 0.440 | 0.558 |
| Fibrinogen | 137 | 0.063 | 0.062 | 0.690 | 0.721 |
| Ferritin | 75 | 0.201 | 0.472 | 0.404 | 0.539 |
| D-dimer | 147 | 0.967 | 0.388 | 0.406 | 0.539 |
| CRP | 457 | 0.001 | 0.000 | **0.000** | **0.000** |
| High sensitivity Troponin | 62 | 0.704 | 0.596 | 0.790 | 0.790 |

### Table S8 - Uni- and multi-variate model analysis of factors associated with progression of hypoxaemia over the first week of invasive mechanical ventilation

| | | Univariate | | | | Multivariate | | | |
|---|---|---|---|---|---|---|---|---|---|
| **FieldLabel** | **median [IQR]** | **Odds ratio** | **95% CI** | | **P Value** | **Odds ratio** | **95% CI** | | **P Value** |
| Age | | 1.366 | 1.186 | 1.572 | 0.000 | 1.236 | 1.060 | 1.441 | 0.007 |
| Male | | 1.314 | 0.882 | 1.959 | 0.179 | | | | |
| BMI | 28.13 [24.90 32.76] | 0.903 | 0.770 | 1.059 | 0.210 | | | | |
| Height | 173.00 [165.00 178.00] | 0.942 | 0.799 | 1.110 | 0.473 | | | | |
| symptoms days | 8.00 [6.00 12.00] | 0.857 | 0.724 | 1.013 | 0.071 | 0.940 | 0.784 | 1.127 | 0.503 |
| Hypertension | | 1.233 | 0.869 | 1.749 | 0.240 | | | | |
| Diabetes mellitus | | 1.549 | 1.047 | 2.293 | 0.029 | 1.381 | 0.886 | 2.153 | 0.155 |
| Oxygen saturation | 95.00 [93.00 98.00] | 0.864 | 0.735 | 1.016 | 0.077 | 0.998 | 0.834 | 1.194 | 0.981 |
| pH | 7.36 [7.30 7.42] | 0.919 | 0.789 | 1.070 | 0.274 | | | | |
| PaCO_2 | 5.96 [5.16 7.15] | 0.964 | 0.827 | 1.123 | 0.635 | | | | |
| HCO^-3 | 24.50 [22.48 26.70] | 0.784 | 0.671 | 0.915 | 0.002 | 0.870 | 0.733 | 1.032 | 0.111 |
| Lactate | 1.20 [1.00 1.60] | 1.197 | 1.024 | 1.398 | 0.024 | 1.030 | 0.862 | 1.231 | 0.748 |
| Peak pressure | 26.00 [23.00 30.00] | 1.158 | 0.992 | 1.351 | 0.062 | 1.158 | 0.967 | 1.385 | 0.110 |
| PEEP | 10.00 [8.00 12.00] | 1.268 | 1.067 | 1.508 | 0.007 | 1.109 | 0.902 | 1.363 | 0.328 |
| Minute ventilation | 8.53 [6.88 10.44] | 0.968 | 0.831 | 1.127 | 0.674 | | | | |
| Dynamic Comp | 31.47 [24.26 40.19] | 0.898 | 0.768 | 1.050 | 0.178 | | | | |
| Oxygenation Index | 8.10 [5.09 12.45] | 1.020 | 0.866 | 1.201 | 0.812 | | | | |
| Ventilatory Ratio | 1.54 [1.22 2.07] | 1.088 | 0.929 | 1.274 | 0.295 | | | | |
| Cum fluid balance | 343.00 [-212.25 1058.32] | 1.104 | 0.947 | 1.287 | 0.207 | | | | |
| Glucose | 8.39 [6.79 10.95] | 1.163 | 0.995 | 1.359 | 0.058 | 1.026 | 0.854 | 1.232 | 0.786 |
| BUN | 7.40 [4.90 11.80] | 1.237 | 1.057 | 1.449 | 0.008 | 1.183 | 0.983 | 1.422 | 0.075 |
| Sodium | 139.00 [136.00 142.00] | 0.793 | 0.678 | 0.926 | 0.004 | 0.818 | 0.686 | 0.974 | 0.024 |
| Potassium | 4.40 [4.00 4.80] | 1.064 | 0.912 | 1.241 | 0.430 | | | | |
| ALP | 77.00 [58.50 113.00] | 0.941 | 0.809 | 1.094 | 0.429 | | | | |
| ALT | 37.00 [24.00 59.00] | 1.050 | 0.902 | 1.223 | 0.526 | | | | |
| Haemoglobin | 114.00 [92.00 128.00] | 1.116 | 0.960 | 1.296 | 0.152 | | | | |
| Neutrophils | 8.10 [5.71 11.30] | 1.090 | 0.935 | 1.270 | 0.271 | | | | |
| Monocytes | 0.41 [0.30 0.70] | 0.844 | 0.725 | 0.982 | 0.028 | 0.921 | 0.767 | 1.106 | 0.376 |
| Lymphocytes | 0.80 [0.50 1.20] | 0.848 | 0.721 | 0.998 | 0.047 | 0.976 | 0.806 | 1.181 | 0.800 |
| Basophils | 0.00 [0.00 0.10] | 0.811 | 0.544 | 1.208 | 0.302 | | | | |
| Eosinophils | 0.00 [0.00 0.10] | 0.503 | 0.353 | 0.717 | 0.000 | 0.648 | 0.430 | 0.976 | 0.038 |
| APTT | 32.10 [28.30 37.40] | 0.926 | 0.781 | 1.098 | 0.376 | | | | |
| PT | 13.85 [12.40 15.20] | 1.065 | 0.903 | 1.256 | 0.456 | | | | |
| Fibrinogen | 6.80 [5.60 8.10] | 1.052 | 0.891 | 1.242 | 0.552 | | | | |
| D-dimer | 2642.00 [990.50 7701.25] | 0.964 | 0.818 | 1.134 | 0.655 | | | | |
| CRP | 215.65 [135.00 311.00] | 1.109 | 0.950 | 1.294 | 0.192 | | | | |
| SOFA Respiratory | | 0.929 | 0.769 | 1.123 | 0.447 | | | | |
| SOFA Nervous | | 1.028 | 0.942 | 1.121 | 0.541 | | | | |
| SOFA Cardio | | 1.241 | 1.068 | 1.442 | 0.005 | 1.197 | 1.011 | 1.418 | 0.037 |
| SOFA Liver | | 0.983 | 0.715 | 1.350 | 0.915 | | | | |
| SOFA Coagulation | | 1.207 | 0.865 | 1.683 | 0.267 | | | | |
| SOFA Kidneys | | 1.059 | 0.901 | 1.245 | 0.484 | | | | |
| SOFA score | | 1.036 | 0.968 | 1.109 | 0.301 | | | | |

* NonResolvers (N=318) vs Resolvers (N=231)

### Table S9 - Clinical and physiological characteristics, outcomes and interventions according to prone responsiveness.

| Clinical Characterisitcs | | | | | Prone responsivness | | | | |
|---|---|---|---|---|---|---|---|---|---|
| | | | ALL | | Responders | | Non-Responders | | |
| | label | units | Total N | median [IQR] / N (%) | Group N | median [IQR] / N (%) | Group N | median [IQR] / N (%) | P Value |
| | Male | | 270 | 203 (75.2%) | 119 | 93 (78.2%) | 151 | 110 (72.8%) | 0.316 |
| | White | | 219 | 115 (52.5%) | 90 | 43 (47.8%) | 129 | 72 (55.8%) | 0.057 |
| | Age | years | 270 | 60.0 [54.0 66.0] | 119 | 58.0 [50.0 64.0] | 151 | 60.0 [56.3 66.0] | 0.001 |
| | BMI | kg/m2 | 239 | 29.0 [25.3 33.2] | 107 | 29.4 [26.2 33.2] | 132 | 28.0 [24.8 33.2] | 0.220 |
| | Time since onset of symptoms | days | 181 | 7.0 [6.0 11.0] | 75 | 8.0 [6.0 12.8] | 106 | 7.0 [6.0 11.0] | 0.282 |
| | ICU length of stay | days | 270 | 18.0 [11.0 29.0] | 119 | 19.0 [12.3 30.8] | 151 | 16.0 [10.0 26.0] | 0.043 |
| | Length of mechanical ventilation | days | 270 | 16.0 [10.0 26.0] | 119 | 18.0 [11.0 28.5] | 151 | 15.0 [10.0 23.8] | 0.055 |
| | ICU Mortality | % | 270 | 142 (52.6%) | 119 | 37 (31.1%) | 151 | 105 (69.5%) | 0.000 |
| **Vent** | $FiO_2$ (%) | | 269 | 65.0 [50.0 80.0] | 119 | 60.0 [50.0 80.0] | 150 | 70.0 [60.0 80.0] | 0.022 |
| | $PaO_2$ to $FiO_2$ ratio | | 269 | 16.4 [12.1 21.9] | 119 | 17.9 [13.5 23.3] | 150 | 15.0 [11.4 19.6] | 0.000 |
| | Tidal Volume (mls/IBW) | ml/Kg(IBW) | 220 | 6.8 [6.1 7.8] | 99 | 6.8 [6.2 7.9] | 121 | 6.8 [6.0 7.7] | 0.581 |
| | Respiratory rate | bpm | 268 | 20.0 [16.0 24.0] | 118 | 20.0 [18.0 24.0] | 150 | 18.0 [16.0 22.0] | 0.021 |
| | Minute ventilation | L/minute | 256 | 8.7 [6.9 10.4] | 115 | 9.1 [7.7 10.6] | 141 | 8.5 [6.7 10.1] | 0.024 |
| | Peak pressure | ml/Kg(RBW) | 256 | 27.0 [24.0 30.0] | 117 | 27.0 [24.0 29.0] | 139 | 28.0 [25.0 30.8] | 0.032 |
| | Plateau pressure | ml/Kg(IBW) | 48 | 28.0 [24.0 29.0] | 20 | 26.5 [22.0 29.5] | 28 | 28.0 [25.5 28.5] | 0.612 |
| | PEEP | cmH_20 | 259 | 10.0 [10.0 12.0] | 116 | 10.0 [10.0 12.0] | 143 | 10.8 [10.0 14.0] | 0.124 |
| | Mean airway pressure | cmH_20 | 143 | 17.0 [14.3 19.0] | 73 | 17.0 [15.0 18.0] | 70 | 16.5 [14.0 21.0] | 0.805 |
| | Pressure support | cmH_20 | 150 | 10.0 [8.0 14.0] | 60 | 10.0 [7.0 15.0] | 90 | 10.0 [8.0 14.0] | 0.504 |
| | Dynamic Compliance | mls/cmH20 | 237 | 30.0 [24.0 38.1] | 109 | 31.0 [23.5 39.9] | 128 | 29.4 [24.4 36.6] | 0.509 |
| | Oxygenation Index | | 143 | 9.3 [6.4 13.6] | 73 | 8.7 [5.9 11.1] | 70 | 11.0 [6.9 16.2] | 0.003 |
| | Ventilatory Ratio | | 219 | 1.7 [1.4 2.2] | 98 | 1.8 [1.4 2.2] | 121 | 1.7 [1.2 2.3] | 0.617 |
| **ABG** | Oxygen saturation | | 242 | 94.5 [92.0 97.0] | 113 | 95.0 [92.0 97.0] | 129 | 94.0 [92.0 96.0] | 0.167 |
| | pH | | 270 | 7.3 [7.3 7.4] | 119 | 7.3 [7.3 7.4] | 151 | 7.4 [7.3 7.4] | 0.916 |
| | $PaO_2$ | kPa | 270 | 10.6 [9.0 12.4] | 119 | 10.5 [9.2 12.6] | 151 | 10.6 [8.8 12.3] | 0.539 |
| | $PaCO_2$ | kPa | 270 | 6.2 [5.3 7.4] | 119 | 6.2 [5.3 7.3] | 151 | 6.4 [5.4 7.4] | 0.358 |
| | Base excess | | 270 | 0.2 [-2.6 2.7] | 119 | 0.2 [-2.4 2.3] | 151 | 0.2 [-2.6 3.2] | 0.794 |
| | $HCO_3^-$ | mmol/L | 269 | 24.4 [22.4 27.0] | 118 | 24.6 [22.4 26.7] | 151 | 24.2 [22.4 27.1] | 0.739 |
| | Lactate | mmol/L | 256 | 1.2 [1.0 1.6] | 110 | 1.1 [0.9 1.5] | 146 | 1.3 [1.1 1.7] | 0.000 |
| **SOFA** | SOFA score | | 174 | 9.0 [7.0 11.0] | 75 | 8.0 [7.0 10.0] | 99 | 9.0 [7.3 11.0] | 0.239 |
| | SOFA Respiratory | | 269 | 3.0 [3.0 4.0] | 119 | 3.0 [3.0 3.0] | 150 | 3.0 [3.0 4.0] | 0.000 |
| | SOFA Nervous | | 205 | 4.0 [0.0 4.0] | 88 | 4.0 [0.0 4.0] | 117 | 4.0 [0.0 4.0] | 0.208 |
| | SOFA Cardiovascular | | 248 | 3.0 [1.0 4.0] | 110 | 3.0 [1.0 3.0] | 138 | 3.0 [3.0 4.0] | 0.000 |
| | SOFA Liver | | 252 | 0.0 [0.0 0.0] | 110 | 0.0 [0.0 0.0] | 142 | 0.0 [0.0 0.0] | 0.968 |
| | SOFA Coagulation | | 267 | 0.0 [0.0 0.0] | 116 | 0.0 [0.0 0.0] | 151 | 0.0 [0.0 0.0] | 0.877 |
| | SOFA Kidneys | | 269 | 0.0 [0.0 1.0] | 118 | 0.0 [0.0 1.0] | 151 | 0.0 [0.0 1.0] | 0.257 |
| **FBC** | Haemoglobin | g/dL | 267 | 116.0 [91.3 128.0] | 116 | 115.5 [95.0 127.8] | 151 | 117.0 [88.3 129.0] | 0.783 |
| | Haematocrit | | 178 | 0.4 [0.3 0.4] | 86 | 0.4 [0.3 0.4] | 92 | 0.4 [0.3 0.4] | 0.564 |
| | White blood cell count | x10^9/L | 267 | 9.7 [7.3 12.8] | 116 | 9.6 [7.1 12.6] | 151 | 9.8 [7.5 12.9] | 0.643 |
| | Neutrophils | x10^9/L | 267 | 8.3 [6.1 11.3] | 116 | 8.1 [5.6 11.1] | 151 | 8.3 [6.5 11.7] | 0.347 |
| | Monocytes | x10^9/L | 264 | 0.4 [0.3 0.6] | 115 | 0.4 [0.3 0.6] | 149 | 0.4 [0.3 0.6] | 0.808 |
| | Lymphocytes | x10^9/L | 265 | 0.7 [0.5 1.1] | 115 | 0.8 [0.5 1.2] | 150 | 0.7 [0.5 1.1] | 0.632 |
| | Basophils | x10^9/L | 223 | 0.0 [0.0 0.1] | 100 | 0.0 [0.0 0.1] | 123 | 0.0 [0.0 0.0] | 0.517 |
| | Eosinophils | x10^9/L | 220 | 0.0 [0.0 0.1] | 101 | 0.0 [0.0 0.1] | 119 | 0.0 [0.0 0.1] | 0.412 |
| **Coag** | Platelet Count | µmol/L | 267 | 247.0 [190.3 331.0] | 116 | 247.5 [196.0 329.5] | 151 | 247.0 [190.0 333.5] | 0.812 |
| | APTT | U/L | 193 | 31.7 [28.0 35.3] | 86 | 31.8 [28.0 35.6] | 107 | 31.4 [27.8 34.9] | 0.451 |
| | PT | U/L | 192 | 13.6 [12.1 14.9] | 86 | 13.3 [12.1 14.7] | 106 | 13.8 [12.1 15.4] | 0.196 |
| | INR | U/L | 127 | 1.1 [1.1 1.2] | 66 | 1.1 [1.1 1.2] | 61 | 1.1 [1.1 1.2] | 0.521 |
| | Fibrinogen | U/L | 188 | 6.9 [5.7 8.2] | 93 | 6.9 [5.6 8.0] | 95 | 7.1 [5.9 8.3] | 0.144 |
| | D-dimer | IU/L | 171 | 2540.0 [947.0 6208.5] | 73 | 1694.0 [880.0 3895.0] | 98 | 3213.5 [1385.0 8329.0] | 0.009 |
| **Electrolytes** | Blood Urea Nitrogen (BUN) | mmol/L | 189 | 7.0 [5.0 11.0] | 67 | 6.6 [5.0 9.9] | 122 | 7.6 [5.1 11.9] | 0.345 |
| | Creatinine | µmol/L | 267 | 84.0 [63.0 125.8] | 117 | 85.0 [65.8 133.3] | 150 | 82.0 [63.0 118.0] | 0.309 |
| | Sodium | mmol/L | 268 | 138.0 [136.0 141.0] | 118 | 138.0 [135.0 140.0] | 150 | 139.0 [136.0 143.0] | 0.128 |
| | Potassium | mmol/L | 268 | 4.4 [4.1 4.7] | 118 | 4.5 [4.2 4.8] | 150 | 4.4 [4.0 4.7] | 0.204 |
| **Liver** | Bilirubin | µmol/L | 252 | 10.0 [7.0 15.0] | 110 | 10.0 [7.0 15.0] | 142 | 10.0 [8.0 15.0] | 0.245 |
| | Alkaline Phosphatase | U/L | 257 | 76.0 [56.8 109.0] | 113 | 76.0 [53.0 110.0] | 144 | 75.5 [60.5 108.0] | 0.548 |
| | AST | U/L | 52 | 61.0 [41.0 80.5] | 22 | 59.0 [42.0 91.0] | 30 | 61.5 [40.0 79.0] | 0.985 |
| | ALT | U/L | 253 | 37.0 [26.0 56.3] | 111 | 37.0 [26.3 57.8] | 142 | 36.5 [25.0 56.0] | 0.978 |
| **Inflammation** | LDH (Lactate dehydrogenase) | IU/L | 110 | 712.5 [520.0 1004.0] | 55 | 783.0 [532.0 1141.8] | 55 | 677.0 [514.0 968.3] | 0.404 |
| | Ferritin | ng/mL | 149 | 1297.0 [711.8 2520.0] | 68 | 1038.0 [629.5 2215.0] | 81 | 1410.0 [773.3 2776.8] | 0.115 |
| | CRP | | 252 | 235.0 [158.6 322.6] | 117 | 229.8 [155.5 323.3] | 135 | 239.3 [160.0 321.7] | 0.597 |
| | Procalcitonin | ug/L | 47 | 0.9 [0.4 2.4] | 19 | 0.6 [0.3 2.3] | 28 | 1.2 [0.6 2.9] | 0.190 |
| **Cardiac** | Creatinine Kinase | U/L | 122 | 248.5 [122.0 638.0] | 61 | 373.0 [187.1 847.3] | 61 | 155.0 [92.8 515.8] | 0.004 |
| | High sensitivity Troponin | | 132 | 21.9 [11.8 57.5] | 65 | 15.0 [8.8 50.8] | 67 | 25.0 [14.4 61.8] | 0.022 |
| | NT Pro BNP | pg/ml | 26 | 621.0 [173.0 1983.0] | 10 | 313.5 [165.0 680.0] | 16 | 810.5 [502.0 2587.0] | 0.058 |
| **Fluid** | Cumulative Fluid balance | L | 251 | 384.0 [-196.6 1160.0] | 109 | 143.0 [-272.3 753.8] | 142 | 622.0 [-114.0 1363.0] | 0.004 |
| **Adjunctive interventions** | | | | | | | | | |
| | Was patient transferred in? | | 270 | 87 (32.2%) | 119 | 48 (40.3%) | 151 | 39 (25.8%) | 0.011 |
| | Tracheostomy | | 243 | 77 (31.7%) | 113 | 41 (36.3%) | 130 | 36 (27.7%) | 0.055 |
| | PEEP>10 | | 268 | 239 (89.2%) | 119 | 102 (85.7%) | 149 | 137 (91.9%) | 0.199 |
| | Neuro-muscular blockade | | 269 | 246 (91.4%) | 118 | 107 (90.7%) | 151 | 139 (92.1%) | 0.540 |
| | Prone positioning | | 270 | 270 (100%) | 119 | 119 (100%) | 151 | 151 (100%) | |
| | Inhaled nitric oxide | | 243 | 66 (27.2%) | 113 | 20 (17.7%) | 130 | 46 (35.4%) | 0.010 |
| | Inhaled prostacyclin | | 243 | 35 (14.4%) | 113 | 9 (7.96%) | 130 | 26 (20%) | 0.019 |
| | Bronchoscopy | | 243 | 34 (14%) | 113 | 18 (15.9%) | 130 | 16 (12.3%) | 0.265 |
| | Renal replacement therapy | | 250 | 112 (44.8%) | 115 | 48 (41.7%) | 135 | 64 (47.4%) | 0.735 |
| | Diuretics | | 268 | 210 (78.4%) | 118 | 90 (76.3%) | 150 | 120 (80%) | 0.451 |
| | Corticosteroids | | 264 | 154 (58.3%) | 118 | 65 (55.1%) | 146 | 89 (61%) | 0.477 |
| | Therapeutic heparin | | 187 | 33 (17.6%) | 75 | 14 (18.7%) | 112 | 19 (17%) | 0.839 |
| | Antibiotics | | 179 | 119 (66.5%) | 87 | 46 (52.9%) | 92 | 73 (79.3%) | 0.111 |
| | **Prone** | | | | | | | | |
| | No of proning episodes | | 270 | 1.0 [1.0 2.0] | 119 | 1.0 [1.0 2.0] | 151 | 1.0 [1.0 2.0] | 0.073 |
| | mech vent prior first prone | days | 270 | 3.0 [1.0 6.0] | 119 | 3.0 [2.0 5.8] | 151 | 3.0 [1.0 7.0] | 0.357 |
| | durationn of first prone | days | 270 | 2.0 [1.0 4.0] | 119 | 2.0 [1.0 4.0] | 151 | 2.0 [1.0 4.0] | 0.413 |
| | responsders to proning | | 270 | 119 (44.1%) | 119 | 119 (100%) | 151 | 0 (0%) | 0.000 |
| | missed windows prior to prone | | 270 | 3.0 [1.0 6.0] | 119 | 2.0 [1.0 5.0] | 151 | 3.0 [1.0 7.0] | 0.022 |
| | | | 0 | NaN [NaN NaN] | 0 | NaN [NaN NaN] | 0 | NaN [NaN NaN] | |
| | **Tracheostomy** | | | | | | | | |
| | mech vent prior to tracheostomy | days | 77 | 16.0 [11.0 20.0] | 41 | 16.0 [12.0 19.3] | 36 | 14.5 [8.5 20.0] | 0.352 |
| | durationn of tracheostomy | days | 77 | 14.0 [6.0 21.3] | 41 | 15.0 [7.8 22.0] | 36 | 11.5 [4.0 20.0] | 0.129 |
| **End of life parameters** | | | | | | | | | |
| | life sustaining therapy withdrawn | | 90 | 61 (67.8%) | 31 | 22 (71%) | 59 | 39 (66.1%) | 0.152 |
| | cardiac arrest during admission | | 139 | 13 (9.35%) | 58 | 7 (12.1%) | 81 | 6 (7.41%) | 0.467 |

#### Table S10 - Time series mixed model ANOVA according to prone responsiveness.

| | | p-values | | | |
|---|---|---|---|---|---|
| **label** | **N** | **Prone Responders vs non-Responders** | **parameter over time** | **interaction** | **FDR interaction** |
| PaO_2 / FiO_2 | 234 | 0.000 | 0.000 | **0.000** | **0.000** |
| Dynamic Compliance | 171 | 0.813 | 0.819 | 0.817 | 0.872 |
| Respiratory rate | 219 | 0.993 | 0.645 | 0.143 | 0.590 |
| FiO_2 (%) | 239 | 0.000 | 0.000 | **0.000** | **0.001** |
| Tidal Volume per Kg | 186 | 0.477 | 0.779 | 0.867 | 0.883 |
| Tidal Volume | 216 | 0.186 | 0.659 | 0.799 | 0.872 |
| Minute ventilation | 197 | 0.803 | 0.449 | 0.368 | 0.619 |
| Peak pressure | 212 | 0.002 | 0.649 | 0.333 | 0.619 |
| PEEP | 197 | 0.699 | 0.929 | 0.826 | 0.872 |
| Dynamic delta pressure (peakP-peep) | 78 | 0.014 | 0.487 | 0.929 | 0.929 |
| Mean airway pressure | 117 | 0.670 | 0.019 | **0.003** | **0.021** |
| I:E ratio | 42 | 0.881 | 0.923 | 0.711 | 0.811 |
| Pressure support | 84 | 0.775 | 0.766 | 0.703 | 0.811 |
| Oxygen saturation | 218 | 0.000 | 0.119 | **0.002** | **0.015** |
| pH | 239 | 0.047 | 0.021 | 0.316 | 0.619 |
| PaO_2 | 239 | 0.000 | 0.011 | **0.001** | **0.010** |
| PaCO_2 | 239 | 0.004 | 0.003 | 0.442 | 0.619 |
| Base excess | 234 | 0.689 | 0.000 | 0.367 | 0.619 |
| HCO^-3 | 236 | 0.529 | 0.000 | 0.857 | 0.883 |
| Lactate | 216 | 0.000 | 0.505 | 0.385 | 0.619 |
| Oxygenation Index | 113 | 0.000 | 0.001 | **0.001** | **0.010** |
| Ventilatory Ratio | 167 | 0.053 | 0.404 | 0.427 | 0.619 |
| Mean arterial pressure (lowest) | 198 | 0.320 | 0.409 | 0.513 | 0.676 |
| Liver-Bilirubin | 70 | 0.668 | 0.727 | 0.696 | 0.811 |
| Daily fluid balance | 227 | 0.038 | 0.295 | 0.257 | 0.619 |
| Cumulative fluid balance | 227 | 0.035 | 0.129 | 0.108 | 0.558 |
| SOFA Score | 97 | 0.045 | 0.445 | 0.298 | 0.619 |
| Non-Respiratory SOFA | 108 | 0.100 | 0.541 | 0.346 | 0.619 |
| Glucose | 208 | 0.930 | 0.771 | 0.677 | 0.811 |
| BUN | 158 | 0.703 | 0.000 | 0.785 | 0.872 |
| Creatinine | 249 | 0.855 | 0.757 | 0.065 | 0.372 |
| Sodium | 251 | 0.192 | 0.036 | 0.194 | 0.619 |
| Potassium | 246 | 0.915 | 0.492 | 0.413 | 0.619 |
| Bilirubin | 192 | 0.884 | 0.277 | 0.227 | 0.619 |
| Alkaline Phosphatase | 205 | 0.808 | 0.000 | 0.342 | 0.619 |
| AST | 35 | 0.098 | 0.054 | 0.225 | 0.619 |
| ALT | 197 | 0.054 | 0.000 | **0.000** | **0.005** |
| Creatinine Kinase | 88 | 0.132 | 0.417 | 0.419 | 0.619 |
| LDH | 52 | 0.162 | 0.374 | 0.359 | 0.619 |
| Haemoglobin | 246 | 0.433 | 0.202 | 0.351 | 0.619 |
| WBC | 246 | 0.441 | 0.001 | 0.138 | 0.590 |
| RBC | 220 | 0.074 | 0.651 | 0.582 | 0.737 |
| Platelet Count | 244 | 0.040 | 0.000 | **0.000** | **0.005** |
| Haematocrit | 162 | 0.608 | 0.000 | 0.437 | 0.619 |
| Neutrophils | 246 | 0.143 | 0.057 | 0.340 | 0.619 |
| Monocytes | 244 | 0.956 | 0.000 | 0.522 | 0.676 |
| Lymphocytes | 245 | 0.881 | 0.000 | **0.043** | 0.275 |
| Basophils | 191 | 0.683 | 0.037 | 0.468 | 0.635 |
| Eosinophils | 193 | 0.874 | 0.002 | 0.153 | 0.590 |
| APTT | 137 | 0.424 | 0.800 | 0.420 | 0.619 |
| PT | 140 | 0.718 | 0.306 | 0.445 | 0.619 |
| INR | 110 | 0.381 | 0.667 | 0.266 | 0.619 |
| Fibrinogen | 80 | 0.033 | 0.021 | 0.654 | 0.811 |
| Ferritin | 55 | 0.224 | 0.565 | 0.344 | 0.619 |
| D-dimer | 70 | 0.009 | 0.562 | 0.374 | 0.619 |
| CRP | 217 | 0.203 | 0.042 | 0.227 | 0.619 |
| High sensitivity Troponin | 47 | 0.963 | 0.395 | 0.155 | 0.590 |

#### Table S11 - Uni- and multi-variate model analysis of pre-pronation factors associated prone responsiveness.

| | | Univariate | | | | Multivariate | | | |
|---|---|---|---|---|---|---|---|---|---|
| **FieldLabel** | **median [IQR]** | **Odds ratio** | **95% CI** | | **P Value** | **Odds ratio** | **95% CI** | | **P Value** |
| Age | | 1.555 | 1.197 | 2.019 | 0.001 | 1.557 | 1.165 | 2.081 | 0.003 |
| Male | | 0.875 | 0.474 | 1.617 | 0.671 | | | | |
| BMI | 29.00 [25.28 33.18] | 0.865 | 0.681 | 1.099 | 0.234 | | | | |
| Height | 172.35 [165.00 178.00] | 1.045 | 0.821 | 1.330 | 0.718 | | | | |
| symptoms days | 7.00 [6.00 11.00] | 1.039 | 0.810 | 1.332 | 0.764 | | | | |
| Hypertension | | 1.061 | 0.616 | 1.827 | 0.832 | | | | |
| Diabetes mellitus | | 1.393 | 0.749 | 2.591 | 0.296 | | | | |
| Oxygen saturation | 93.00 [91.00 96.00] | 0.758 | 0.589 | 0.975 | 0.031 | 0.982 | 0.714 | 1.351 | 0.911 |
| pH | 7.37 [7.31 7.42] | 0.865 | 0.683 | 1.094 | 0.225 | | | | |
| PaCO_2 | 6.70 [5.80 7.67] | 1.257 | 0.989 | 1.598 | 0.062 | 1.198 | 0.890 | 1.611 | 0.234 |
| HCO^-3 | 26.60 [24.00 30.65] | 0.988 | 0.784 | 1.247 | 0.922 | | | | |
| Lactate | 1.30 [1.10 1.80] | 1.610 | 1.260 | 2.057 | 0.000 | 1.331 | 0.992 | 1.785 | 0.057 |
| Peak pressure | 28.06 [25.00 31.00] | 1.568 | 1.232 | 1.995 | 0.000 | 1.423 | 1.063 | 1.906 | 0.018 |
| PEEP | 11.00 [10.00 13.00] | 1.359 | 1.062 | 1.738 | 0.015 | 1.256 | 0.927 | 1.702 | 0.142 |
| Minute ventilation | 9.88 [8.19 11.96] | 1.083 | 0.854 | 1.374 | 0.510 | | | | |
| Dynamic Comp | 30.14 [24.00 38.59] | 0.981 | 0.770 | 1.251 | 0.880 | | | | |
| Ventilatory Ratio | 2.11 [1.61 2.57] | 1.159 | 0.903 | 1.488 | 0.246 | | | | |
| Cum fluid balance | 1743.00 [216.14 4269.60] | 1.445 | 1.133 | 1.843 | 0.003 | 1.232 | 0.919 | 1.650 | 0.162 |
| Glucose | 9.10 [7.50 11.10] | 0.982 | 0.775 | 1.244 | 0.882 | | | | |
| BUN | 10.00 [6.93 15.60] | 1.095 | 0.857 | 1.399 | 0.466 | | | | |
| Sodium | 141.00 [139.00 146.00] | 1.217 | 0.964 | 1.537 | 0.099 | 1.189 | 0.890 | 1.589 | 0.241 |
| Potassium | 4.53 [4.30 4.90] | 1.107 | 0.877 | 1.397 | 0.392 | | | | |
| ALP | 85.00 [65.00 136.00] | 0.991 | 0.782 | 1.255 | 0.940 | | | | |
| ALT | 40.00 [27.00 58.00] | 0.834 | 0.659 | 1.056 | 0.132 | | | | |
| Haemoglobin | 103.00 [75.50 116.88] | 0.931 | 0.738 | 1.174 | 0.543 | | | | |
| Haematocrit | 0.33 [0.30 0.38] | 0.970 | 0.753 | 1.250 | 0.816 | | | | |
| Neutrophils | 8.80 [6.92 11.78] | 1.149 | 0.910 | 1.453 | 0.244 | | | | |
| Monocytes | 0.40 [0.30 0.70] | 0.920 | 0.718 | 1.179 | 0.510 | | | | |
| Lymphocytes | 0.80 [0.55 1.10] | 0.898 | 0.715 | 1.128 | 0.356 | | | | |
| Basophils | 0.02 [0.00 0.10] | 1.232 | 0.921 | 1.650 | 0.160 | | | | |
| Eosinophils | 0.10 [0.00 0.20] | 0.986 | 0.715 | 1.360 | 0.931 | | | | |
| APTT | 32.83 [29.00 37.85] | 0.874 | 0.681 | 1.123 | 0.293 | | | | |
| PT | 14.00 [12.10 15.30] | 1.138 | 0.883 | 1.466 | 0.318 | | | | |
| Fibrinogen | 7.30 [6.10 8.91] | 0.967 | 0.758 | 1.235 | 0.790 | | | | |
| D-dimer | 4017.50 [1630.00 10580.00] | 1.141 | 0.886 | 1.470 | 0.308 | | | | |
| CRP | 277.50 [183.45 339.00] | 1.080 | 0.850 | 1.371 | 0.530 | | | | |
| SOFA Respiratory | 3.00 [3.00 3.00] | 1.894 | 1.352 | 2.654 | 0.000 | 1.708 | 1.168 | 2.499 | 0.006 |
| SOFA Nervous | | 0.932 | 0.810 | 1.072 | 0.326 | | | | |
| SOFA Cardio | | 1.442 | 1.168 | 1.781 | 0.001 | 1.346 | 1.039 | 1.742 | 0.024 |
| SOFA Liver | | 0.933 | 0.615 | 1.414 | 0.744 | | | | |
| SOFA Coagulation | | 1.616 | 0.866 | 3.017 | 0.132 | | | | |
| SOFA Kidneys | | 0.948 | 0.749 | 1.199 | 0.654 | | | | |
| Prone initiation day | | 1.044 | 0.981 | 1.110 | 0.175 | | | | |

* non-resp (N=127) vs repsonsive (N=101)

#### Table S12 - Uni- and multi-variate model analysis of post- pronation factors associated prone responsiveness.

| | | Univariate | | | | Multivariate | | | |
|---|---|---|---|---|---|---|---|---|---|
| FieldLabel | median [IQR] | Odds ratio | 95% CI | | P Value | Odds ratio | 95% CI | | P Value |
| Age | | 1.485 | 1.164 | 1.893 | 0.001 | 1.431 | 1.082 | 1.892 | 0.012 |
| Male | | 1.344 | 0.734 | 2.460 | 0.338 | | | | |
| BMI | 29.00 [25.28 33.18] | 0.849 | 0.672 | 1.073 | 0.171 | | | | |
| Height | 172.35 [165.00 178.00] | 1.057 | 0.835 | 1.339 | 0.643 | | | | |
| symptoms days | 7.00 [6.00 11.00] | 1.057 | 0.829 | 1.347 | 0.654 | | | | |
| Hypertension | | 1.085 | 0.644 | 1.828 | 0.758 | | | | |
| Diabetes mellitus | | 1.395 | 0.777 | 2.505 | 0.265 | | | | |
| Oxygen saturation | 93.00 [91.00 95.00] | 0.683 | 0.540 | 0.864 | 0.001 | 0.799 | 0.603 | 1.059 | 0.119 |
| pH | 7.35 [7.29 7.41] | 0.851 | 0.680 | 1.065 | 0.160 | | | | |
| PaCO_2 | 6.85 [6.01 8.10] | 1.225 | 0.976 | 1.536 | 0.080 | 1.210 | 0.930 | 1.575 | 0.156 |
| HCO^-3 | 26.80 [23.90 30.20] | 1.011 | 0.808 | 1.264 | 0.926 | | | | |
| Lactate | 1.30 [1.00 1.70] | 1.640 | 1.290 | 2.086 | 0.000 | 1.316 | 0.998 | 1.737 | 0.052 |
| Peak pressure | 28.00 [25.88 31.00] | 1.468 | 1.163 | 1.854 | 0.001 | 1.432 | 1.101 | 1.862 | 0.007 |
| PEEP | 11.00 [10.00 13.00] | 1.214 | 0.961 | 1.535 | 0.104 | | | | |
| Minute ventilation | 9.89 [7.91 11.38] | 0.956 | 0.764 | 1.196 | 0.693 | | | | |
| Dynamic Comp | 30.18 [21.83 40.96] | 0.924 | 0.733 | 1.166 | 0.506 | | | | |
| Ventilatory Ratio | 2.05 [1.64 2.50] | 1.184 | 0.934 | 1.501 | 0.162 | | | | |
| Cum fluid balance | 2319.00 [389.06 5227.73] | 1.217 | 0.965 | 1.536 | 0.098 | 0.922 | 0.693 | 1.227 | 0.579 |
| Glucose | 8.95 [7.39 11.33] | 1.181 | 0.933 | 1.494 | 0.167 | | | | |
| BUN | 10.65 [6.90 16.35] | 1.152 | 0.911 | 1.458 | 0.238 | | | | |
| Sodium | 142.00 [138.00 146.00] | 1.151 | 0.916 | 1.446 | 0.227 | | | | |
| Potassium | 4.60 [4.30 5.00] | 0.963 | 0.766 | 1.212 | 0.748 | | | | |
| ALP | 94.00 [67.00 142.25] | 1.051 | 0.837 | 1.320 | 0.666 | | | | |
| ALT | 41.00 [30.00 61.00] | 0.928 | 0.738 | 1.168 | 0.525 | | | | |
| Haemoglobin | 100.00 [79.00 117.25] | 0.941 | 0.750 | 1.179 | 0.596 | | | | |
| Haematocrit | 0.34 [0.29 0.37] | 0.922 | 0.727 | 1.171 | 0.507 | | | | |
| Neutrophils | 8.60 [6.86 11.90] | 1.128 | 0.903 | 1.410 | 0.289 | | | | |
| Monocytes | 0.43 [0.30 0.70] | 0.818 | 0.652 | 1.027 | 0.083 | 0.758 | 0.581 | 0.990 | 0.042 |
| Lymphocytes | 0.72 [0.50 1.10] | 0.865 | 0.687 | 1.090 | 0.219 | | | | |
| Basophils | 0.01 [0.00 0.10] | 0.965 | 0.712 | 1.307 | 0.817 | | | | |
| Eosinophils | 0.10 [0.00 0.20] | 0.947 | 0.697 | 1.285 | 0.725 | | | | |
| APTT | 32.40 [28.65 37.82] | 1.042 | 0.821 | 1.323 | 0.733 | | | | |
| PT | 13.70 [11.90 14.95] | 1.117 | 0.881 | 1.416 | 0.360 | | | | |
| CRP | 289.00 [183.50 348.75] | 0.946 | 0.751 | 1.192 | 0.638 | | | | |
| SOFA Respiratory | 3.00 [3.00 4.00] | 2.233 | 1.513 | 3.297 | 0.000 | 1.946 | 1.231 | 3.078 | 0.004 |
| SOFA Nervous | | 0.910 | 0.791 | 1.046 | 0.186 | | | | |
| SOFA Cardio | | 1.420 | 1.163 | 1.735 | 0.001 | 1.134 | 0.893 | 1.441 | 0.301 |
| SOFA Liver | | 0.996 | 0.654 | 1.518 | 0.986 | | | | |
| SOFA Coagulation | | 1.190 | 0.726 | 1.951 | 0.490 | | | | |
| SOFA Kidneys | | 0.932 | 0.747 | 1.162 | 0.531 | | | | |
| Prone initiation day | | 1.044 | 0.982 | 1.111 | 0.167 | | | | |

* non-resp (N=138) vs repsonsive (N=109)

### Table S13- Clinical and physiological characteristics, outcomes and interventions according to ICU outcome.

| Clinical Characterisitcs | | | | | Outcome | | | | |
|---|---|---|---|---|---|---|---|---|---|
| | | | ALL | | Survivors | | Non-Survivors | | |
| | label | units | Total N | median [IQR] / N (%) | Group N | median [IQR] / N (%) | Group N | median [IQR] / N (%) | P Value |
| | Male | | 633 | 481 (76%) | 365 | 263 (72.1%) | 268 | 218 (81.3%) | 0.007 |
| | White | | 542 | 250 (46.1%) | 310 | 143 (46.1%) | 232 | 107 (46.1%) | 0.849 |
| | Age | years | 633 | 59.0 [51.0 66.0] | 365 | 56.0 [47.0 63.0] | 268 | 63.0 [57.0 70.0] | 0.000 |
| | BMI | kg/m2 | 524 | 28.1 [24.9 32.8] | 314 | 28.7 [25.1 33.6] | 210 | 27.8 [24.7 31.2] | 0.134 |
| | Time since onset of symptoms | days | 408 | 8.0 [6.0 12.0] | 242 | 9.0 [6.0 12.0] | 166 | 7.0 [5.0 12.0] | 0.101 |
| | ICU length of stay | days | 633 | 14.0 [8.0 23.0] | 365 | 17.0 [9.8 30.0] | 268 | 11.0 [7.0 18.0] | 0.000 |
| | Length of mechanical ventilation | days | 633 | 13.0 [7.0 22.0] | 365 | 15.0 [8.0 28.0] | 268 | 11.0 [6.0 16.0] | 0.000 |
| | ICU Mortality | % | 633 | 268 (42.3%) | 365 | 0 (0%) | 268 | 268 (100%) | 0.000 |
| **Vent** | $FiO_2$ (%) | | 628 | 60.0 [45.0 80.0] | 360 | 55.5 [40.0 70.0] | 268 | 65.0 [55.0 80.0] | 0.000 |
| | $PaO_2$ to $FiO_2$ ratio | | 626 | 18.3 [13.0 25.0] | 359 | 19.8 [13.9 27.5] | 267 | 17.0 [12.3 23.0] | 0.000 |
| | Tidal Volume (mls/IBW) | ml/Kg(IBW) | 472 | 6.8 [6.0 7.8] | 286 | 6.9 [6.1 7.8] | 186 | 6.8 [6.0 7.7] | 0.351 |
| | Respiratory rate | bpm | 627 | 18.8 [16.0 22.0] | 360 | 18.0 [16.0 22.0] | 267 | 20.0 [16.0 22.0] | 0.357 |
| | Minute ventilation | L/minute | 606 | 8.5 [6.9 10.4] | 348 | 8.5 [6.9 10.4] | 258 | 8.6 [6.9 10.5] | 0.449 |
| | Peak pressure | ml/Kg(RBW) | 599 | 26.0 [23.0 30.0] | 344 | 25.3 [22.0 29.0] | 255 | 27.0 [24.0 30.0] | 0.000 |
| | Plateau pressure | ml/Kg(IBW) | 80 | 26.0 [22.5 28.5] | 38 | 24.0 [21.0 28.0] | 42 | 26.5 [24.0 29.0] | 0.012 |
| | PEEP | cmH_20 | 603 | 10.0 [8.0 12.0] | 347 | 10.0 [8.0 12.0] | 256 | 10.0 [10.0 12.0] | 0.000 |
| | Mean airway pressure | cmH_20 | 387 | 16.0 [13.2 19.0] | 239 | 15.0 [13.0 18.0] | 148 | 17.0 [14.0 20.0] | 0.001 |
| | Pressure support | cmH_20 | 371 | 10.0 [5.3 14.0] | 217 | 10.0 [6.0 14.0] | 154 | 10.0 [5.0 12.0] | 0.516 |
| | Dynamic Compliance | mls/cmH20 | 564 | 31.5 [24.3 40.2] | 327 | 32.5 [25.4 41.2] | 237 | 30.0 [23.3 39.2] | 0.033 |
| | Oxygenation Index | | 387 | 8.1 [5.1 12.5] | 239 | 7.1 [4.7 10.4] | 148 | 9.7 [6.4 14.4] | 0.000 |
| | Ventilatory Ratio | | 470 | 1.5 [1.2 2.1] | 284 | 1.5 [1.2 2.0] | 186 | 1.6 [1.3 2.1] | 0.033 |
| **ABG** | Oxygen saturation | | 518 | 95.0 [93.0 98.0] | 308 | 96.0 [93.0 98.0] | 210 | 95.0 [92.0 97.0] | 0.006 |
| | pH | | 630 | 7.4 [7.3 7.4] | 363 | 7.4 [7.3 7.4] | 267 | 7.3 [7.3 7.4] | 0.000 |
| | $PaO_2$ | kPa | 630 | 10.7 [9.2 13.1] | 363 | 10.7 [9.1 12.8] | 267 | 10.9 [9.2 13.4] | 0.441 |
| | $PaCO_2$ | kPa | 630 | 6.0 [5.2 7.2] | 363 | 5.9 [5.2 6.9] | 267 | 6.1 [5.2 7.3] | 0.136 |
| | Base excess | | 630 | -0.3 [-2.6 2.2] | 363 | 0.2 [-2.1 2.5] | 267 | -0.9 [-3.2 1.5] | 0.001 |
| | $HCO_3^-$ | mmol/L | 629 | 24.5 [22.5 26.7] | 362 | 24.9 [22.9 27.0] | 267 | 23.9 [21.9 26.0] | 0.000 |
| | Lactate | mmol/L | 604 | 1.2 [1.0 1.6] | 347 | 1.1 [0.9 1.5] | 257 | 1.3 [1.1 1.8] | 0.000 |
| **SOFA** | SOFA score | | 428 | 9.0 [7.0 11.0] | 246 | 8.0 [6.0 11.0] | 182 | 10.0 [8.0 12.0] | 0.000 |
| | SOFA Respiratory | | 626 | 3.0 [3.0 4.0] | 359 | 3.0 [2.0 3.0] | 267 | 3.0 [3.0 4.0] | 0.000 |
| | SOFA Nervous | | 508 | 3.0 [0.0 4.0] | 301 | 0.0 [0.0 4.0] | 207 | 4.0 [0.0 4.0] | 0.024 |
| | SOFA Cardiovascular | | 579 | 3.0 [3.0 4.0] | 328 | 3.0 [3.0 3.0] | 251 | 3.0 [3.0 4.0] | 0.000 |
| | SOFA Liver | | 588 | 0.0 [0.0 0.0] | 335 | 0.0 [0.0 0.0] | 253 | 0.0 [0.0 0.0] | 0.678 |
| | SOFA Coagulation | | 619 | 0.0 [0.0 0.0] | 355 | 0.0 [0.0 0.0] | 264 | 0.0 [0.0 0.0] | 0.004 |
| | SOFA Kidneys | | 629 | 0.0 [0.0 1.0] | 362 | 0.0 [0.0 1.0] | 267 | 0.0 [0.0 1.0] | 0.007 |
| **FBC** | Haemoglobin | g/dL | 619 | 114.0 [92.0 128.0] | 355 | 115.0 [95.0 128.0] | 264 | 113.0 [85.8 129.0] | 0.380 |
| | Haematocrit | | 312 | 0.4 [0.3 0.4] | 168 | 0.4 [0.3 0.4] | 144 | 0.4 [0.3 0.4] | 0.030 |
| | White blood cell count | x10^9/L | 619 | 9.6 [7.0 13.1] | 355 | 9.4 [6.9 12.6] | 264 | 9.9 [7.3 13.5] | 0.209 |
| | Neutrophils | x10^9/L | 618 | 8.1 [5.7 11.3] | 355 | 7.6 [5.3 10.7] | 263 | 8.6 [6.2 12.3] | 0.010 |
| | Monocytes | x10^9/L | 614 | 0.4 [0.3 0.7] | 355 | 0.5 [0.3 0.7] | 259 | 0.4 [0.2 0.7] | 0.003 |
| | Lymphocytes | x10^9/L | 615 | 0.8 [0.5 1.2] | 355 | 0.9 [0.6 1.3] | 260 | 0.7 [0.5 1.0] | 0.000 |
| | Basophils | x10^9/L | 498 | 0.0 [0.0 0.1] | 281 | 0.0 [0.0 0.1] | 217 | 0.0 [0.0 0.0] | 0.001 |
| | Eosinophils | x10^9/L | 493 | 0.0 [0.0 0.1] | 286 | 0.0 [0.0 0.1] | 207 | 0.0 [0.0 0.1] | 0.000 |
| **Coag** | Platelet Count | μmol/L | 619 | 246.0 [185.3 320.8] | 355 | 261.0 [199.0 334.8] | 264 | 229.5 [171.0 305.5] | 0.001 |
| | APTT | U/L | 398 | 32.1 [28.3 37.4] | 209 | 32.0 [28.7 37.1] | 189 | 32.1 [28.0 37.5] | 0.603 |
| | PT | U/L | 396 | 13.9 [12.4 15.2] | 209 | 13.9 [12.5 15.1] | 187 | 13.8 [12.4 15.5] | 0.713 |
| | INR | U/L | 212 | 1.1 [1.1 1.2] | 113 | 1.1 [1.1 1.2] | 99 | 1.1 [1.1 1.2] | 0.616 |
| | Fibrinogen | U/L | 410 | 6.8 [5.6 8.1] | 246 | 6.8 [5.8 8.0] | 164 | 6.7 [5.4 8.2] | 0.321 |
| | D-dimer | IU/L | 391 | 2642.0 [990.5 7701.3] | 221 | 2270.0 [902.3 5680.0] | 170 | 3290.5 [1239.0 15575.0] | 0.002 |
| **Electrolytes** | Blood Urea Nitrogen (BUN) | mmol/L | 498 | 7.4 [4.9 11.8] | 277 | 6.8 [4.7 10.4] | 221 | 8.4 [5.7 13.3] | 0.000 |
| | Creatinine | μmol/L | 621 | 88.0 [66.0 140.0] | 356 | 82.0 [63.0 123.0] | 265 | 99.0 [71.8 152.1] | 0.000 |
| | Sodium | mmol/L | 621 | 139.0 [136.0 142.0] | 357 | 139.0 [136.0 142.0] | 264 | 138.0 [135.0 141.0] | 0.071 |
| | Potassium | mmol/L | 620 | 4.4 [4.0 4.8] | 356 | 4.4 [4.0 4.8] | 264 | 4.4 [4.1 4.8] | 0.462 |
| **Liver** | Bilirubin | μmol/L | 588 | 10.0 [7.0 15.0] | 335 | 10.0 [7.0 14.0] | 253 | 10.0 [7.0 15.0] | 0.068 |
| | Alkaline Phosphatase | U/L | 600 | 77.0 [58.5 113.0] | 342 | 76.0 [53.0 116.0] | 258 | 79.5 [62.0 110.0] | 0.341 |
| | AST | U/L | 99 | 59.0 [39.3 85.0] | 50 | 53.0 [37.0 63.0] | 49 | 70.0 [43.3 96.8] | 0.018 |
| | ALT | U/L | 592 | 37.0 [24.0 59.0] | 338 | 37.0 [23.0 62.0] | 254 | 37.0 [24.0 56.0] | 0.820 |
| **Inflammation** | LDH (Lactate dehydrogenase) | IU/L | 194 | 649.0 [452.0 921.0] | 109 | 614.0 [397.3 818.8] | 85 | 683.0 [506.3 977.3] | 0.079 |
| | Ferritin | ng/mL | 290 | 1218.5 [696.0 2320.0] | 173 | 1144.0 [681.5 2056.3] | 117 | 1410.0 [714.8 2814.0] | 0.025 |
| | CRP | | 592 | 215.7 [135.0 311.0] | 350 | 200.5 [117.4 298.0] | 242 | 242.2 [159.0 325.6] | 0.000 |
| | Procalcitonin | ug/L | 125 | 0.7 [0.3 2.2] | 86 | 0.5 [0.3 1.3] | 39 | 1.6 [0.6 5.5] | 0.000 |
| **Cardiac** | Creatinine Kinase | U/L | 243 | 217.0 [83.5 637.3] | 132 | 202.0 [84.0 578.8] | 111 | 222.0 [82.8 639.5] | 0.733 |
| | High sensitivity Troponin | | 253 | 21.8 [11.0 61.0] | 147 | 16.0 [9.0 41.3] | 106 | 29.4 [13.0 103.0] | 0.001 |
| | NT Pro BNP | pg/ml | 58 | 537.5 [165.0 1478.0] | 37 | 349.0 [139.0 823.5] | 21 | 857.0 [565.0 3416.8] | 0.005 |
| **Fluid** | Cumulative Fluid balance | L | 589 | 343.0 [-212.3 1058.3] | 342 | 273.4 [-262.0 938.0] | 247 | 413.0 [-182.4 1360.0] | 0.033 |
| **Adjunctive interventions** | | | | | | | | | |
| | Was patient transferred in? | | 633 | 159 (25.1%) | 365 | 103 (28.2%) | 268 | 56 (20.9%) | 0.036 |
| | Tracheostomy | | 516 | 145 (28.1%) | 305 | 122 (40%) | 211 | 23 (10.9%) | 0.000 |
| | PEEP>10 | | 622 | 459 (73.8%) | 359 | 231 (64.3%) | 263 | 228 (86.7%) | 0.000 |
| | Neuro-muscular blockade | | 617 | 434 (70.3%) | 354 | 215 (60.7%) | 263 | 219 (83.3%) | 0.000 |
| | Prone positioning | | 551 | 273 (49.5%) | 328 | 129 (39.3%) | 223 | 144 (64.6%) | 0.000 |
| | Inhaled nitric oxide | | 521 | 73 (14%) | 309 | 29 (9.39%) | 212 | 44 (20.8%) | 0.001 |
| | Inhaled prostacyclin | | 521 | 55 (10.6%) | 309 | 30 (9.71%) | 212 | 25 (11.8%) | 0.624 |
| | Bronchoscopy | | 521 | 51 (9.79%) | 309 | 33 (10.7%) | 212 | 18 (8.49%) | 0.288 |
| | Renal replacement therapy | | 555 | 211 (38%) | 322 | 89 (27.6%) | 233 | 122 (52.4%) | 0.000 |
| | Diuretics | | 598 | 443 (74.1%) | 348 | 256 (73.6%) | 250 | 187 (74.8%) | 0.922 |
| | Corticosteroids | | 582 | 304 (52.2%) | 342 | 173 (50.6%) | 240 | 131 (54.6%) | 0.712 |
| | Therapeutic heparin | | 438 | 55 (12.6%) | 255 | 30 (11.8%) | 183 | 25 (13.7%) | 0.624 |
| | Antibiotics | | 313 | 219 (70%) | 168 | 109 (64.9%) | 145 | 110 (75.9%) | 0.003 |
| | **Prone** | | | | | | | | |
| | No of proning episodes | | 273 | 1.0 [1.0 2.0] | 129 | 1.0 [1.0 2.0] | 144 | 1.0 [1.0 2.0] | 0.864 |
| | mech vent prior first prone | days | 273 | 3.0 [1.8 6.0] | 129 | 3.0 [1.0 6.0] | 144 | 4.0 [2.0 7.0] | 0.065 |
| | durationn of first prone | days | 273 | 2.0 [1.0 4.0] | 129 | 2.0 [1.0 4.0] | 144 | 2.0 [1.0 3.5] | 0.552 |
| | responders to proning | | 270 | 119 (44.1%) | 128 | 82 (64.1%) | 142 | 37 (26.1%) | 0.006 |
| | missed windows prior to prone | | 273 | 3.0 [1.0 6.0] | 129 | 2.0 [0.8 5.0] | 144 | 3.0 [1.0 7.0] | 0.020 |
| | missed windows - unproned | | 360 | 3.0 [1.0 10.0] | 236 | 2.0 [0.0 6.0] | 124 | 7.0 [3.0 15.5] | 0.000 |
| | **Tracheostomy** | | | | | | | | |
| | mech vent prior to tracheostomy | days | 145 | 13.0 [9.0 18.0] | 122 | 14.0 [10.0 18.0] | 23 | 13.0 [4.0 18.8] | 0.127 |
| | durationn of tracheostomy | days | 145 | 14.0 [6.0 21.3] | 122 | 15.0 [7.0 22.0] | 23 | 6.0 [2.3 12.8] | 0.001 |
| **End of life parameters** | | | | | | | | | |
| | life sustaining therapy withdrawn | | 130 | 85 (65.4%) | 0 | 0 (NaN%) | 130 | 85 (65.4%) | 0.000 |
| | cardiac arrest during admission | | 244 | 21 (8.61%) | 122 | 8 (6.56%) | 122 | 13 (10.7%) | 0.065 |

#### Table S14 - Time series mixed model ANOVA according to ICU outcome.

| | | p-values | | | |
|---|---|---|---|---|---|
| **label** | **N** | **Dead vs Alive** | **parameter over time** | **interaction** | **FDR interaction** |
| PaO_2 / FiO_2 | 472 | 0.000 | 0.000 | **0.000** | **0.000** |
| Dynamic Compliance | 341 | 0.001 | 0.000 | **0.000** | **0.000** |
| Respiratory rate | 458 | 0.062 | 0.001 | 0.561 | 0.632 |
| FiO_2 (%) | 503 | 0.000 | 0.000 | **0.000** | **0.000** |
| Tidal Volume per Kg | 347 | 0.993 | 0.343 | 0.537 | 0.632 |
| Tidal Volume | 433 | 0.285 | 0.388 | 0.699 | 0.724 |
| Minute ventilation | 404 | 0.039 | 0.000 | 0.285 | 0.405 |
| Peak pressure | 426 | 0.000 | 0.000 | **0.000** | **0.000** |
| PEEP | 390 | 0.000 | 0.001 | 0.104 | 0.204 |
| Dynamic delta pressure (peakP-peep) | 124 | 0.001 | 0.058 | **0.009** | **0.022** |
| Mean airway pressure | 256 | 0.000 | 0.000 | **0.000** | **0.000** |
| I:E ratio | 93 | 0.050 | 0.552 | 0.261 | 0.391 |
| Pressure support | 169 | 0.021 | 0.045 | 0.063 | 0.134 |
| Oxygen saturation | 417 | 0.000 | 0.125 | **0.002** | **0.007** |
| pH | 487 | 0.000 | 0.000 | **0.000** | **0.000** |
| PaO_2 | 484 | 0.482 | 0.274 | 0.167 | 0.280 |
| PaCO_2 | 487 | 0.000 | 0.105 | **0.000** | **0.000** |
| Base excess | 478 | 0.000 | 0.000 | 0.405 | 0.513 |
| HCO^-3 | 479 | 0.000 | 0.000 | 0.380 | 0.504 |
| Lactate | 451 | 0.000 | 0.187 | **0.011** | **0.026** |
| Oxygenation Index | 237 | 0.000 | 0.000 | **0.000** | **0.000** |
| Ventilatory Ratio | 314 | 0.009 | 0.000 | 0.116 | 0.212 |
| Mean arterial pressure (lowest) | 386 | 0.000 | 0.000 | **0.000** | **0.000** |
| Liver-Bilirubin | 167 | 0.017 | 0.103 | 0.248 | 0.382 |
| Daily fluid balance | 480 | 0.000 | 0.000 | 0.348 | 0.472 |
| Cumulative fluid balance | 480 | 0.000 | 0.006 | **0.000** | **0.000** |
| SOFA Score | 192 | 0.000 | 0.000 | **0.000** | **0.000** |
| Non-Respiratory SOFA | 224 | 0.000 | 0.000 | **0.000** | **0.000** |
| Glucose | 465 | 0.021 | 0.020 | 0.114 | 0.212 |
| BUN | 382 | 0.000 | 0.004 | **0.001** | **0.002** |
| Creatinine | 520 | 0.000 | 0.120 | **0.000** | **0.000** |
| Sodium | 523 | 0.706 | 0.000 | 0.393 | 0.509 |
| Potassium | 518 | 0.000 | 0.781 | **0.008** | **0.020** |
| Bilirubin | 401 | 0.003 | 0.000 | **0.000** | **0.000** |
| Alkaline Phosphatase | 441 | 0.893 | 0.000 | 0.339 | 0.471 |
| AST | 55 | 0.988 | 0.517 | 0.559 | 0.632 |
| ALT | 418 | 0.092 | 0.000 | 0.231 | 0.366 |
| Creatinine Kinase | 127 | 0.643 | 0.275 | 0.467 | 0.579 |
| LDH | 66 | 0.845 | 0.001 | 0.283 | 0.405 |
| Haemoglobin | 515 | 0.001 | 0.080 | 0.081 | 0.165 |
| WBC | 513 | 0.017 | 0.003 | **0.014** | **0.031** |
| RBC | 415 | 0.256 | 0.486 | 0.535 | 0.632 |
| Platelet Count | 511 | 0.000 | 0.000 | **0.001** | **0.002** |
| Haematocrit | 252 | 0.754 | 0.000 | **0.001** | **0.003** |
| Neutrophils | 511 | 0.000 | 0.019 | **0.002** | **0.006** |
| Monocytes | 509 | 0.000 | 0.000 | 0.138 | 0.238 |
| Lymphocytes | 509 | 0.000 | 0.000 | **0.008** | **0.020** |
| Basophils | 361 | 0.020 | 0.007 | 0.570 | 0.632 |
| Eosinophils | 386 | 0.000 | 0.000 | 0.614 | 0.660 |
| APTT | 255 | 0.008 | 0.223 | **0.002** | **0.005** |
| PT | 260 | 0.671 | 0.632 | 0.654 | 0.691 |
| INR | 163 | 0.895 | 0.954 | 0.836 | 0.836 |
| Fibrinogen | 137 | 0.518 | 0.007 | 0.577 | 0.632 |
| Ferritin | 75 | 0.123 | 0.184 | 0.119 | 0.212 |
| D-dimer | 147 | 0.000 | 0.949 | 0.772 | 0.785 |
| CRP | 457 | 0.000 | 0.000 | **0.007** | **0.019** |
| High sensitivity Troponin | 62 | 0.204 | 0.069 | 0.198 | 0.323 |

#### Table S15- Uni- and multi-variate model analysis of factors associated with ICU mortality.

| | | Univariate | | | | Multivariate | | | |
|---|---|---|---|---|---|---|---|---|---|
| **FieldLabel** | **median [IQR]** | **Odds ratio** | **95% CI** | | **P Value** | **Odds ratio** | **95% CI** | | **P Value** |
| Age | | 2.017 | 1.694 | 2.401 | 0.000 | 1.951 | 1.584 | 2.402 | 0.000 |
| Male | | 1.560 | 1.032 | 2.357 | 0.035 | 2.052 | 1.168 | 3.607 | 0.012 |
| BMI | 28.13 [24.90 32.76] | 0.850 | 0.724 | 0.997 | 0.046 | 0.844 | 0.687 | 1.038 | 0.108 |
| Height | 173.00 [165.00 178.00] | 0.929 | 0.788 | 1.095 | 0.381 | | | | |
| symptoms days | 8.00 [6.00 12.00] | 0.880 | 0.745 | 1.040 | 0.133 | | | | |
| Hypertension | | 1.318 | 0.932 | 1.863 | 0.119 | | | | |
| Diabetes mellitus | | 1.191 | 0.815 | 1.741 | 0.366 | | | | |
| Oxygen saturation | 95.00 [93.00 98.00] | 0.816 | 0.695 | 0.959 | 0.013 | 1.078 | 0.872 | 1.332 | 0.487 |
| pH | 7.36 [7.30 7.42] | 0.796 | 0.683 | 0.928 | 0.004 | 0.849 | 0.670 | 1.074 | 0.172 |
| PaCO_2 | 5.96 [5.16 7.15] | 1.056 | 0.907 | 1.230 | 0.484 | | | | |
| HCO^-3 | 24.50 [22.48 26.70] | 0.764 | 0.654 | 0.892 | 0.001 | 0.807 | 0.641 | 1.018 | 0.070 |
| Lactate | 1.20 [1.00 1.60] | 1.522 | 1.294 | 1.791 | 0.000 | 1.523 | 1.210 | 1.917 | 0.000 |
| Peak pressure | 26.00 [23.00 30.00] | 1.240 | 1.061 | 1.449 | 0.007 | 0.963 | 0.731 | 1.268 | 0.788 |
| PEEP | 10.00 [8.00 12.00] | 1.407 | 1.175 | 1.685 | 0.000 | 1.150 | 0.896 | 1.475 | 0.272 |
| Minute ventilation | 8.53 [6.88 10.44] | 0.967 | 0.831 | 1.125 | 0.663 | | | | |
| Dynamic Comp | 31.47 [24.26 40.19] | 0.860 | 0.735 | 1.005 | 0.058 | 0.706 | 0.543 | 0.919 | 0.010 |
| Oxygenation Index | 8.10 [5.09 12.45] | 1.371 | 1.160 | 1.621 | 0.000 | 1.204 | 0.903 | 1.605 | 0.205 |
| Ventilatory Ratio | 1.54 [1.22 2.07] | 1.152 | 0.983 | 1.349 | 0.081 | 1.151 | 0.911 | 1.453 | 0.238 |
| Cum fluid balance | 343.00 [-212.25 1058.32] | 1.163 | 0.997 | 1.356 | 0.055 | 0.874 | 0.715 | 1.068 | 0.188 |
| Glucose | 8.39 [6.79 10.95] | 1.246 | 1.065 | 1.457 | 0.006 | 1.067 | 0.876 | 1.300 | 0.521 |
| BUN | 7.40 [4.90 11.80] | 1.412 | 1.202 | 1.657 | 0.000 | 1.058 | 0.805 | 1.392 | 0.685 |
| Sodium | 139.00 [136.00 142.00] | 0.880 | 0.754 | 1.026 | 0.103 | | | | |
| Potassium | 4.40 [4.00 4.80] | 1.040 | 0.891 | 1.213 | 0.620 | | | | |
| ALP | 77.00 [58.50 113.00] | 1.056 | 0.908 | 1.227 | 0.480 | | | | |
| ALT | 37.00 [24.00 59.00] | 0.966 | 0.830 | 1.124 | 0.652 | | | | |
| Haemoglobin | 114.00 [92.00 128.00] | 0.928 | 0.800 | 1.078 | 0.329 | | | | |
| Neutrophils | 8.10 [5.71 11.30] | 1.206 | 1.034 | 1.407 | 0.017 | 1.078 | 0.858 | 1.354 | 0.520 |
| Monocytes | 0.41 [0.30 0.70] | 0.794 | 0.682 | 0.925 | 0.003 | 0.838 | 0.671 | 1.046 | 0.119 |
| Lymphocytes | 0.80 [0.50 1.20] | 0.700 | 0.593 | 0.827 | 0.000 | 0.967 | 0.771 | 1.213 | 0.773 |
| Basophils | 0.00 [0.00 0.10] | 0.465 | 0.304 | 0.712 | 0.000 | 0.684 | 0.391 | 1.195 | 0.182 |
| Eosinophils | 0.00 [0.00 0.10] | 0.400 | 0.276 | 0.579 | 0.000 | 0.442 | 0.267 | 0.733 | 0.002 |
| APTT | 32.10 [28.30 37.40] | 0.968 | 0.817 | 1.148 | 0.710 | | | | |
| PT | 13.85 [12.40 15.20] | 1.105 | 0.937 | 1.303 | 0.236 | | | | |
| Fibrinogen | 6.80 [5.60 8.10] | 0.953 | 0.808 | 1.125 | 0.571 | | | | |
| D-dimer | 2642.00 [990.50 7701.25] | 1.356 | 1.149 | 1.602 | 0.000 | 1.164 | 0.941 | 1.439 | 0.161 |
| CRP | 215.65 [135.00 311.00] | 1.286 | 1.100 | 1.505 | 0.002 | 1.039 | 0.842 | 1.281 | 0.722 |
| SOFA Respiratory | | 1.419 | 1.160 | 1.736 | 0.001 | 1.235 | 0.858 | 1.776 | 0.256 |
| SOFA Nervous | | 1.100 | 1.008 | 1.201 | 0.032 | 1.110 | 0.877 | 1.405 | 0.386 |
| SOFA Cardio | | 1.227 | 1.050 | 1.433 | 0.010 | 1.115 | 0.861 | 1.445 | 0.409 |
| SOFA Liver | | 1.079 | 0.787 | 1.479 | 0.638 | | | | |
| SOFA Coagulation | | 1.590 | 1.138 | 2.221 | 0.007 | 1.951 | 1.167 | 3.262 | 0.011 |
| SOFA Kidneys | | 1.192 | 1.017 | 1.399 | 0.031 | 1.172 | 0.836 | 1.643 | 0.356 |
| SOFA score | | 1.195 | 1.113 | 1.284 | 0.000 | 0.921 | 0.740 | 1.147 | 0.463 |

* Died (N=235) vs Discharged (N=314)

#### Table S16 - Time series mixed model ANOVA according to ARDS on admission

| | | p-values | | | |
|---|---|---|---|---|---|
| **label** | **N** | **ARDS Group** | **parameter over time** | **interaction** | **FDR interaction** |
| PaO_2 / FiO_2 | 472 | 0.000 | 0.000 | **0.000** | **0.000** |
| Dynamic Compliance | 341 | 0.000 | 0.011 | 0.304 | 0.656 |
| Respiratory rate | 458 | 0.118 | 0.000 | 0.820 | 0.922 |
| FiO_2 (%) | 503 | 0.000 | 0.000 | **0.000** | **0.000** |
| Tidal Volume per Kg | 347 | 0.506 | 0.267 | 0.854 | 0.922 |
| Tidal Volume | 433 | 0.606 | 0.119 | 0.811 | 0.922 |
| Minute ventilation | 404 | 0.041 | 0.000 | 0.750 | 0.913 |
| Peak pressure | 426 | 0.000 | 0.000 | **0.000** | **0.003** |
| PEEP | 390 | 0.000 | 0.103 | 0.566 | 0.798 |
| Dynamic delta pressure (peakP-peep) | 124 | 0.022 | 0.001 | 0.227 | 0.656 |
| Mean airway pressure | 256 | 0.000 | 0.173 | 0.117 | 0.450 |
| I:E ratio | 93 | 0.015 | 0.656 | 0.461 | 0.798 |
| Pressure support | 169 | 0.415 | 0.878 | 0.917 | 0.934 |
| Oxygen saturation | 417 | 0.001 | 0.000 | **0.001** | **0.007** |
| pH | 487 | 0.000 | 0.014 | 0.574 | 0.798 |
| PaO_2 | 484 | 0.000 | 0.000 | **0.000** | **0.000** |
| PaCO_2 | 487 | 0.000 | 0.000 | **0.043** | 0.196 |
| Base excess | 478 | 0.022 | 0.000 | 0.577 | 0.798 |
| HCO^-3 | 479 | 0.001 | 0.000 | 0.525 | 0.798 |
| Lactate | 451 | 0.046 | 0.000 | **0.009** | 0.065 |
| Oxygenation Index | 237 | 0.000 | 0.000 | **0.000** | **0.000** |
| Ventilatory Ratio | 314 | 0.003 | 0.000 | 0.381 | 0.736 |
| Mean arterial pressure (lowest) | 386 | 0.610 | 0.045 | 0.847 | 0.922 |
| Liver-Bilirubin | 167 | 0.574 | 0.133 | 0.294 | 0.656 |
| Daily fluid balance | 480 | 0.042 | 0.194 | 0.281 | 0.656 |
| Cumulative fluid balance | 480 | 0.013 | 0.424 | 0.145 | 0.497 |
| SOFA Score | 192 | 0.019 | 0.712 | 0.740 | 0.913 |
| Non-Respiratory SOFA | 224 | 0.019 | 0.066 | 0.163 | 0.517 |
| Glucose | 465 | 0.290 | 0.312 | 0.761 | 0.913 |
| BUN | 382 | 0.075 | 0.000 | 0.532 | 0.798 |
| Creatinine | 520 | 0.314 | 0.841 | **0.011** | 0.065 |
| Sodium | 523 | 0.431 | 0.000 | 0.820 | 0.922 |
| Potassium | 518 | 0.139 | 0.853 | **0.010** | 0.065 |
| Bilirubin | 401 | 0.124 | 0.192 | 0.706 | 0.913 |
| Alkaline Phosphatase | 441 | 0.116 | 0.000 | 0.147 | 0.497 |
| AST | 55 | 0.018 | 0.729 | 0.626 | 0.845 |
| ALT | 418 | 0.912 | 0.000 | **0.014** | 0.076 |
| Creatinine Kinase | 127 | 0.433 | 0.796 | 0.719 | 0.913 |
| Haemoglobin | 515 | 0.978 | 0.147 | 0.083 | 0.343 |
| WBC | 513 | 0.751 | 0.002 | 0.487 | 0.798 |
| RBC | 415 | 0.049 | 0.743 | 0.967 | 0.967 |
| Platelet Count | 511 | 0.191 | 0.000 | 0.264 | 0.656 |
| Haematocrit | 252 | 0.190 | 0.001 | **0.039** | 0.192 |
| Neutrophils | 511 | 0.054 | 0.020 | 0.288 | 0.656 |
| Monocytes | 509 | 0.002 | 0.000 | 0.563 | 0.798 |
| Lymphocytes | 509 | 0.256 | 0.001 | 0.558 | 0.798 |
| Basophils | 361 | 0.883 | 0.002 | 0.369 | 0.736 |
| Eosinophils | 386 | 0.899 | 0.000 | 0.539 | 0.798 |
| APTT | 255 | 0.146 | 0.309 | 0.248 | 0.656 |
| PT | 260 | 0.322 | 0.697 | 0.469 | 0.798 |
| INR | 163 | 0.002 | 0.741 | 0.907 | 0.934 |
| Fibrinogen | 137 | 0.711 | 0.090 | 0.916 | 0.934 |
| D-dimer | 147 | 0.027 | 0.504 | 0.282 | 0.656 |
| CRP | 457 | 0.000 | 0.039 | 0.326 | 0.676 |

### Table S17 - Missing values for each parameter – by site

| Site | All | A | B | C | D | E | F | G | H | I | J | K | L | M | N | O | P | Q | R |
|---|---|---|---|---|---|---|---|---|---|---|---|---|---|---|---|---|---|---|---|
| N | **633** | **2** | **1** | **18** | **35** | **49** | **19** | **36** | **57** | **34** | **34** | **3** | **64** | **54** | **5** | **9** | **31** | **9** | **173** |
| % of Missing Data | | | | | | | | | | | | | | | | | | | |
| PaO_2 / FiO_2 | 6% | 0% | 2% | 5% | 3% | 8% | 7% | 1% | 10% | 4% | 22% | 0% | 1% | 6% | 0% | 0% | 5% | 5% | 5% |
| Oxygen saturation | 21% | 7% | 0% | 7% | 0% | 1% | 100% | 100% | 100% | 0% | 2% | 0% | 1% | 0% | 0% | 0% | 0% | 0% | 0% |
| pH | 5% | 0% | 2% | 4% | 1% | 2% | 6% | 1% | 10% | 4% | 13% | 0% | 1% | 6% | 0% | 0% | 3% | 6% | 4% |
| PaCO_2 | 5% | 0% | 2% | 4% | 1% | 2% | 6% | 1% | 10% | 4% | 13% | 0% | 1% | 6% | 0% | 0% | 3% | 6% | 4% |
| Base excess | 6% | 0% | 2% | 5% | 2% | 3% | 17% | 2% | 12% | 4% | 14% | 0% | 2% | 7% | 0% | 0% | 3% | 6% | 4% |
| HCO^-3 | 5% | 0% | 2% | 4% | 2% | 2% | 13% | 1% | 10% | 5% | 13% | 0% | 1% | 6% | 0% | 0% | 3% | 6% | 5% |
| Lactate | 9% | 0% | 100% | 4% | 3% | 2% | 6% | 1% | 10% | 5% | 13% | 0% | 1% | 7% | 0% | 0% | 90% | 6% | 5% |
| Peak pressure | 13% | 0% | 4% | 12% | 14% | 12% | 7% | 7% | 4% | 11% | 17% | 8% | 4% | 6% | 0% | 1% | 10% | 3% | 21% |
| PEEP | 14% | 0% | 2% | 16% | 20% | 9% | 0% | 2% | 0% | 6% | 18% | 0% | 2% | 4% | 0% | 1% | 4% | 2% | 30% |
| Mean airway pressure | 41% | 0% | 25% | 100% | 22% | 14% | 82% | 28% | 12% | 96% | 20% | 32% | 100% | 97% | 0% | 22% | 58% | 100% | 32% |
| Tidal Volume per Kg | 30% | 100% | 8% | 51% | 14% | 28% | 8% | 22% | 89% | 6% | 14% | 0% | 15% | 37% | 67% | 97% | 47% | 86% | 21% |
| Respiratory rate | 12% | 0% | 57% | 11% | 13% | 34% | 0% | 0% | 0% | 0% | 10% | 0% | 0% | 0% | 0% | 0% | 5% | 0% | 20% |
| Minute ventilation | 15% | 0% | 62% | 34% | 14% | 37% | 8% | 6% | 3% | 2% | 16% | 0% | 4% | 4% | 0% | 1% | 21% | 2% | 22% |
| Dynamic Compliance | 19% | 0% | 9% | 37% | 20% | 14% | 13% | 9% | 5% | 16% | 20% | 0% | 6% | 4% | 3% | 1% | 26% | 5% | 31% |
| Oxygenation Index | 43% | 0% | 25% | 100% | 22% | 16% | 82% | 29% | 20% | 96% | 25% | 32% | 100% | 97% | 0% | 22% | 59% | 100% | 32% |
| Ventilatory Ratio | 34% | 100% | 62% | 54% | 15% | 48% | 14% | 23% | 90% | 9% | 20% | 0% | 16% | 39% | 67% | 97% | 48% | 86% | 22% |
| SOFA Score | 47% | 13% | 96% | 21% | 22% | 17% | 65% | 54% | 68% | 58% | 81% | 84% | 76% | 55% | 6% | 47% | 80% | 59% | 31% |
| Non-Respiratory SOFA | 44% | 13% | 96% | 18% | 18% | 8% | 63% | 52% | 59% | 56% | 78% | 84% | 76% | 52% | 6% | 47% | 79% | 55% | 29% |
| Cumulative fluid balance | 11% | 7% | 100% | 5% | 4% | 7% | 3% | 4% | 28% | 0% | 10% | 4% | 3% | 2% | 3% | 1% | 83% | 0% | 4% |
| Glucose | 12% | 13% | 100% | 10% | 1% | 3% | 6% | 1% | 3% | 8% | 21% | 12% | 4% | 61% | 6% | 4% | 90% | 2% | 2% |
| BUN | 26% | 0% | 100% | 19% | 10% | 100% | 8% | 4% | 5% | 4% | 100% | 8% | 7% | 3% | 3% | 10% | 100% | 8% | 11% |
| Creatinine | 6% | 0% | 4% | 6% | 6% | 3% | 8% | 4% | 5% | 4% | 6% | 8% | 4% | 3% | 3% | 6% | 6% | 8% | 7% |
| Sodium | 5% | 0% | 4% | 7% | 7% | 1% | 5% | 1% | 3% | 4% | 6% | 8% | 3% | 3% | 3% | 3% | 6% | 8% | 7% |
| Potassium | 5% | 0% | 4% | 6% | 7% | 4% | 5% | 1% | 3% | 4% | 6% | 8% | 4% | 3% | 6% | 3% | 6% | 9% | 8% |
| Bilirubin | 16% | 13% | 11% | 6% | 14% | 2% | 19% | 5% | 21% | 8% | 6% | 12% | 3% | 37% | 6% | 46% | 60% | 47% | 14% |
| Alkaline Phosphatase | 11% | 0% | 11% | 10% | 9% | 2% | 8% | 4% | 5% | 11% | 6% | 12% | 2% | 32% | 6% | 27% | 61% | 33% | 8% |
| AST | 92% | 13% | 100% | 97% | 100% | 100% | 100% | 100% | 100% | 16% | 98% | 76% | 22% | 100% | 78% | 99% | 97% | 100% | 100% |
| ALT | 13% | 13% | 11% | 6% | 11% | 2% | 9% | 9% | 6% | 7% | 6% | 12% | 3% | 38% | 3% | 46% | 60% | 68% | 10% |
| Creatinine Kinase | 71% | 100% | 98% | 99% | 100% | 8% | 83% | 40% | 71% | 14% | 26% | 48% | 8% | 99% | 33% | 100% | 87% | 91% | 100% |
| LDH | 80% | 47% | 98% | 95% | 97% | 11% | 100% | 100% | 99% | 17% | 50% | 96% | 51% | 74% | 8% | 99% | 88% | 98% | 95% |
| Haemoglobin | 5% | 0% | 4% | 2% | 6% | 1% | 8% | 4% | 5% | 3% | 7% | 8% | 2% | 4% | 3% | 4% | 6% | 20% | 7% |
| WBC | 5% | 0% | 4% | 2% | 6% | 1% | 8% | 4% | 5% | 3% | 7% | 8% | 2% | 4% | 3% | 5% | 6% | 20% | 7% |
| RBC | 25% | 0% | 4% | 2% | 6% | 1% | 100% | 100% | 100% | 6% | 7% | 8% | 4% | 5% | 3% | 5% | 6% | 20% | 7% |
| Platelet Count | 6% | 0% | 4% | 2% | 6% | 1% | 9% | 4% | 5% | 5% | 7% | 8% | 2% | 4% | 3% | 6% | 6% | 20% | 8% |
| Haematocrit | 63% | 73% | 4% | 5% | 100% | 1% | 100% | 100% | 100% | 3% | 7% | 8% | 2% | 3% | 3% | 5% | 6% | 20% | 100% |
| Neutrophils | 6% | 0% | 4% | 3% | 6% | 1% | 9% | 6% | 6% | 3% | 7% | 8% | 3% | 5% | 6% | 6% | 6% | 20% | 7% |
| Monocytes | 6% | 0% | 4% | 3% | 7% | 1% | 9% | 6% | 6% | 3% | 7% | 8% | 4% | 5% | 6% | 6% | 6% | 20% | 7% |
| Lymphocytes | 6% | 0% | 4% | 3% | 6% | 1% | 9% | 6% | 6% | 4% | 7% | 8% | 4% | 5% | 6% | 6% | 6% | 20% | 7% |
| Basophils | 20% | 0% | 4% | 3% | 33% | 1% | 9% | 6% | 6% | 4% | 7% | 8% | 4% | 6% | 6% | 6% | 6% | 21% | 44% |
| Eosinophils | 16% | 0% | 4% | 3% | 28% | 1% | 9% | 6% | 6% | 4% | 7% | 8% | 4% | 6% | 6% | 6% | 6% | 20% | 34% |
| APTT | 50% | 27% | 9% | 3% | 100% | 3% | 10% | 6% | 6% | 14% | 13% | 8% | 8% | 53% | 17% | 42% | 45% | 74% | 100% |
| PT | 50% | 27% | 9% | 3% | 100% | 3% | 10% | 6% | 7% | 14% | 12% | 8% | 7% | 54% | 8% | 44% | 45% | 74% | 100% |
| INR | 75% | 100% | 100% | 2% | 100% | 3% | 98% | 99% | 96% | 11% | 12% | 8% | 6% | 100% | 8% | 97% | 100% | 100% | 100% |
| Fibrinogen | 50% | 27% | 40% | 16% | 23% | 10% | 100% | 100% | 100% | 99% | 34% | 52% | 22% | 55% | 17% | 70% | 56% | 76% | 38% |
| Ferritin | 63% | 47% | 98% | 75% | 57% | 18% | 100% | 100% | 100% | 19% | 58% | 68% | 44% | 72% | 31% | 32% | 80% | 100% | 56% |
| D-dimer | 48% | 60% | 100% | 72% | 63% | 17% | 11% | 11% | 11% | 21% | 58% | 60% | 49% | 73% | 6% | 31% | 93% | 94% | 63% |
| Triglicerides | 77% | 93% | 94% | 96% | 58% | 99% | 100% | 100% | 100% | 97% | 60% | 56% | 88% | 81% | 89% | 54% | 86% | 95% | 59% |
| CRP | 11% | 7% | 4% | 20% | 6% | 2% | 7% | 5% | 5% | 13% | 6% | 16% | 2% | 74% | 6% | 4% | 6% | 94% | 8% |
| Procalcitonin | 80% | 87% | 98% | 79% | 57% | 93% | 100% | 100% | 100% | 98% | 94% | 64% | 91% | 99% | 100% | 61% | 91% | 61% | 59% |
| High sensitivity Troponin | 64% | 80% | 100% | 79% | 55% | 7% | 100% | 100% | 100% | 92% | 47% | 52% | 44% | 73% | 33% | 43% | 88% | 98% | 55% |
| NT Pro BNP | 97% | 80% | 100% | 78% | 97% | 100% | 100% | 100% | 100% | 97% | 89% | 100% | 100% | 100% | 97% | 98% | 98% | 97% | 96% |

### Table S18 - Missing values for each parameter – by day

| Site | Day 0 | Day 1 | Day 2 | Day 3 | Day 4 | Day 5 | Day 6 | Day 7 | Day 8 | Day 9 | Day10 | Day11 | Day12 | Day13 | Day14 | Day15 | Day16 | Day17 | Day18 | Day19 | Day20 | Day21 |
|---|---|---|---|---|---|---|---|---|---|---|---|---|---|---|---|---|---|---|---|---|---|---|
| N | **633** | **616** | **588** | **570** | **547** | **522** | **486** | **469** | **435** | **404** | **377** | **344** | **321** | **300** | **275** | **255** | **234** | **219** | **202** | **185** | **174** | **162** |
| % of Missing Data | | | | | | | | | | | | | | | | | | | | | | |
| PaO_2 / FiO_2 | 5% | 2% | 2% | 3% | 3% | 2% | 2% | 3% | 4% | 3% | 5% | 3% | 5% | 5% | 5% | 6% | 6% | 6% | 9% | 6% | 8% | 8% |
| Oxygen saturation | 19% | 19% | 18% | 18% | 18% | 19% | 19% | 19% | 18% | 19% | 20% | 22% | 21% | 22% | 21% | 23% | 22% | 22% | 22% | 23% | 24% | 23% |
| pH | 2% | 1% | 2% | 2% | 3% | 2% | 2% | 3% | 3% | 2% | 3% | 2% | 3% | 3% | 3% | 4% | 4% | 5% | 5% | 4% | 6% | 7% |
| PaCO_2 | 2% | 1% | 2% | 2% | 3% | 2% | 2% | 3% | 3% | 2% | 3% | 2% | 3% | 3% | 3% | 4% | 4% | 5% | 5% | 4% | 6% | 7% |
| Base excess | 3% | 1% | 2% | 3% | 4% | 3% | 2% | 4% | 4% | 3% | 4% | 3% | 4% | 5% | 4% | 5% | 6% | 6% | 7% | 5% | 7% | 9% |
| HCO^-3 | 3% | 1% | 2% | 3% | 3% | 2% | 2% | 3% | 3% | 3% | 4% | 3% | 3% | 3% | 3% | 5% | 6% | 5% | 6% | 5% | 7% | 7% |
| Lactate | 7% | 5% | 6% | 7% | 7% | 6% | 7% | 8% | 9% | 7% | 8% | 8% | 9% | 9% | 8% | 9% | 9% | 9% | 9% | 8% | 11% | 10% |
| Peak pressure | 12% | 6% | 4% | 5% | 6% | 8% | 6% | 7% | 8% | 7% | 7% | 9% | 12% | 10% | 9% | 12% | 14% | 12% | 13% | 11% | 15% | 17% |
| PEEP | 10% | 6% | 8% | 9% | 11% | 12% | 9% | 11% | 12% | 12% | 11% | 11% | 11% | 13% | 11% | 11% | 14% | 13% | 13% | 14% | 16% | 18% |
| Mean airway pressure | 46% | 39% | 37% | 37% | 38% | 39% | 36% | 37% | 39% | 37% | 36% | 35% | 36% | 35% | 36% | 38% | 40% | 39% | 41% | 41% | 41% | 41% |
| Tidal Volume per Kg | 32% | 25% | 24% | 25% | 27% | 27% | 24% | 26% | 26% | 26% | 27% | 28% | 30% | 29% | 28% | 29% | 29% | 28% | 28% | 26% | 28% | 30% |
| Respiratory rate | 5% | 1% | 3% | 4% | 6% | 8% | 6% | 6% | 8% | 7% | 8% | 10% | 12% | 10% | 10% | 11% | 13% | 11% | 12% | 14% | 14% | 17% |
| Minute ventilation | 14% | 5% | 6% | 8% | 9% | 11% | 8% | 9% | 11% | 10% | 11% | 12% | 14% | 14% | 14% | 16% | 19% | 17% | 15% | 16% | 19% | 20% |
| Dynamic Compliance | 21% | 13% | 12% | 13% | 13% | 16% | 11% | 15% | 17% | 16% | 14% | 15% | 16% | 16% | 15% | 16% | 18% | 18% | 17% | 17% | 20% | 20% |
| Oxygenation Index | 48% | 40% | 38% | 39% | 40% | 40% | 37% | 38% | 40% | 38% | 38% | 36% | 36% | 36% | 38% | 39% | 41% | 41% | 44% | 42% | 44% | 44% |
| Ventilatory Ratio | 34% | 26% | 26% | 28% | 29% | 30% | 27% | 28% | 30% | 29% | 30% | 31% | 32% | 31% | 31% | 33% | 35% | 32% | 30% | 30% | 33% | 38% |
| SOFA Score | 75% | 40% | 39% | 42% | 41% | 42% | 41% | 44% | 43% | 44% | 41% | 43% | 42% | 47% | 42% | 46% | 45% | 44% | 46% | 45% | 49% | 48% |
| Non-Respiratory SOFA | 71% | 38% | 36% | 37% | 38% | 39% | 38% | 42% | 41% | 42% | 38% | 40% | 40% | 43% | 38% | 44% | 43% | 40% | 42% | 43% | 44% | 42% |
| Cumulative fluid balance | 47% | 7% | 7% | 8% | 7% | 7% | 7% | 10% | 9% | 8% | 9% | 9% | 9% | 8% | 8% | 9% | 8% | 8% | 9% | 6% | 7% | 6% |
| Glucose | 12% | 12% | 11% | 12% | 10% | 10% | 10% | 11% | 10% | 11% | 11% | 12% | 12% | 11% | 11% | 10% | 12% | 11% | 12% | 10% | 10% | 6% |
| BUN | 50% | 22% | 21% | 21% | 20% | 21% | 24% | 23% | 26% | 24% | 25% | 26% | 27% | 27% | 25% | 27% | 29% | 26% | 28% | 28% | 25% | 25% |
| Creatinine | 35% | 3% | 1% | 2% | 2% | 2% | 2% | 2% | 3% | 2% | 3% | 4% | 3% | 3% | 2% | 5% | 5% | 3% | 3% | 2% | 1% | 4% |
| Sodium | 34% | 3% | 1% | 2% | 1% | 1% | 1% | 2% | 2% | 2% | 3% | 3% | 2% | 3% | 2% | 4% | 3% | 3% | 3% | 2% | 1% | 2% |
| Potassium | 35% | 3% | 1% | 2% | 2% | 1% | 1% | 2% | 3% | 2% | 3% | 3% | 3% | 2% | 3% | 4% | 3% | 4% | 3% | 2% | 1% | 2% |
| Bilirubin | 38% | 12% | 13% | 11% | 11% | 11% | 13% | 14% | 12% | 13% | 12% | 15% | 13% | 15% | 12% | 17% | 16% | 17% | 15% | 16% | 14% | 14% |
| Alkaline Phosphatase | 36% | 10% | 10% | 8% | 8% | 7% | 10% | 9% | 8% | 8% | 8% | 10% | 9% | 10% | 8% | 9% | 9% | 8% | 8% | 8% | 8% | 7% |
| AST | 88% | 87% | 86% | 85% | 86% | 87% | 87% | 89% | 89% | 89% | 90% | 95% | 95% | 96% | 95% | 95% | 94% | 97% | 97% | 97% | 98% | 99% |
| ALT | 38% | 11% | 11% | 10% | 9% | 9% | 11% | 11% | 10% | 10% | 10% | 12% | 11% | 13% | 10% | 13% | 12% | 11% | 10% | 10% | 10% | 9% |
| Creatinine Kinase | 70% | 67% | 67% | 65% | 67% | 66% | 67% | 67% | 66% | 68% | 67% | 72% | 71% | 73% | 71% | 67% | 72% | 71% | 76% | 71% | 73% | 71% |
| LDH | 78% | 77% | 77% | 76% | 75% | 73% | 76% | 77% | 74% | 76% | 76% | 79% | 81% | 79% | 80% | 79% | 80% | 82% | 83% | 83% | 82% | 80% |
| Haemoglobin | 34% | 3% | 1% | 2% | 1% | 2% | 1% | 1% | 2% | 2% | 2% | 4% | 3% | 3% | 2% | 4% | 6% | 4% | 3% | 3% | 3% | 3% |
| WBC | 34% | 3% | 1% | 2% | 1% | 2% | 1% | 1% | 2% | 2% | 2% | 4% | 3% | 3% | 2% | 4% | 6% | 4% | 3% | 3% | 3% | 3% |
| RBC | 52% | 22% | 19% | 20% | 19% | 20% | 20% | 20% | 20% | 21% | 22% | 25% | 24% | 24% | 23% | 25% | 25% | 25% | 25% | 25% | 26% | 25% |
| Platelet Count | 34% | 3% | 2% | 2% | 2% | 3% | 1% | 1% | 2% | 2% | 2% | 4% | 4% | 2% | 3% | 4% | 6% | 4% | 3% | 4% | 3% | 2% |
| Haematocrit | 56% | 53% | 53% | 54% | 54% | 55% | 55% | 56% | 56% | 58% | 59% | 63% | 64% | 63% | 63% | 63% | 64% | 65% | 65% | 66% | 67% | 69% |
| Neutrophils | 34% | 3% | 2% | 2% | 1% | 3% | 2% | 1% | 2% | 3% | 3% | 5% | 4% | 3% | 3% | 4% | 6% | 4% | 5% | 4% | 3% | 3% |
| Monocytes | 35% | 4% | 2% | 2% | 2% | 3% | 2% | 1% | 2% | 3% | 3% | 5% | 4% | 3% | 3% | 4% | 6% | 4% | 5% | 4% | 3% | 4% |
| Lymphocytes | 35% | 4% | 2% | 2% | 1% | 3% | 2% | 1% | 2% | 3% | 3% | 5% | 4% | 3% | 3% | 4% | 6% | 4% | 5% | 4% | 3% | 3% |
| Basophils | 38% | 24% | 23% | 25% | 23% | 23% | 21% | 21% | 18% | 22% | 19% | 20% | 19% | 16% | 16% | 15% | 15% | 13% | 14% | 11% | 12% | 14% |
| Eosinophils | 38% | 24% | 18% | 16% | 12% | 13% | 13% | 13% | 17% | 17% | 16% | 19% | 18% | 16% | 13% | 14% | 14% | 14% | 11% | 6% | 8% | 9% |
| APTT | 42% | 44% | 47% | 47% | 46% | 47% | 44% | 47% | 46% | 46% | 47% | 49% | 49% | 51% | 49% | 49% | 50% | 47% | 50% | 49% | 49% | 52% |
| PT | 42% | 44% | 47% | 46% | 45% | 46% | 44% | 47% | 46% | 46% | 47% | 50% | 49% | 51% | 48% | 49% | 52% | 48% | 51% | 50% | 49% | 52% |
| INR | 70% | 67% | 69% | 69% | 68% | 68% | 69% | 71% | 70% | 70% | 72% | 77% | 77% | 77% | 74% | 76% | 76% | 76% | 78% | 77% | 78% | 78% |
| Fibrinogen | 65% | 44% | 48% | 48% | 46% | 47% | 48% | 48% | 47% | 46% | 47% | 48% | 45% | 51% | 46% | 43% | 44% | 47% | 49% | 49% | 52% | 52% |
| Ferritin | 75% | 64% | 62% | 60% | 56% | 56% | 56% | 58% | 58% | 56% | 59% | 58% | 63% | 61% | 57% | 63% | 63% | 67% | 67% | 64% | 65% | 59% |
| D-dimer | 61% | 48% | 52% | 48% | 46% | 45% | 48% | 46% | 49% | 44% | 46% | 47% | 43% | 45% | 42% | 45% | 45% | 43% | 44% | 43% | 46% | 39% |
| Triglicerides | 89% | 78% | 79% | 77% | 77% | 78% | 76% | 78% | 78% | 76% | 76% | 73% | 72% | 79% | 71% | 77% | 74% | 74% | 77% | 74% | 74% | 73% |
| CRP | 39% | 13% | 11% | 9% | 10% | 8% | 9% | 8% | 8% | 9% | 7% | 9% | 8% | 8% | 7% | 7% | 9% | 8% | 8% | 8% | 5% | 7% |
| Procalcitonin | 95% | 84% | 85% | 82% | 82% | 80% | 80% | 81% | 81% | 78% | 80% | 78% | 79% | 80% | 75% | 78% | 74% | 77% | 77% | 70% | 77% | 71% |
| High sensitivity Troponin | 78% | 69% | 72% | 73% | 69% | 70% | 73% | 71% | 70% | 68% | 65% | 65% | 66% | 62% | 61% | 63% | 61% | 63% | 61% | 62% | 54% | 50% |
| NT Pro BNP | 96% | 94% | 96% | 97% | 96% | 96% | 96% | 97% | 98% | 97% | 98% | 97% | 97% | 97% | 98% | 95% | 97% | 97% | 98% | 97% | 98% | 98% |

#### Figure S1 – Adjunctive interventions application

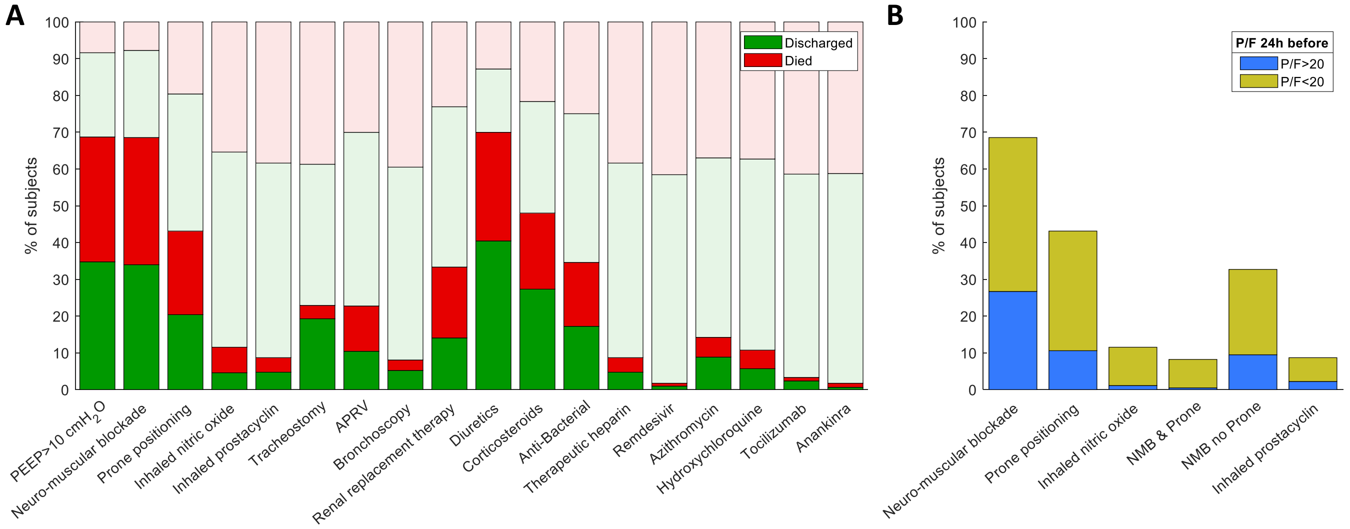

Figure S2 – Time-series comparison between resolvers and non- resolvers

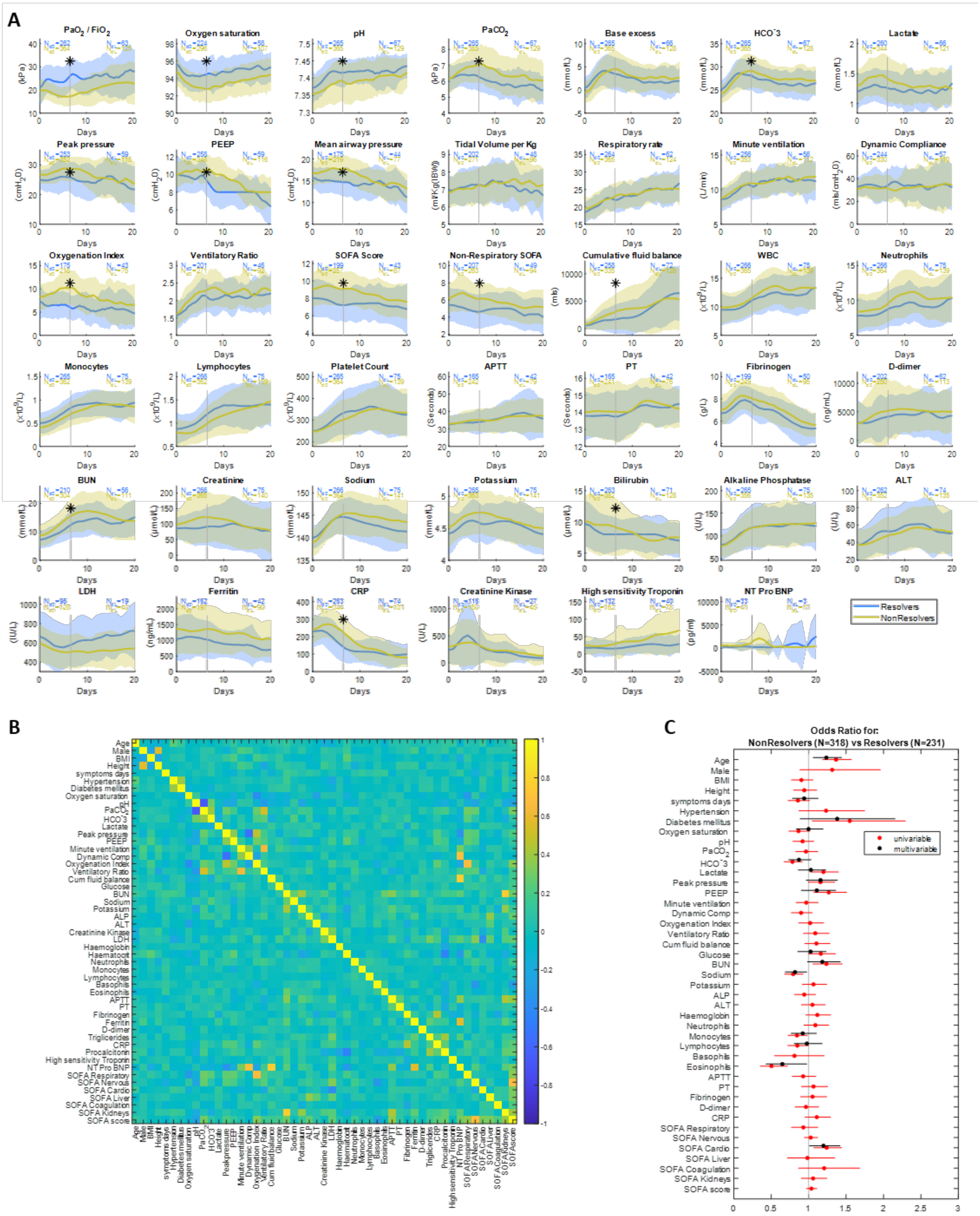

#### Figure S3 – Time-series comparison between prone responders and non-responders

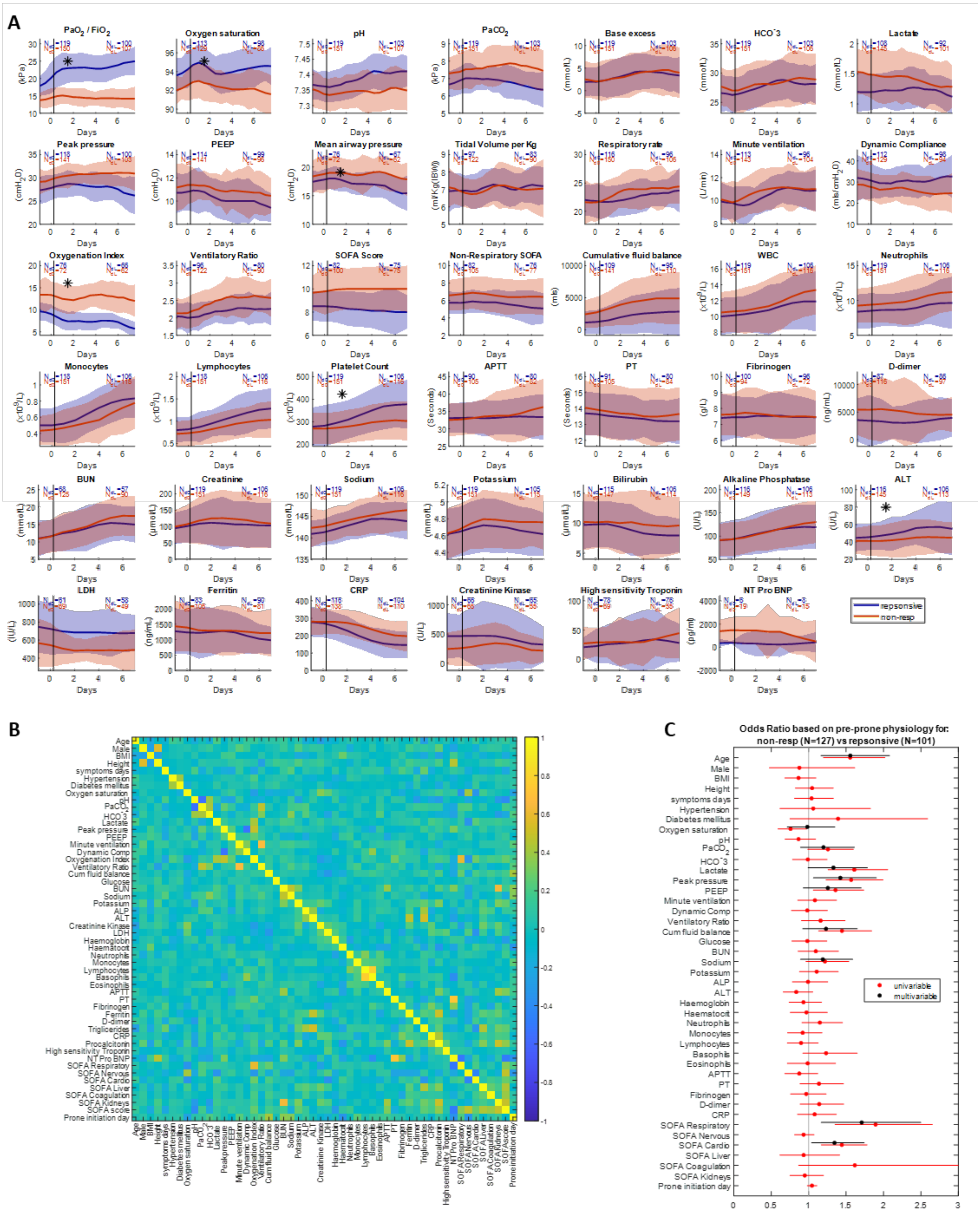

#### Figure S4 – Logistic Regression for prone response based on post-prone physiology

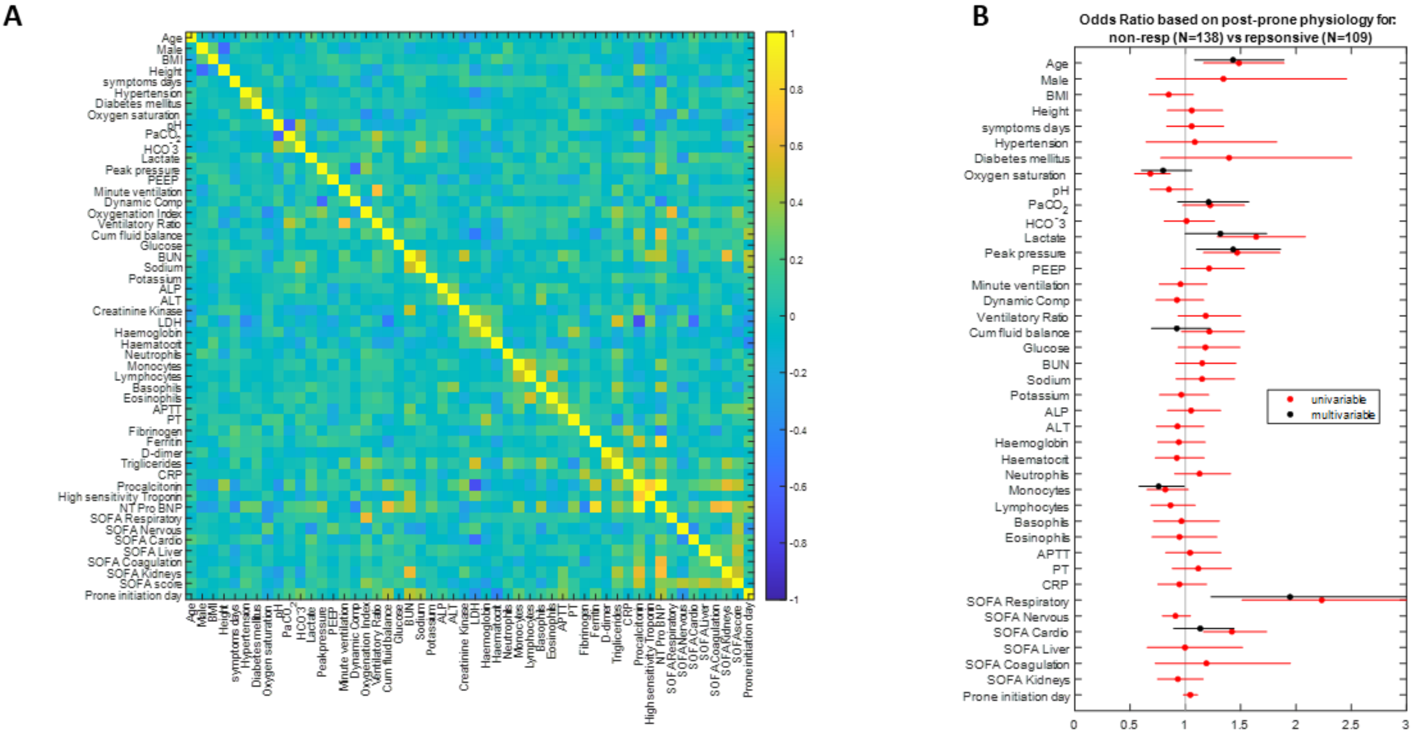

### Figure S5 – Time-series comparison between admission ARDS severities

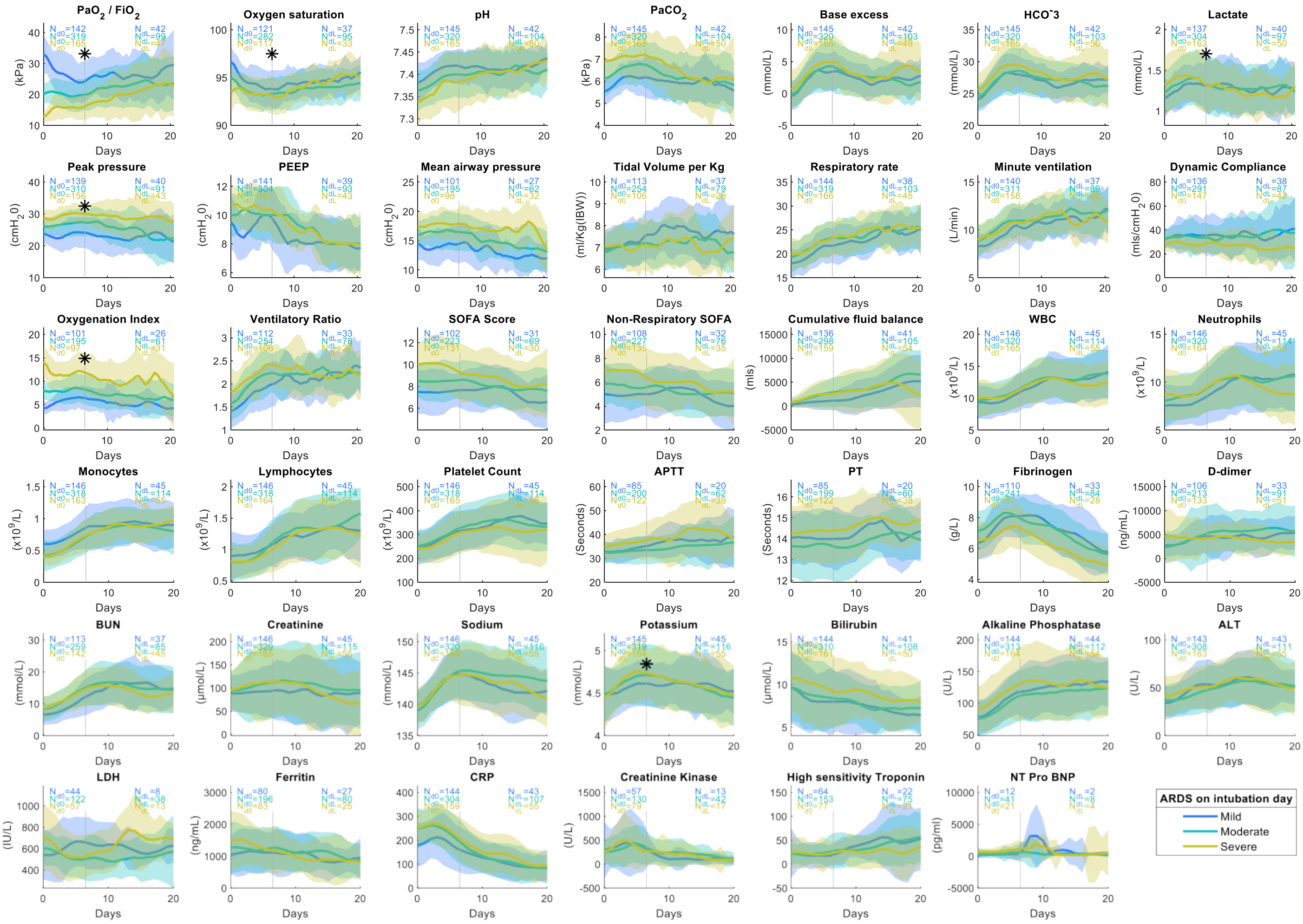

#### Figure S6 – Time series comparison between outcome groups

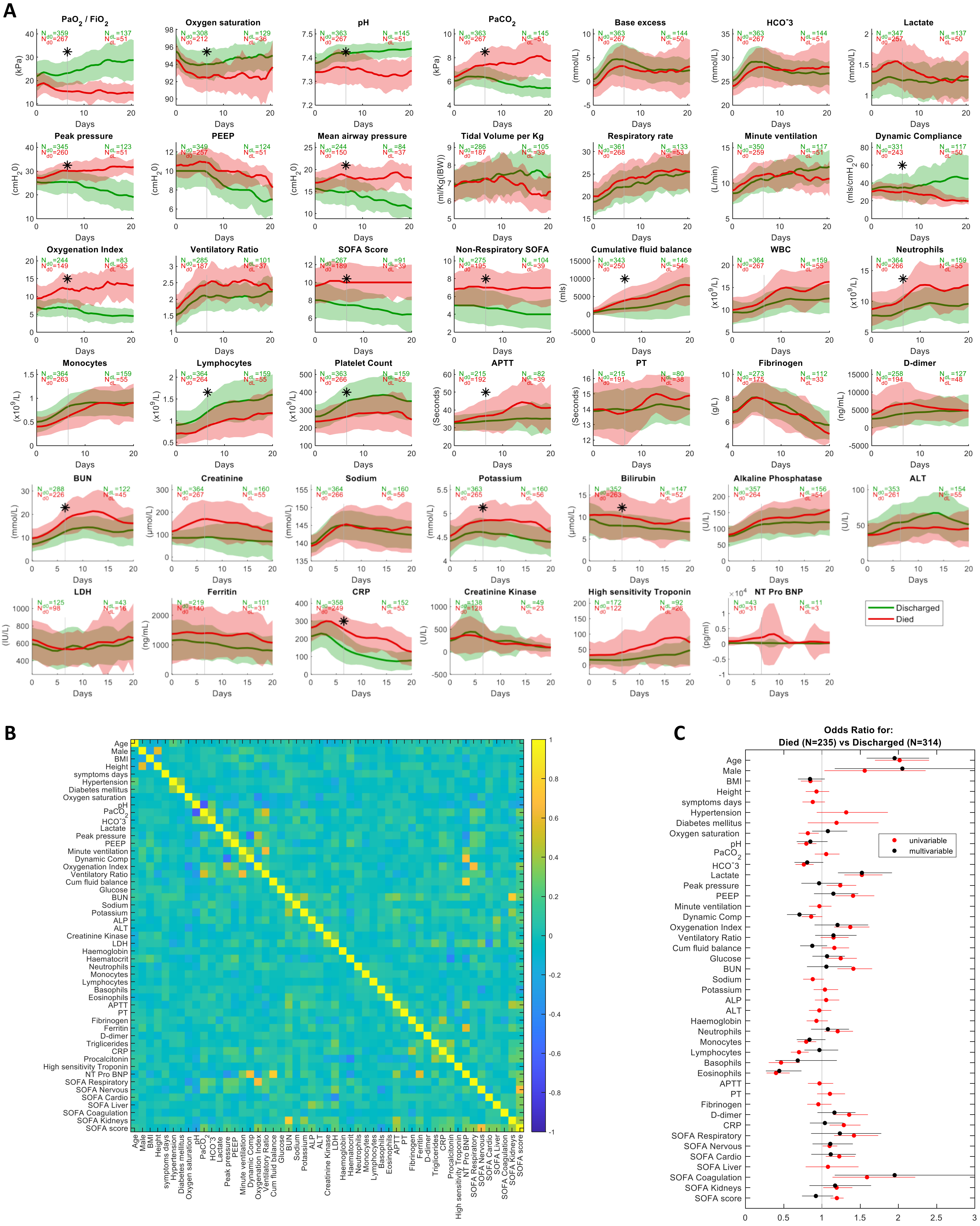

#### Figure S7 – Site variation of application of interventions

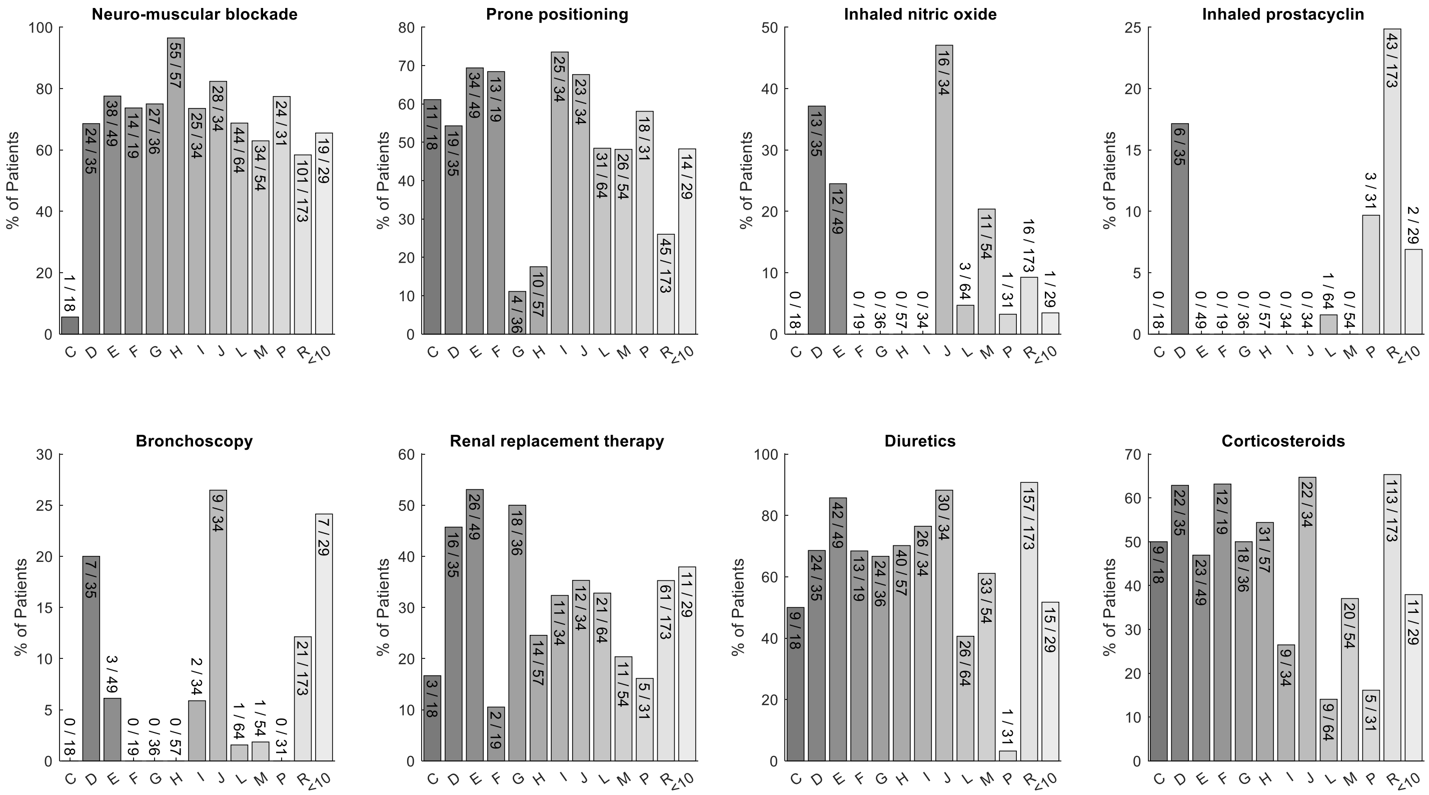

Figure S8 – Oxygenation Index dependency on P/F ratio

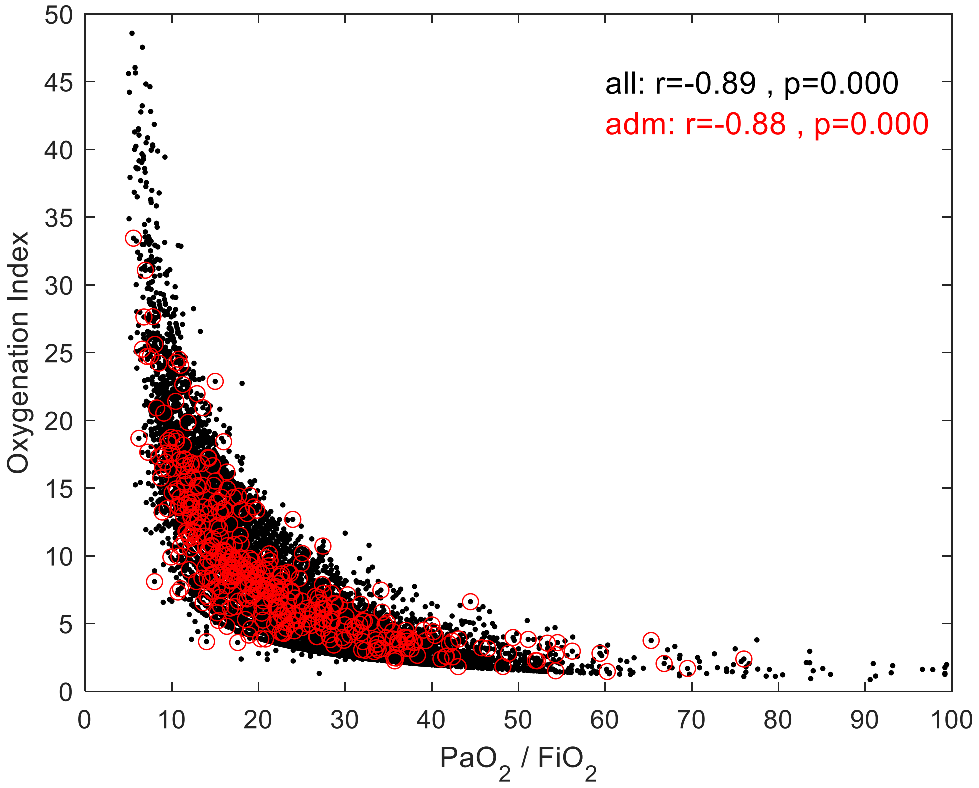
